# Risk of Bias in Psychotherapy Trials for Depression A large-scale empirical examination using a structured assessment tool developed for psychotherapy research

**DOI:** 10.1101/2025.06.26.25330349

**Authors:** Clara Miguel, Mathias Harrer, Ethan Sahker, Masatsugu Sakata, Lingyao Tong, Olga-Maria Panagiotopoulou, Yingying Wang, Marketa Ciharova, Paula Kuper, Antonia A. Sprenger, Esra Kaya Aglio, Rie Toyomoto, Keisuke Yoshimura, Aran Tajika, Eirini Karyotaki, Toshi A. Furukawa, Pim Cuijpers

**Author notes:** **Correspondence to:** Clara Miguel.

## Abstract

Risk of bias (RoB) in psychotherapy trials may inflate effect size estimates, yet its prevalence, trends over time, and associations with trial characteristics and effect sizes have not been comprehensively examined in the psychotherapy literature. We therefore applied Metapsy RoB, a new structured 24-item tool developed for this project and tailored to psychotherapy research, to 514 randomized controlled trials of psychotherapy for adult depression from a living meta-analytic database, with each trial independently rated by a pair of raters. Reliability was strong (mean item-level Cohen’s κ = 0.86; domain-level Gwet’s AC2 = 0.68–0.93). Risk of bias was widespread: only 14.4% of trials were rated low risk, while 52.1% were rated high risk. Measurement of the outcome was most often rated low risk (93.2%), whereas missing outcome data was most often rated high risk (36%). High-risk trials became less common over time, with low-risk trials emerging from 2009 onwards. In proportional odds regression, larger trials and guided self-help interventions were associated with lower RoB. Using correlated and hierarchical effects models, pooled effect sizes were larger in trials at higher risk of bias (Hedges’ *g* = 0.48 for low risk vs. 0.82 for high risk; Δ*g* = −0.34, 95% CI [−0.45, −0.23]). These findings suggest that effect estimates from psychotherapy trials for depression are substantially larger when trials are at high risk of bias. Even among low-risk trials, however, psychotherapy probably shows a meaningful benefit. Systematic reviews should routinely stratify analyses by risk of bias.

**Public Significance Statement:** This study of 514 trials of psychotherapy for depression found that most had weaknesses in how they were run or reported that can bias results, and only about one in seven was at low risk of bias. Trials at higher risk of bias were associated with larger treatment effects, although psychotherapy still showed a benefit in the trials at low risk of bias. Decisions about psychotherapy should therefore consider how trustworthy the supporting trials are.

## Introduction

Systematic reviews and meta-analyses of randomized controlled trials (RCTs) are considered the gold standard in evidence-based decision making, significantly influencing clinical guidelines and policies. The trustworthiness of their conclusions ultimately depends on the quality of the included primary studies. RCTs are considered the best design to examine the effects of an intervention due to their ability to provide causal evidence. However, flaws in the design or execution of a trial, such as improper randomization or selective reporting, may lead to systematic errors or deviations from the true effect of an intervention, known as biases (Sterne et al., 2019). These biases can result in distorted estimates of treatment efficacy, either by under-or overestimation of effects, and must be carefully evaluated in any well-conducted systematic review. The most widely used framework to evaluate risk of bias in RCTs is Cochrane’s Risk of Bias 2 tool (Sterne et al., 2019), which assesses five domains covering the randomization process, deviations from intended interventions, missing outcome data, measurement of the outcome, and selective outcome reporting, typically by two independent raters who resolve discrepancies through discussion or by consulting a third reviewer.

Evaluating the risk of bias (RoB) is particularly challenging in psychotherapy research, where standard double-blind RCT procedures are not easily applicable. Unlike drug trials, psychotherapy trials cannot achieve full blinding, as therapists and participants cannot be masked to the treatment being delivered, and control groups do not have a placebo equivalent (Cuijpers, 2025). However, non-blinding does not automatically imply bias. The critical question is whether departures from blinding led to deviations from intended interventions that affected the outcome. Similarly, prospective trial registration has historically been rare in psychotherapy research (Bradley et al., 2017; Miguel et al., 2021), making the assessment of selective outcome reporting particularly challenging. Attrition is common in psychotherapy trials and often poorly handled (Juul et al., 2024), and the most commonly used outcome measures are self-reported scales, for which there is no clear consensus on how to rate measurement bias in the context of unblinded administration. As a result, researchers frequently apply very different criteria for RoB ratings or skip entire RoB domains, leading to inconsistent and potentially misleading conclusions in clinical guidelines (Munder & Barth, 2018). Existing RoB frameworks have also proven difficult to apply reliably in practice, and domains that were particularly difficult to rate include missing outcome data, measurement of the outcome in unblinded trials, and deviations from intended interventions (Crocker et al., 2023; Minozzi et al., 2020), which are precisely the domains that pose the greatest interpretive challenges in psychotherapy trials. Developing implementation guidance tailored to the research context has been shown to reduce rating time and improve inter-rater reliability (Crocker et al., 2023; Minozzi et al., 2022; Tomlinson et al., 2024), but to the best of our knowledge no such guidance exists specifically for psychotherapy trials. Reliable RoB assessment is further complicated in large-scale meta-analytic projects, where many raters must apply consistent criteria across hundreds of trials, making structured and reproducible approaches particularly important.

Methodological weaknesses leading to risk of bias in psychotherapy trials can distort the conclusions of evidence syntheses. Meta-research has suggested that such biases can inflate effect size estimates (Cuijpers & Cristea, 2015), yet the prevalence of risk of bias, its trends over time, and its association with effect size estimates have not been comprehensively examined across the complete body of psychotherapy trials for depression. Such an examination can provide important insights into which methodological weaknesses are most common in the field, whether trial quality has improved over time, and how different types of bias are associated with effect size estimates. This information is critical for interpreting existing evidence and for guiding the design of future trials. Depression is one of the most studied conditions in psychotherapy research (Harrer et al., 2025), making it an ideal starting point for such an examination.

In the current study, we applied a structured RoB assessment tool developed specifically for this project to a complete cohort of RCTs on psychotherapy for depression, drawn from a living meta-analytic database (Cuijpers et al., 2023). The tool was developed as a precondition for this project to address, on one hand, the particular challenges of psychotherapy trials described above, and on the other, the reliability demands in large-scale meta-analytic work involving many raters across hundreds of trials. Our objectives were designed to provide an initial empirical examination of the tool and its findings: 1) to examine the inter-rater reliability of the ratings in a subsample of trials; 2) to provide an overview of the prevalence of different types of biases in the field; 3) to examine overall RoB and domain-specific changes over time; 4) to test the association between RoB and trial characteristics; and 5) to test the association between RoB and effect size estimates.

## Metʜod

### Identification, Selection of Studies, and Data Extraction

Randomized controlled trials were obtained from a living database on psychotherapy for depression (Cuijpers et al., 2023), part of the Metapsy initiative (www.metapsy.org; osf.io/825c6/), a series of living, open meta-analytic databases of interventions for mental health problems built on a unified methodology and infrastructure (Harrer et al., 2025). It is updated by running searches every four months in major bibliographic databases (PubMed, PsycINFO, Embase). We used the last update, conducted on 1st January 2026.

We included individually randomized controlled trials on adults with depression (elevated symptoms based on an established cut-off or diagnosed through a clinical interview) examining the effects of psychotherapy versus control conditions (waitlist, care as usual, attention placebo, etc.) on post-treatment depressive symptomatology, measured through valid self-reports or clinician-rated instruments. Psychotherapy was defined as “the informed and intentional application of clinical methods and interpersonal stances derived from established psychological principles for the purpose of assisting people to modify their behaviors, cognitions, emotions, and/or other personal characteristics in directions that the participants deem desirable” (Norcross, 1990). We included any type of psychotherapy (e.g., cognitive behavioral therapy, interpersonal psychotherapy, psychodynamic therapy) delivered in any format (individual, group, guided self-help) as long as it included human clinical support.

After studies were included, data were extracted on study characteristics (year of publication; country; income level of the country where the trial was conducted according to the World Bank classification, based on the year when the study was published); participant characteristics (target group; mean age; gender distribution), psychotherapy characteristics (type of psychotherapy; number of sessions; delivery format), and depression outcomes (means, standard deviations, and number of participants of any reported measurement of depressive symptoms at post-treatment). All review stages, from inclusion of studies to data extraction and RoB assessment, were completed by two independent reviewers. Further details about searches, inclusion and exclusion criteria, and data extraction procedures are available in Supplement A.

### Risk of Bias Assessment

Since the database was founded, the initial risk of bias of included studies had been rated using four items of the Cochrane risk of bias tool (version 1) (Higgins et al., 2011): sequence generation, allocation concealment, blinding of outcome assessors, and intention-to-treat analyses. Selective outcome reporting was also examined in a dedicated study (Miguel et al., 2021).

#### Development of Metapsy RoB

In this study, risk of bias was assessed using Metapsy RoB, a structured assessment tool developed specifically for this project, considering the particular challenges of psychotherapy trials and the reliability demands of large-scale meta-analytic work. Metapsy RoB was developed through an empirical and consensus-based process. Following a preliminary assessment of risk of bias in 35 trials using Cochrane’s RoB 2 (Sterne et al., 2019), which identified systematic challenges in applying the tool in the psychotherapy context, we conducted a year-long expert consensus process involving experienced psychotherapy researchers from the authorship team (PC, TAF, EK, ES, MS, CM), with additional input from external experts on specific methodological points. This process resulted in 24 structured items organized around the five most relevant bias domains identified in the RoB literature and RoB rating frameworks, including Cochrane’s RoB 2, accompanied by detailed psychotherapy-specific guidance. Item development and interpretation were guided by meta-epidemiological evidence, iterative piloting, and expert judgment, resulting in psychotherapy-specific decision rules intended to improve reliability and consistency across multiple raters in large-scale meta-analytic projects. A full description of Metapsy RoB, its items, scoring algorithms, and digital assistant tools is provided in Supplement B, C, and at metapsy.org/rob. Further details on the theoretical and empirical rationale for the interpretation of the ratings of each of the domains are provided in Supplement D.

#### Scoring Structure

The 24 items can be answered with Yes/Probably Yes (Yes/ PY), No/Probably No (No/PN), and No Information (NI). All items are formulated in the same direction, so that Yes/PY answers are always indicative of lower RoB. If wished, seven of the 24 items can be pre-filled automatically from variables already extracted as part of the data collection process, including the number of randomized participants per arm, baseline symptom severity, and attrition at posttreatment, using the digital helper tools within the metapsyTools R package (Harrer et al., 2022) (tools.metapsy.org/reference/createrobratings); otherwise, they can be completed manually. One item is a free-text field for the trial registration number, and the remaining 16 items are rated independently by each rater. Based on the answers to the 24 items, an RoB score for each domain can be calculated through the provided algorithms or by means of our digital helper tools (www.metapsy.org/ rob). These algorithms result in low risk, some concerns, or high-risk scores for each of the five domains: 1) randomization process, 2) deviations from intended interventions, 3) missing outcome data, 4) measurement of the outcome, and 5) selective outcome reporting, consistent with current risk of bias frameworks for randomized trials such as Cochrane’s RoB 2 (Sterne et al., 2019).

#### Specifications

Several specifications addressed particularities of psychotherapy trials not directly resolved by generic RoB guidance, and others reflected the reliability demands of applying the tool across a large number of raters and trials, for which fixed cut-offs (e.g., for sample sizes and proportion of missing data) were introduced as a deliberate compromise to support standardization. The key specification for each domain is summarized in Table 1. The extended rationale for each specification is available in Supplement D.

**Table 1.** Key Metapsy RoB specifications per domain.

| Domain | Items | Description | Level of assessment |
|---|---|---|---|
| **D1**<br>**Randomization process** | 1–4 | Assesses whether the allocation sequence was generated by a truly random method and adequately concealed, and whether baseline groups are balanced. Since current frameworks leave open which prognostic factors to examine for imbalance, two quantifiable indicators were selected: substantive imbalance in the number of randomized participants (allocated proportion and 99% CI, with intervals excluding the expected proportion considered suggestive of imbalance) and in baseline symptom severity (SMD and 99% CI, with SMD > 0.2 and lower 99% CI limit > 0 considered suggestive of imbalance), the latter chosen for its consistent association with treatment outcome across psychotherapy trials (Bower et al., 2013; Driessen et al., 2010; Furukawa et al., 2021; Karyotaki et al., 2018). | Study/comparison level, as information on randomization and baseline balance is reported at the trial level rather than separately for each instrument. |
| **D2**<br>**Deviations from intended interventions** | 5–9 | Since blinding of participants and personnel is essentially unfeasible in psychotherapy trials, all trials are assumed unblinded by default. However, non-blinding does not automatically imply bias. The absence of masking can nonetheless introduce bias through subtle psychological mechanisms, for instance when waitlist participants reduce their help-seeking behavior or motivation knowing they will eventually receive the active intervention. Waitlist controlled trials have consistently been associated with larger effect estimates than trials using more active control conditions (Furukawa et al., 2014; Michopoulos et al., 2021). Assessment therefore focuses on whether unbalanced deviations from protocolized interventions attributable to the trial context (Sterne et al., 2019), such as receipt of non-protocol co-interventions related to the outcome, occurred, rather than on blinding success per se. Per-protocol and as-treated analyses are considered inappropriate; a 5% exclusion threshold serves as a rough rule of thumb, though smaller exclusions may still matter depending on context. | Study/comparison level, as information on deviations from intended interventions and ITT analyses is reported at the trial level rather than separately for each instrument. |
| **D3**<br>**Missing outcome data** | 10–14 | Assesses whether missing outcome data could bias results. Bias is considered unlikely when outcome data are available for at least 95% of randomized participants. Where missingness exceeds this threshold, approved methods for handling its impact include mixed models for repeated measures using multiple assessment points, multiple imputation following best practice standards (Rubin's rules, relevant auxiliary variables, arm-specific imputation, or controlled imputation) (Barnard & Rubin, 1999; Little et al., 2012; Sullivan et al., 2018), and sensitivity analyses across a plausible range of assumptions about missing data. Last observation carried forward is not considered appropriate. When no approved method was used or reporting is unclear, indirect evidence is assessed via three indicators: whether reported reasons for missingness suggest a relationship with the outcome's true value, whether there is a large imbalance in missing data between groups (more than 20 percentage points as a rule of thumb), and whether reasons for missingness differ systematically between groups. | Study/comparison level, as attrition is almost universally reported as a single figure for all participants rather than separately by instrument, making instrument-level assessment impractical. |
| **D4**<br>**Measurement of the outcome** | 15–18 | Assesses whether outcomes were measured appropriately and consistently between groups, and whether the assessor's knowledge of treatment allocation could have influenced the measurement. When the appropriateness of the measurement method is unclear, some concerns is assigned. Self-reported outcomes are rated as low risk by default, based on evidence that they do not produce meaningfully larger effect estimates than masked clinician ratings in psychotherapy trials (Cuijpers, Li, et al., 2010; Miguel et al., 2025), provided there is no evidence or plausible risk that participants underreported their symptoms to please their therapist or researcher, for example by completing questionnaires in their presence. Unmasked clinician-administered assessments are rated as high risk, as unmasked assessors may unconsciously rate participants differently depending on their treatment condition. | Per instrument, as self-reports and clinician-rated scales within the same trial can lead to different risk of bias ratings for this domain. |
| **D5**<br>**Selective outcome reporting** | 19–24 | Assesses whether results were selectively reported based on their direction, magnitude, or statistical significance. Pre-specification of outcomes and analysis plan is assessed via the availability of a registration, protocol, or statistical analysis plan specifying at minimum the outcome scale and time point of interest. Where such prospective information exists, reviewers check whether all pre-specified outcomes were reported and whether the reported analysis matches the plan. If any instrument shows evidence of selective reporting at any level, the domain 5 rating for the whole study is conservatively assigned as high risk. | Study level, as complete omission of an outcome cannot be detected at the instrument level since omitted outcomes would not appear in the database. |

*Note*. SMD = standardized mean difference; CI = confidence interval; ITT = intention to treat. The full questionnaire with all 24 items is provided in Supplements B and C. Extended rationale and empirical background for each domain specification are provided in Supplement D.

#### Level of Assessment

Risk of bias was assessed for each numeric result of depressive symptom instruments at post-treatment, specifically the post-treatment means, standard deviations, and sample sizes from which the standardized mean difference between psychotherapy and control condition is calculated in the Metapsy depression database. When both post-treatment scores and change scores were available for the same instrument, post-treatment scores were prioritized. All assessments pertained to the effect of assignment to intervention (the intention-to-treat effect), which is the effect of interest for the Metapsy depression database. The level of assessment varied by domain, as detailed in Table 1.

#### Overall Risk of Bias Score

Overall low risk requires all five domains to be rated as low risk. High risk is assigned when at least one domain is rated as high risk, or when three or more domains have some concerns. Otherwise, the rating “some concerns” is assigned. The tool produced two overall RoB scores for each study: a score at the instrument level, which can differ across instruments due to domain 4, and a study-level score, which takes the most conservative rating across all instruments when a study reports more than one. Further details about the tool are provided in Supplement B.

### Rating Procedure

Pairs of researchers, all part of the co-author team, independently rated all included randomized trials. Discrepancies were resolved through discussion and, where needed, consulting a third researcher. In this study, the seven pre-fillable items were pre-filled using already collected data from the Metapsy database. The seven pre-filled items were included in the item-level kappa calculation, as they are part of the tool’s structured assessment; however, since they are identical by construction for both raters within each pair, a sensitivity analysis excluding them was conducted (further explanations in the ‘Statistical Analyses’ section).

Prior to independent rating, all raters read the guidance document and attended a 2-hour training workshop that included group discussion of example studies. Each rater then independently rated five trials, after which rater pairs met to discuss discrepancies and calibrate their understanding of the rating criteria. Inter-rater reliability was assessed in a subsample of 237 trials selected at the outset of the project, excluding trials already involved in existing rating processes, rated by 16 raters organized in 10 independent pairs. Further details are reported in Supplement E.

### Effect Size Calculation

For each included depression instrument, a standardized mean difference (SMD; we used Hedges’ *g* in view of the small sample sizes of many of the included trials) was calculated to estimate the difference between arms within a trial at post-treatment. Effect sizes were calculated using post-treatment means, standard deviations, and number of participants in each group. If these were not available, we extracted change scores or binary data, or ultimately we attempted to convert other available statistics into Hedges’ *g* (e.g., F-value, t-value). We prioritized analyses employing appropriate imputation for missing data (e.g., mixed models), followed by less justifiable imputation (e.g., last observation carried forward), and then completer analyses.

### Statistical Analyses

The current study had five objectives: 1) to examine the inter-rater reliability of Metapsy RoB ratings in a subsample of 237 trials; 2) to provide an overview of the prevalence of different types of biases in the field; 3) to examine overall RoB and domain-specific changes over time; 4) to test the association between RoB and trial characteristics; and 5) to test the association between RoB and effect size estimates. The study-level RoB score was used for objectives 2 to 4, and the instrument-level RoB score for objective 5, pairing each effect size with the RoB rating of its corresponding instrument. Details on deviations from the preregistered analysis plan (Miguel et al., 2026) are provided in Supplement F.

Inter-rater reliability (objective 1) was estimated separately for each of the ten rater pairs, at two levels. First, one overall Cohen’s kappa was computed at the item level, on a binarized version of the ratings (Yes/Probably Yes vs. No/Probably No or No Information), as implemented in the checkRobDiscrepancies function from the metapsyTools package (Harrer et al., 2022), including only cells where both raters provided a valid rating. As a sensitivity analysis, kappa was recomputed restricting to the 16 manually rated items only, excluding seven items pre-filled automatically from the Metapsy database (items 3, 4, 10, 12, 13, 16, and 17), which are identical by construction for both raters and contribute to observed agreement independently of genuine independent judgment, as well as the one free-text item (item 20, extraction of trial registration number). Second, Gwet’s Agreement Coefficient 2 (AC2) with ordinal weights was computed at the score level, comparing the final RoB scores (low risk, some concerns, high risk) assigned by each rater independently, using the irrCAC package (Gwet, 2026), yielding six AC2 values: one for each of the five domains and one for the overall RoB score. Ordinal weights were used to account for the severity of disagreement, penalizing discrepancies between distant categories (e.g., low risk vs. high risk) more heavily than adjacent ones (e.g., low risk vs. some concerns). Observed percentage agreement was also reported for each domain. All values were averaged across the ten rater pairs. Different statistics were used at the two levels because the score-level distributions were highly skewed, with near-zero variance in some domains, which renders kappa-family coefficients unstable (see Supplement E and F).

To provide an overview of the prevalence of bias (objective 2), we calculated the number and percentage of trials rated as low risk, some concerns, or high risk of bias for each domain and for the overall RoB rating, relative to the total number of included trials.

To examine trends in RoB over time (objective 3), we fitted Poisson regression models with the annual count of trials in each RoB category as the dependent variable. Publication year was entered as a predictor alongside RoB category, and an interaction term between year and RoB category tested whether growth rates differed across categories. Because the total number of published trials grows over time, this interaction effectively tests whether the proportion of trials in each RoB category changes over time, rather than just whether absolute counts increase. Model-based predicted counts were plotted against publication year for each RoB category. Analyses were conducted for both overall RoB and separately for each domain.

To identify trial characteristics associated with RoB (objective 4), we examined the relationship between RoB category and control group type (waitlist vs. CAU and other control conditions), psychotherapy type (CBT vs. other), delivery format (individual vs. group vs. guided self-help vs. other), inclusion criterion for depression (clinical diagnosis vs. elevated symptoms based on a cut-off score), income level of the country where the trial was conducted (low/middle vs. high income, according to the World Bank country income classification matched to the publication year of each trial), and sample size (continuous, log-transformed), using proportional odds logistic regression models for both overall RoB and each domain separately.

All predictors were entered simultaneously. The proportional odds model estimates the odds of being rated in a higher risk of bias category, assuming this odds ratio is constant across thresholds. Results are reported as odds ratios (OR) with 95% confidence intervals. Studies with missing data on any predictor were excluded (*n* = 2, due to unclassifiable income in two multinational trials).

To examine the association between RoB and effect size estimates (objective 5), the instrument-level RoB score was used, pairing each effect size with its corresponding RoB rating at the instrument level. Effect sizes were pooled using a correlated and hierarchical effects (CHE) model (Pustejovsky & Tipton, 2022), with random effects at the study level and within-study level to account for the clustering of multiple effect sizes within studies. The within-study correlation between effect sizes from the same study was assumed to be *ρ* = 0.6 (Pustejovsky & Tipton, 2022). RoB category was included as a fixed stratification term, allowing estimation of pooled effect sizes separately for each RoB category. Confidence and prediction intervals for these pooled estimates come directly from the fitted model. Pairwise contrasts were computed to compare effect sizes across RoB categories, using cluster-robust variance estimation with the CR2 small-sample correction (Pustejovsky & Tipton, 2018). Moderation tests used model-based inference from the fitted CHE models. Analyses were conducted for overall RoB (instrument-level) and each domain separately, both unadjusted and adjusted for control group type (waitlist vs. non-waitlist), which was included as a covariate in the adjusted models; pooled effect sizes from the adjusted models refer to trials with non-waitlist control groups. Differences in effect sizes (Δ*g*, defined as the difference in pooled effect sizes between RoB categories) were interpreted in light of their magnitude and the uncertainty reflected in the 95% confidence intervals, without applying a fixed threshold. Heterogeneity was assessed through τ² and prediction intervals.

As an exploratory extension of objective 5, we examined whether Δ*g* varied systematically according to study characteristics (those explored in objective 4). For this purpose, RoB was dichotomized into low risk versus higher risk, combining some concerns and high risk into a single category, as maintaining three separate categories would result in subgroups too small for stable model estimation. Moderation was tested by adding interaction terms between RoB group (low risk vs. higher risk) and each predictor in meta-regression models based on the CHE framework.

As a sensitivity analysis, all meta-epidemiological analyses (objectives 2, 4, and 5) were repeated restricting the sample to trials published from 2001 onwards, the year the revised CONSORT statement was published (Moher et al., 2001).

All analyses were conducted using R version 4.5.3, with packages metapsyTools (Harrer et al., 2022), irrCAC (Gwet, 2026), MASS (Venables & Ripley, 2002), metafor (Viechtbauer, 2010), and clubSandwich (Pustejovsky, 2026).

## Results

### Study Inclusion and Characteristics

The PRISMA flowchart describing the inclusion process and the references of included studies are reported in Supplement G. After screening 29,508 references and 5,208 full-texts, 514 RCTs met inclusion criteria for this study. The 514 RCTs had 572 comparisons between psychotherapy and control conditions, and included 66,127 participants.

Most studies examined cognitive behavioral therapies (*n* = 261, 50.8%), delivered face-to-face individually (*n* = 165, 32.1%) or in groups (*n* = 146, 28.4%), and compared psychotherapy to care-as-usual (*n* = 239, 46.5%) or waitlist (*n* = 192, 37.4%) control conditions. Participants were mostly middle-aged adults (mean age 43.3 years, 72.7% women) with elevated depressive symptoms based on a cut-off score (*n* = 284, 55.3%) or a clinical diagnosis (*n* = 230, 44.7%), recruited from the community (*n* = 208, 40.5%). Further description of the characteristics of included studies is provided in Supplement H.

### Objective 1: Inter-Rater Reliability

Inter-rater reliability estimates are reported in Table 2 and Supplement E, based on 10 rater pairs and 237 studies. At the item level, mean Cohen’s kappa across the ten rater pairs was 0.86 (range: 0.73–0.93), indicating strong agreement on individual item ratings. The sensitivity analysis restricting to the 16 manually rated items only yielded a mean kappa of 0.77 (range: 0.55–0.90), reflecting a more conservative estimate of independent agreement. At the score level, mean Gwet’s AC2 across the five domains ranged from 0.68 to 0.93, and was 0.78 for the overall RoB score (range across pairs: 0.60–0.89). Observed percentage agreement ranged from 77.8% to 94.7% across domains, and was 72.3% for the overall RoB score. Agreement was highest for Domain 4 (measurement of the outcome; AC2 = 0.93, 94.7% observed agreement) and Domains 1 and 5 (randomization process, selective outcome reporting; both AC2 = 0.88; 84.4% and 85.1% observed agreement), and lowest for Domain 3 (missing outcome data; AC2 = 0.68, 77.8% observed agreement) and Domain 2 (deviations from intended interventions; AC2 = 0.73, 80.0% observed agreement).

**Table 2.**
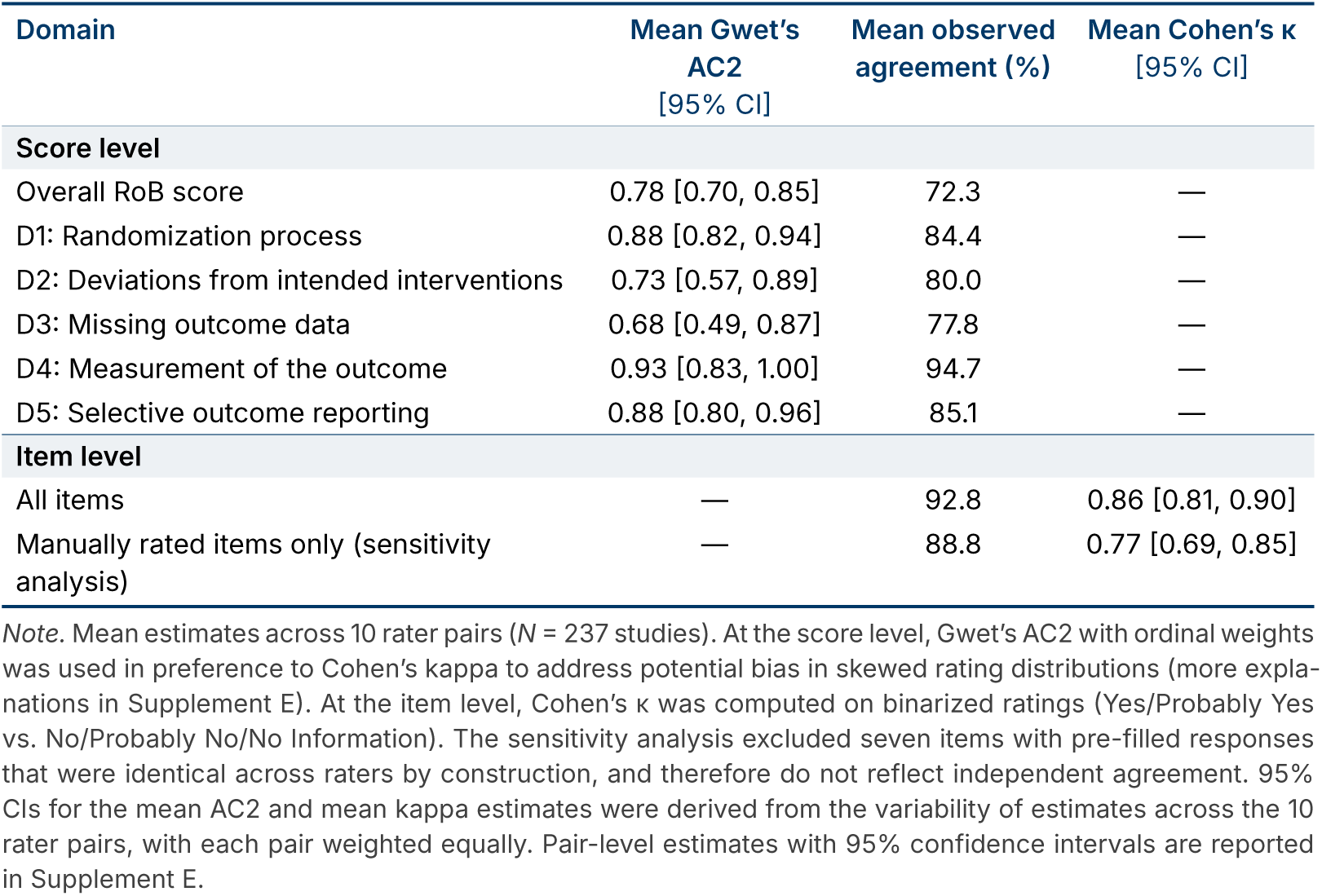
Inter-rater reliability of RoB ratings: score-level agreement (Gwet’s AC2) and item-level agreement (Cohen’s κ)

| **Domain** | **Mean Gwet’s AC2**<br>[95% CI] | **Mean observed agreement (%)** | **Mean Cohen’s κ**<br>[95% CI] |
|---|---|---|---|
| **Score level** |  |  |  |
| Overall RoB score | 0.78 [0.70, 0.85] | 72.3 | — |
| D1: Randomization process | 0.88 [0.82, 0.94] | 84.4 | — |
| D2: Deviations from intended interventions | 0.73 [0.57, 0.89] | 80.0 | — |
| D3: Missing outcome data | 0.68 [0.49, 0.87] | 77.8 | — |
| D4: Measurement of the outcome | 0.93 [0.83, 1.00] | 94.7 | — |
| D5: Selective outcome reporting | 0.88 [0.80, 0.96] | 85.1 | — |
| **Item level** |  |  |  |
| All items | — | 92.8 | 0.86 [0.81, 0.90] |
| Manually rated items only (sensitivity analysis) | — | 88.8 | 0.77 [0.69, 0.85] |

*Note*. Mean estimates across 10 rater pairs (*N* = 237 studies). At the score level, Gwet’s AC2 with ordinal weights was used in preference to Cohen’s kappa to address potential bias in skewed rating distributions (more expla-nations in Supplement E). At the item level, Cohen’s κ was computed on binarized ratings (Yes/Probably Yes vs. No/Probably No/No Information). The sensitivity analysis excluded seven items with pre-filled responses that were identical across raters by construction, and therefore do not reflect independent agreement. 95% CIs for the mean AC2 and mean kappa estimates were derived from the variability of estimates across the 10 rater pairs, with each pair weighted equally. Pair-level estimates with 95% confidence intervals are reported in Supplement E.

### Objective 2: Prevalence of Risk of Bias

The distribution of RoB ratings varied considerably across domains (Figure 1). Domain 4 (measurement of the outcome) was the domain with the largest percentage of low-risk trials (93.2%). Domain 2 (deviations from intended interventions) was also predominantly low risk (71%). Domain 5 (selective outcome reporting) was characterized by a high proportion of some concerns ratings (70%), with relatively few trials rated as low risk (25.5%) or high risk (4.5%). Domain 1 (randomization process) showed a more even distribution, with 51.2% of trials rated as low risk and 45.5% as some concerns. Domain 3 (missing outcome data) had the largest percentage of high-risk ratings (36%).

**Figure 1.**
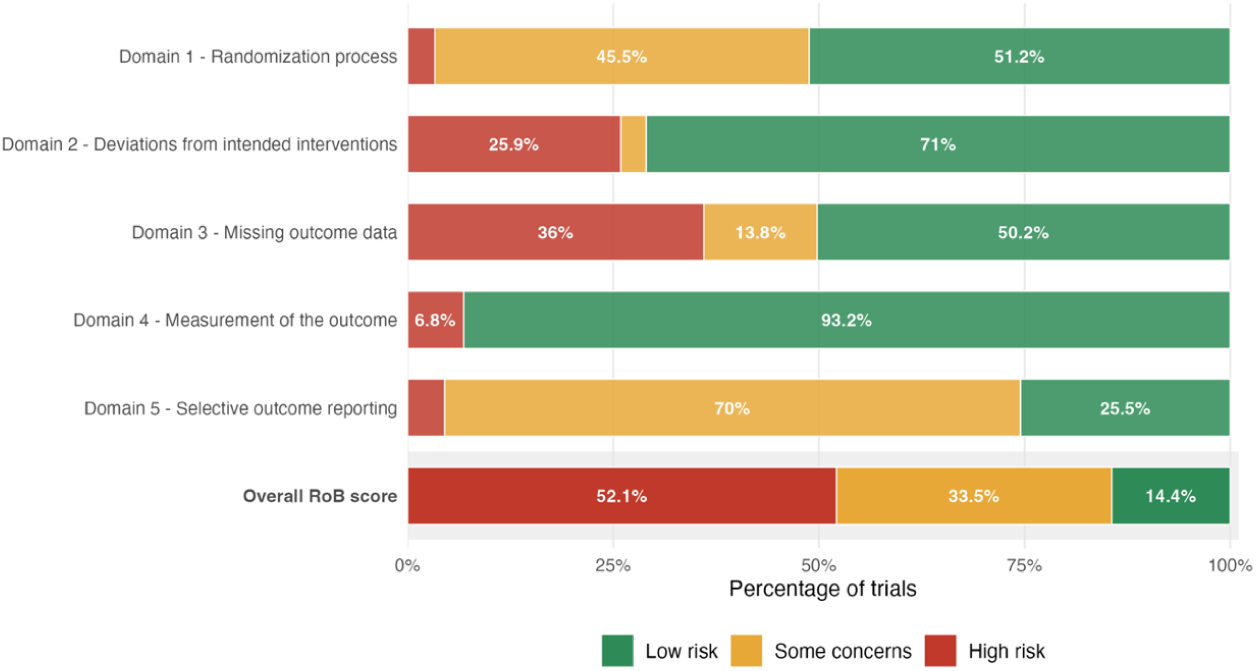
Percentage of trials rated as low risk, some concerns, or high risk of bias for each domain and the overall risk of bias score (*N* = 514).

At the overall study level, only 14.4% of trials were rated as low risk, while 33.5% received some concerns and 52.1% were rated as high risk, reflecting the cumulative impact of domain-level ratings on the overall score.

Sensitivity analyses restricted to post-2001 trials (*n* = 461) showed a consistent pattern of modestly improved RoB profiles across all domains, with the largest difference observed for Domain 2 (high risk: 21% vs. 25.9%) and overall RoB (high risk: 48.2% vs. 52.1%), suggesting some improvement in trial conduct following the publication of the revised CONSORT statement. Results are reported in Supplement I.

### Objective 3: Trends in Risk of Bias Over Time

The share of trials in each overall RoB category per 5-year period is shown in Figure 2, and Poisson regression model coefficients are reported in Supplement J. Poisson regression models included an interaction term between publication year and RoB category, with high risk as the reference category, to test whether the growth rate of each category differed from that of high-risk trials over time.

**Figure 2.**
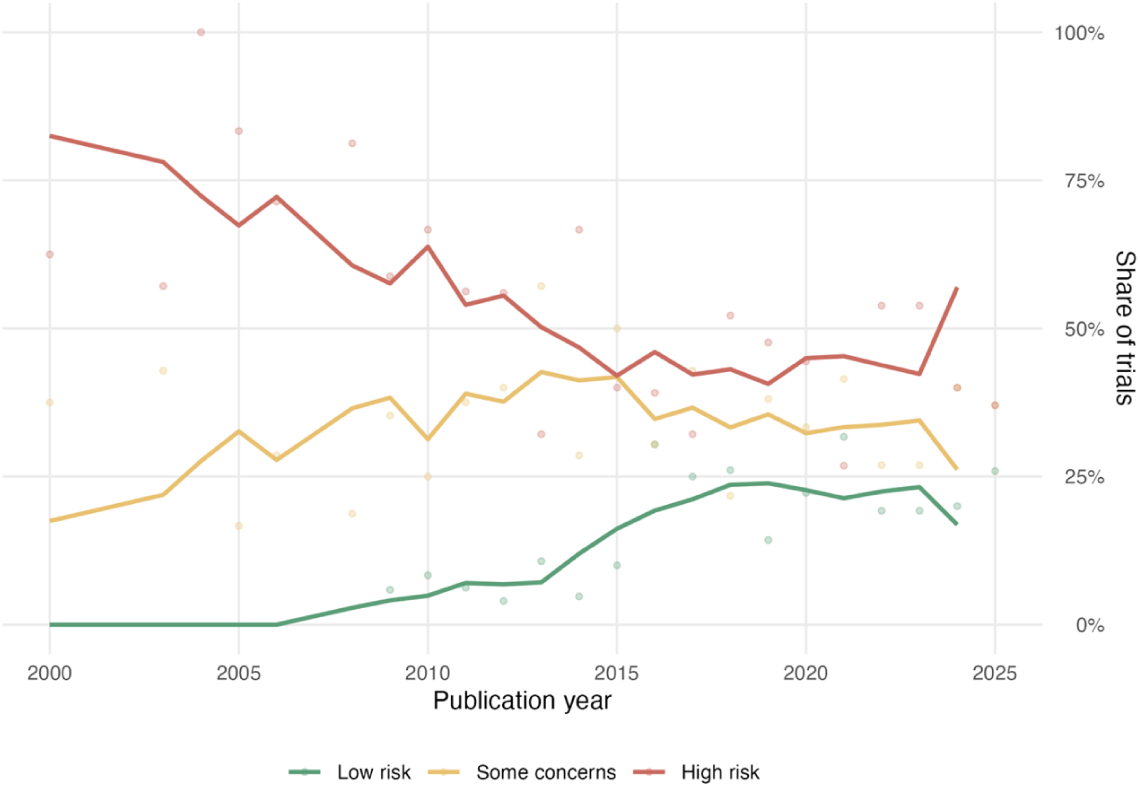
Share of trials rated as low risk, some concerns, or high risk for the overall risk of bias score, by publication year (2000–2025). Data prior to 2000 are not shown due to small annual trial counts. Lines represent 5-year rolling averages; dots show observed annual proportions. Years with fewer than five published trials are excluded due to unstable estimates. The rolling average at the beginning and end of the series incorporates fewer years and should be interpreted with caution. The proportion of high-risk trials declined substantially over the study period, while the share of low-risk trials increased from near zero after 2009. Domain-level results are shown in Supplement J.

At the overall RoB level, the share of both low risk and some concerns trials grew significantly faster than that of high-risk trials (low risk: β = 1.250, SE = 0.245, *p* <. 001; some concerns: β = 0.476, SE = 0.129, *p* <.001), indicating a declining proportion of high-risk trials over time. No low-risk trials were observed before 2009. At the domain level, patterns varied. For Domain 1, a stable distribution over time was observed. For Domains 2, 3, and 4, the share of low-risk trials grew significantly faster than that of high-risk trials (β = 0.677, *p* <.001; β = 0.768, *p* <.001; β = 0.782, *p* <.001, respectively), indicating an improving trend. For Domain 5, the proportion of some concerns ratings declined relative to high risk over time (β = −1.382, SE = 0.452, *p* =.002).

### Objective 4: Trial Characteristics Associated with Risk of Bias

Results of the ordinal regression models are reported in Table 3. Sample size was the strongest and most consistent predictor of RoB: larger trials had lower odds of worse RoB across the overall score (OR = 0.43, 95% CI [0.33, 0.55]) and all five domains (OR range: 0.42–0.56, all 95% CIs excluding 1.00). Guided self-help trials had lower odds of worse RoB compared to individually delivered trials at the overall level and for Domains 1, 3, and 5 (OR range: 0.26–0.55, all 95% CIs excluding 1.00), with the strongest effect observed for Domain 5 (OR = 0.26, 95% CI [0.15, 0.46]). Trials from low-or middle-income countries had more than twice the odds of higher RoB on Domain 2 compared to trials from high-income countries (OR = 2.40, 95% CI [1.51, 3.82]). Trials recruiting based on a cut-off score had higher odds of higher RoB in missing data ratings than trials requiring a clinical diagnosis (Domain 3; OR = 1.47, 95% CI [1.03, 2.11]). No other predictors were significantly associated with RoB.

**Table 3.**
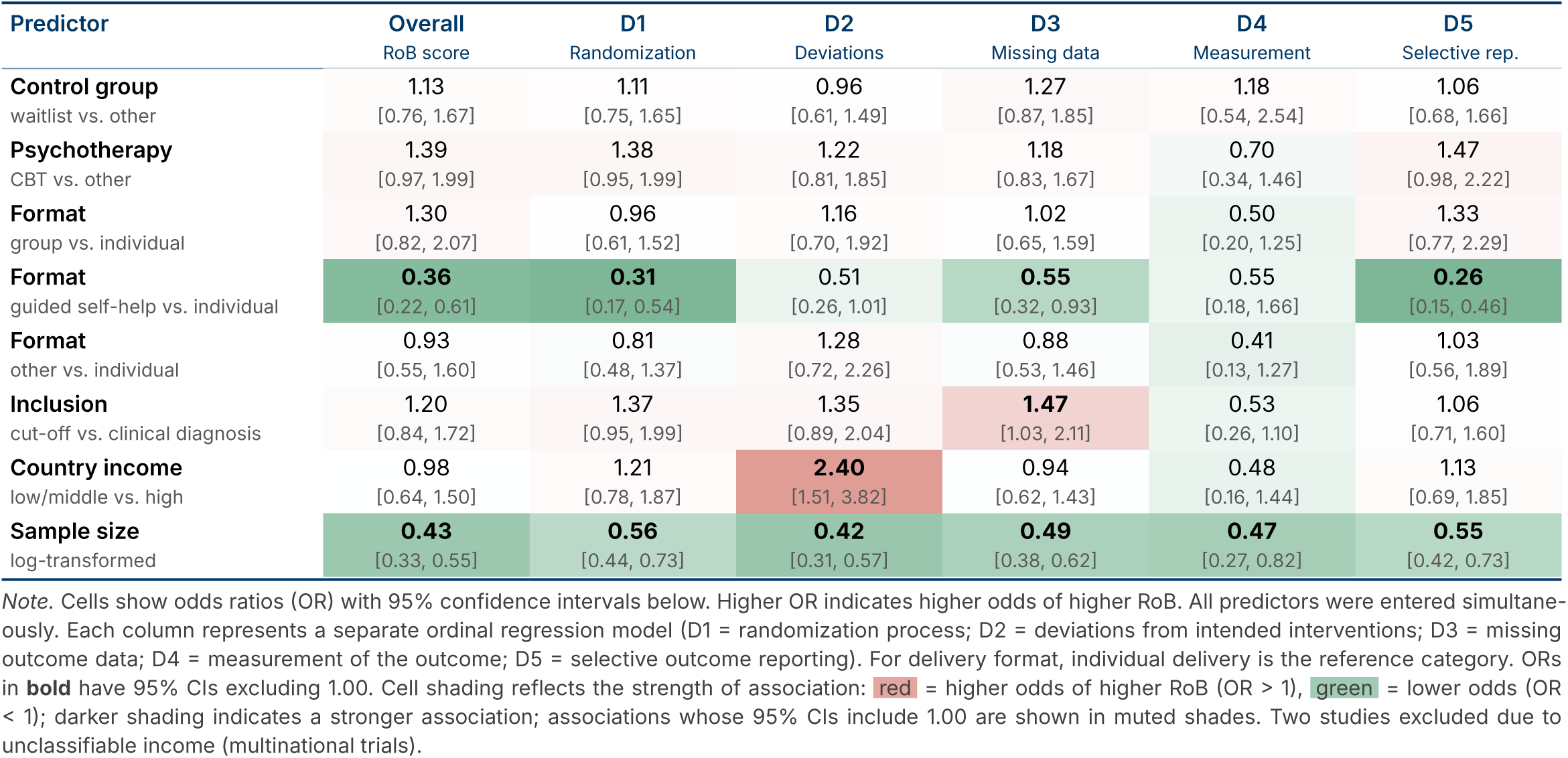
Ordinal regression results for trial characteristics associated with risk of bias (n = 512)

| Predictor | Overall | D1 | D2 | D3 | D4 | D5 |
|---|---|---|---|---|---|---|
|  | RoB score | Randomization | Deviations | Missing data | Measurement | Selective rep. |
| **Control group**<br>waitlist vs. other | 1.13<br>[0.76, 1.67] | 1.11<br>[0.75, 1.65] | 0.96<br>[0.61, 1.49] | 1.27<br>[0.87, 1.85] | 1.18<br>[0.54, 2.54] | 1.06<br>[0.68, 1.66] |
| **Psychotherapy**<br>CBT vs. other | 1.39<br>[0.97, 1.99] | 1.38<br>[0.95, 1.99] | 1.22<br>[0.81, 1.85] | 1.18<br>[0.83, 1.67] | 0.70<br>[0.34, 1.46] | 1.47<br>[0.98, 2.22] |
| **Format**<br>group vs. individual | 1.30<br>[0.82, 2.07] | 0.96<br>[0.61, 1.52] | 1.16<br>[0.70, 1.92] | 1.02<br>[0.65, 1.59] | 0.50<br>[0.20, 1.25] | 1.33<br>[0.77, 2.29] |
| **Format**<br>guided self-help vs. individual | **0.36**<br>[0.22, 0.61] | **0.31**<br>[0.17, 0.54] | 0.51<br>[0.26, 1.01] | **0.55**<br>[0.32, 0.93] | 0.55<br>[0.18, 1.66] | **0.26**<br>[0.15, 0.46] |
| **Format**<br>other vs. individual | 0.93<br>[0.55, 1.60] | 0.81<br>[0.48, 1.37] | 1.28<br>[0.72, 2.26] | 0.88<br>[0.53, 1.46] | 0.41<br>[0.13, 1.27] | 1.03<br>[0.56, 1.89] |
| **Inclusion**<br>cut-off vs. clinical diagnosis | 1.20<br>[0.84, 1.72] | 1.37<br>[0.95, 1.99] | 1.35<br>[0.89, 2.04] | **1.47**<br>[1.03, 2.11] | 0.53<br>[0.26, 1.10] | 1.06<br>[0.71, 1.60] |
| **Country income**<br>low/middle vs. high | 0.98<br>[0.64, 1.50] | 1.21<br>[0.78, 1.87] | **2.40**<br>[1.51, 3.82] | 0.94<br>[0.62, 1.43] | 0.48<br>[0.16, 1.44] | 1.13<br>[0.69, 1.85] |
| **Sample size**<br>log-transformed | **0.43**<br>[0.33, 0.55] | **0.56**<br>[0.44, 0.73] | **0.42**<br>[0.31, 0.57] | **0.49**<br>[0.38, 0.62] | **0.47**<br>[0.27, 0.82] | **0.55**<br>[0.42, 0.73] |

*Note.* Cells show odds ratios (OR) with 95% confidence intervals below. Higher OR indicates higher odds of higher RoB. All predictors were entered simultane-ously. Each column represents a separate ordinal regression model (D1 = randomization process; D2 = deviations from intended interventions; D3 = missing outcome data; D4 = measurement of the outcome; D5 = selective outcome reporting). For delivery format, individual delivery is the reference category. ORs in **bold** have 95% CIs excluding 1.00. Cell shading reflects the strength of association: red = higher odds of higher RoB (OR > 1), green = lower odds (OR < 1); darker shading indicates a stronger association; associations whose 95% CIs include 1.00 are shown in muted shades. Two studies excluded due to unclassifiable income (multinational trials).

Sensitivity analyses restricted to trials published from 2001 onwards (*n* = 459) showed consistent results, with the association between symptom-based inclusion criteria and Domain 3 somewhat stronger (OR = 1.61, 95% CI [1.10, 2.36]) and the income effect on Domain 2 more pronounced (OR = 3.50, 95% CI [2.16, 5.76]; Supplement I).

### Objective 5: Association Between Risk of Bias and Effect Size Estimates

Pooled effect sizes per RoB category across all domains are displayed in Figure 3. The main results of the CHE models are reported in Table 4, with full details in Supplement K. At the overall level, pooled effect sizes differed across RoB categories, with low-risk trials showing the smallest effects (*g* = 0.48, 95% CI [0.34, 0.62]), followed by some concerns (*g* = 0.68, 95% CI [0.59, 0.77]), and high-risk trials showing the largest pooled effect sizes (*g* = 0.82, 95% CI [0.74, 0.89]). Prediction intervals were wide and similar in width across categories (low risk: −0.72 to 1.67; some concerns: −0.51 to 1.87; high risk: −0.37 to 2.01), reflecting substantial heterogeneity (I² = 77.1%). All three pairwise contrasts showed a difference between categories (low risk vs. some concerns: Δ*g* = −0.20, 95% CI [−0.31, −0.09]; low risk vs. high risk: Δ*g* = −0.34, 95% CI [−0.45, −0.23]; some concerns vs. high risk: Δ*g* = −0.13, 95% CI [−0.25, −0.02]). Adjusting for control group type had minimal impact on these estimates (Supplement K).

**Figure 3.**
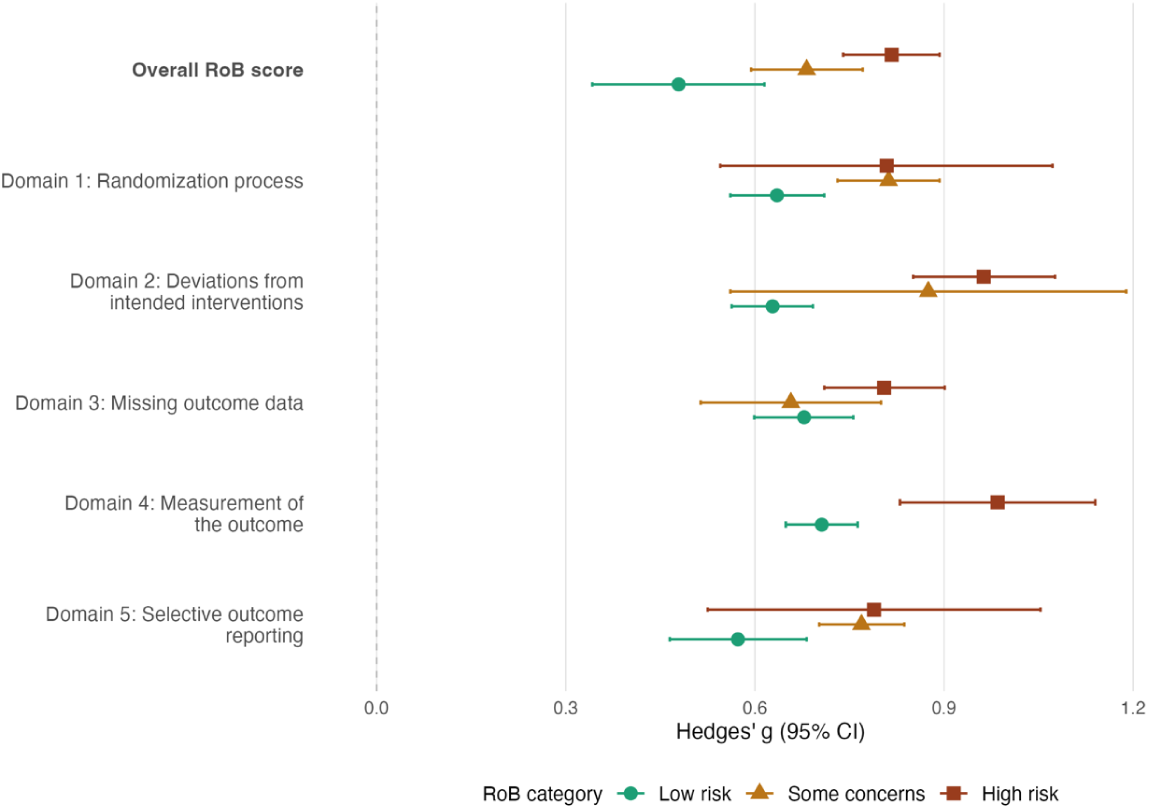
Pooled effect sizes (psychotherapies vs. control conditions) per risk of bias category across the overall risk of bias score and five domains (*N* = 514 trials, 941 effect sizes). Points represent pooled Hedges’ *g* estimated from correlated and hierarchical effects (CHE) models; horizontal lines represent 95% confidence intervals. Note that overlapping confidence intervals do not necessarily indicate non-significant differences; pairwise contrasts are reported in Table 4.

**Table 4.**
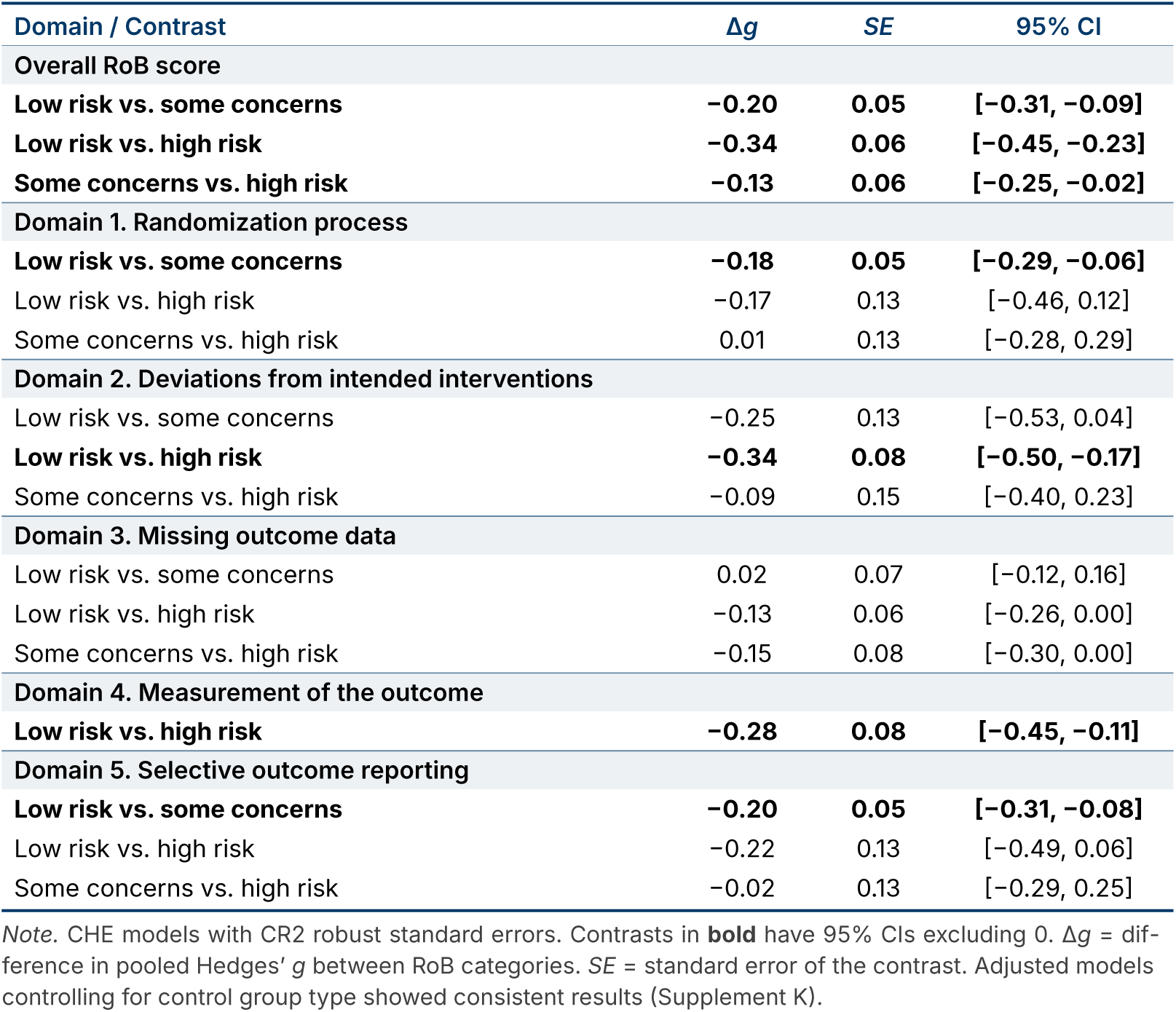
Pairwise contrasts in effect sizes across risk of bias categories (N = 514 trials, 941 effect sizes)

| Domain / Contrast | Δ*g* | *SE* | 95% CI |
|---|---|---|---|
| Overall RoB score |  |  |  |
| **Low risk vs. some concerns** | **−0.20** | **0.05** | **[−0.31, −0.09]** |
| **Low risk vs. high risk** | **−0.34** | **0.06** | **[−0.45, −0.23]** |
| **Some concerns vs. high risk** | **−0.13** | **0.06** | **[−0.25, −0.02]** |
| Domain 1. Randomization process |  |  |  |
| **Low risk vs. some concerns** | **−0.18** | **0.05** | **[−0.29, −0.06]** |
| Low risk vs. high risk | −0.17 | 0.13 | [−0.46, 0.12] |
| Some concerns vs. high risk | 0.01 | 0.13 | [−0.28, 0.29] |
| Domain 2. Deviations from intended interventions |  |  |  |
| Low risk vs. some concerns | −0.25 | 0.13 | [−0.53, 0.04] |
| **Low risk vs. high risk** | **−0.34** | **0.08** | **[−0.50, −0.17]** |
| Some concerns vs. high risk | −0.09 | 0.15 | [−0.40, 0.23] |
| Domain 3. Missing outcome data |  |  |  |
| Low risk vs. some concerns | 0.02 | 0.07 | [−0.12, 0.16] |
| Low risk vs. high risk | −0.13 | 0.06 | [−0.26, 0.00] |
| Some concerns vs. high risk | −0.15 | 0.08 | [−0.30, 0.00] |
| Domain 4. Measurement of the outcome |  |  |  |
| **Low risk vs. high risk** | **−0.28** | **0.08** | **[−0.45, −0.11]** |
| Domain 5. Selective outcome reporting |  |  |  |
| **Low risk vs. some concerns** | **−0.20** | **0.05** | **[−0.31, −0.08]** |
| Low risk vs. high risk | −0.22 | 0.13 | [−0.49, 0.06] |
| Some concerns vs. high risk | −0.02 | 0.13 | [−0.29, 0.25] |

*Note*. CHE models with CR2 robust standard errors. Contrasts in **bold** have 95% CIs excluding 0. Δ*g* = difference in pooled Hedges' *g* between RoB categories. *SE* = standard error of the contrast. Adjusted models controlling for control group type showed consistent results (Supplement K).

At the domain level, Domain 1 showed a difference between low risk and some concerns trials (Δ*g* = −0.18, 95% CI [−0.29, −0.06]); no clear difference was found between low risk and high-risk trials (95% CI [−0.46, 0.12]). For Domain 2, low-risk trials differed from high-risk trials (Δ*g* = −0.34, 95% CI [−0.50, −0.17]); no clear difference was found between low risk and some concerns trials (95% CI [−0.53, 0.04]). Domain 3 showed no clear differences across RoB categories. Domain 4 showed a difference between low-risk and high-risk trials (Δ*g* = −0.28, 95% CI [−0.45, −0.11]). For Domain 5, low-risk trials differed from some concerns trials (Δ*g* = −0.20, 95% CI [−0.31, −0.08]); no clear difference was found between low risk and high-risk trials (Δ*g* = −0.22, 95% CI [−0.49, 0.06]). Adjusted domain-level results were very similar to unadjusted results and are reported in full in Supplement K.

In the exploratory moderation analysis, country income level was the only moderator associated with the RoB–effect size association: this was more pronounced in low-and middle-income countries than in high-income countries (interaction estimate = 0.54, 95% CI [0.16, 0.93]). No other study characteristics moderated the association between RoB and effect size.

Sensitivity analyses restricted to trials published from 2001 onwards (*n* = 461) showed consistent results (Supplement I).

## Discussion

This study examined risk of bias in a complete cohort of 514 randomized controlled trials on psychotherapy compared with control conditions for adult depression, covering its prevalence, trends over time, association with trial characteristics, and association with effect size estimates. Risk of bias was widespread, with only 14.4% of trials rated as overall low risk and 52.1% as high risk. The temporal analyses showed that the proportion of high-risk trials has declined over time, while low-risk trials have increased. Higher RoB was systematically associated with larger effect sizes, with high-risk trials showing pooled effects approximately 70% larger than low-risk trials. This difference is large enough to have meaningful implications for how psychotherapy evidence is interpreted and used in clinical guidelines.

### Prevalence, Trends, and Correlates of Risk of Bias in Psychotherapy Trials for Depression

The overall prevalence figures are considerably more concerning than prior estimates from this field. In earlier work using the same database but applying only four items of the Cochrane RoB 1 tool, only 23% of trials could be considered at low risk of bias (Cuijpers et al., 2019). Our current estimate of 14.4% low risk is lower than this prior figure, which likely reflects the more comprehensive and stricter criteria used in the current study, since rating more domains makes an overall low-risk rating harder to achieve, though the two estimates are not directly comparable given the differences in tools and analytic approach. The prevalence of RoB varied across domains. Domain 4 (measurement of the outcome) showed the most favorable profile, with 93% of trials rated as low risk. This pattern reflects a deliberate rating decision in the tool, in which self-report instruments, the most frequently used measures in depression psychotherapy (Sprenger et al., 2026), are rated as low risk for measurement bias even when administered to unblinded participants. This decision was based on empirical evidence showing that self-reports produce effect estimates comparable to those of masked clinician ratings, while unmasked clinician-administered assessments produce substantially larger effect estimates, suggesting that the source of measurement bias lies in the lack of assessor masking rather than in the use of self-reports (Cuijpers, Li, et al., 2010; Miguel et al., 2025). Domain 3 (missing outcome data) showed the highest prevalence of high-risk ratings (36%), reflecting how common and often poorly handled attrition is in this literature. Domain 5 (selective outcome reporting) was mostly rated as some concerns (70%) due to the widespread absence of a prospective trial registration and/or unclear specification of outcomes in prospective registrations.

Consistent with the Poisson regression results, the proportion of high-risk trials declined over time while low-risk trials increased from 2009 onwards, in line with the gradual adherence to CONSORT reporting standards (Moher et al., 2001). Since ratings are necessarily based on what is reported in publications, it is difficult to disentangle whether improvements in ratings over time reflect genuine improvements in trial conduct or rather better reporting of procedures. Domain 5 shows this most clearly: as registration increased following the ICMJE mandate in 2005 (De Angelis et al., 2004), discrepancies between planned and published outcomes became detectable, reclassifying trials from some concerns into high risk rather than reflecting a true increase in selective reporting. Prospective registration rates in psychotherapy research have been typically low, with previous estimates suggesting that only 24-40% of trials are prospectively registered (Bradley et al., 2017; Miguel et al., 2021; Stoll et al., 2020), though the improvement over time observed in our data suggests the situation may be gradually changing.

Several trial characteristics were associated with RoB. Sample size was the strongest and most consistent predictor of RoB, associated with lower odds of worse ratings for the overall score and all five domains, consistent with broader meta-epidemiological evidence that smaller trials produce systematically larger effect size estimates (Dechartres et al., 2013). Underpowered trials thus may not only be imprecise and contribute to effect heterogeneity (Kuper et al., 2025) but are systematically more likely to be biased. Trials examining guided self-help interventions were associated with lower risk of bias in several domains. This possibly reflects the combination of standardized delivery platforms, pre-specified protocols, and the fact that these trials have been conducted more recently, with higher awareness about registration and reporting standards. Trials from low-and middle-income countries had more than twice the odds of worse ratings on Domain 2 (deviations from intended interventions), likely reflecting structural differences in trial infrastructure such as less standardized and monitored psychotherapy and control conditions. These findings are consistent with previous evidence that psychotherapy trials conducted in countries with limited research infrastructure tend to show higher rates of methodological weaknesses (Tong et al., 2023), for example a lower likelihood of conducting intention-to-treat analyses. Trials recruiting participants based on a symptom cut-off score showed higher odds of worse missing data ratings than trials requiring a formal clinical diagnosis, though this association was not consistent across other domains and its interpretation remains unclear.

### Association Between Risk of Bias and Effect Size Estimates

Effect sizes and RoB were strongly associated, with effects increasing steadily from low to high-risk trials. High-risk trials yielded pooled effects approximately 70% larger than low-risk trials (*g* = 0.82 vs. *g* = 0.48). This pattern held across all three pairwise comparisons between risk categories: low risk versus some concerns (Δ*g* = −0.20, *p* <.001), low risk versus high risk (Δ*g* = −0.34, *p* <.001), and some concerns versus high risk (Δ*g* = −0.13, *p* =.021), and remained significant after adjusting for control group type (waitlist vs other controls), suggesting that the observed effect size inflation may not be attributable to the known overestimation associated with waitlist comparisons (Furukawa et al., 2014) but may represent independent bias. The association between RoB and effect size was moderated by country income level, being significantly stronger in trials from low-and middle-income countries. These findings are broadly consistent with prior work in psychotherapy trials, where methodological weaknesses have been associated with larger effect size estimates, though estimates have not always been consistent across studies and designs (Barth et al., 2013; Cuijpers et al., 2019; Cuijpers, Li, et al., 2010; Cuijpers, van Straten, et al., 2010; Munder & Barth, 2018). Using the same database but applying only four items of the Cochrane RoB 1 tool, Cuijpers et al. (2019) found an unadjusted pooled effect size of *g* = 0.70 that dropped to *g* = 0.31 after adjusting for risk of bias, publication bias, and type of control group.

At the domain level, the patterns observed here are generally consistent with some of the wider meta-epidemiological literature, though direct comparisons are complicated by important differences in how some bias domains are operationalized. For Domain 1, the association between higher RoB and larger effect sizes aligns with evidence that inadequate randomization and allocation concealment are associated with overestimation of treatment effects (Savović et al., 2018; Savović et al., 2012; Wang et al., 2024; Wood et al., 2008), though not all pairwise contrasts were significant within each domain, partly reflecting the uneven distribution of trials across risk categories. The absence of a significant effect size association for Domain 3 is somewhat unexpected given its high prevalence of high-risk ratings. One explanation relates to a practical limitation in how Domain 3 could be assessed. When a trial described appropriate methods for handling missing data, we rated it as low risk even when it was not possible to verify from the results tables whether the reported data were truly based on those methods. This means that some trials rated as low risk may have contributed effect sizes based on analyses that did not fully implement what was described, which would attenuate the association between Domain 3 ratings and effect size estimates. A further possibility is that attrition bias operates in opposite directions depending on whether dropout occurs from the treatment or the control arm: severely distressed patients dropping out of waitlist conditions may leave a less distressed comparison group, leading to underestimation of effects, while patients dropping out of active treatment due to its demands may leave a more treatment-tolerant group, leading to overestimation (Munder & Barth, 2018). These opposing mechanisms may cancel out in aggregate, consistent with the wider meta-epidemiological literature, which has shown mixed findings for the association between incomplete outcome data and effect estimates (Savović et al., 2018; Wang et al., 2024). Domain 3 also showed the lowest interrater agreement of all domains, which may further attenuate its predictive validity.

For Domain 4, direct comparison is complicated by how outcomes are classified. Previous studies define ‘subjective outcomes’ as a category that merges patientrated instruments such as the BDI together with clinician-administered scales such as the Hamilton Depression Rating Scale, without distinguishing between them (Moustgaard et al., 2020; Savović et al., 2018; Savović et al., 2012; Wood et al., 2008). In our tool, self-report instruments are mainly rated as low risk based on evidence that they do not overestimate psychotherapy effects relative to masked clinician ratings (Miguel et al., 2025), while clinician-administered scales may be rated as high risk when assessors are unmasked. This distinction means our comparison is more precise than the broader literature, and null findings in previous studies such as Moustgaard et al. (2020) may partly reflect the dilution of any blinding effect by including self-reports and clinician-rated instruments within the same category. The significant contrast we observe for Domain 4 is consistent with evidence from a more precise within-trial design comparing self-report and clinician-rated outcomes within the same trials (Miguel et al., 2025), and extends this evidence to a larger and more representative cohort of psychotherapy trials for depression. Regarding Domain 5, previous meta-epidemiological studies across fields deliberately excluded selective reporting from their analyses on the grounds that it concerns nonreporting rather than bias in available results (Savović et al., 2018; Wang et al., 2024). However, there is evidence from previous studies that selective outcome reporting meaningfully distorts effect size magnitude. Statistically significant outcomes are more than twice as likely to be fully reported as non-significant ones (Chan et al., 2004), adjustment for suspected outcome reporting bias reduces treatment effect estimates by 20% or more in nearly a quarter of Cochrane reviews (Kirkham et al., 2010), and discrepancies between registries and publications are associated with substantially larger effect sizes in psychotherapy trials specifically (Miguel et al., 2021). Our Domain 5 finding is consistent with this evidence. Notably, the significant contrast was observed between low risk and some concerns trials rather than between low and high-risk trials, which is consistent with the rating structure described earlier, as some concerns captures most trials in this literature.

### Reliability and Validity of Metapsy RoB

The most widely used tool for RoB assessment in RCTs has proven difficult to apply reliably in practice, with independent evaluations reporting low inter-rater agreement, high complexity, and poor adherence to guidance (Crocker et al., 2023; Minozzi et al., 2020, 2022). In contrast, the present study provides initial empirical support for the reliability and validity of Metapsy RoB. Reliable RoB assessment depends not only on the conceptual soundness of an instrument but also on whether it can be applied consistently across reviewers. The tool demonstrated good inter-rater reliability across all domains, suggesting that its structured guidance and explicit decision rules can be applied reproducibly in psychotherapy trials. Furthermore, the systematic association between higher RoB ratings and larger effect sizes across multiple domains provides indirect evidence for the construct validity of the tool, suggesting that its ratings reflect meaningful methodological differences rather than arbitrary variation in judgment. Direct comparison with reliability estimates from other RoB tools should be interpreted cautiously, as differences may reflect not only tool design but also differences in clinical area, rater training, and study populations assessed. It is also worth noting that Cochrane’s RoB 2 was deliberately designed to allow individualized judgment tailored to each trial, while Metapsy RoB prioritizes consistency by introducing explicit interpretive rules and fixed thresholds for several items, reducing the room for idiosyncratic trial-specific judgment. This approach may support inter-rater agreement, though it inevitably involves some loss of nuance compared to fully individualized assessment. Future research should directly compare inter-rater reliability between Metapsy RoB and other RoB tools, such as RoB 2, applied to the same trials.

### Limitations and Strengths

These findings should be interpreted considering some limitations. All risk of bias ratings were based on what was reported in trial publications rather than on actual procedures, which is an inherent limitation of all RoB tools, and some trials may have implemented correct procedures that were not reported. In line with this, our RoB ratings were primarily based on the primary trial report. Published trial protocols were not systematically reviewed for additional information due to feasibility constraints. This means that relevant information for certain domains, particularly deviations from intended interventions and selective outcome reporting, may not have been captured.

Inter-rater agreement was more modest for Domains 2 and 3, which may introduce measurement error in analyses for those domains. Seven items were pre-filled automatically rather than independently rated, which is a deliberate design feature to improve standardization, but implies that the reliability estimates reported here likely represent an upper bound on agreement in contexts where pre-filling is not available (excluding pre-filled items reduced mean kappa from 0.86 to 0.77).

The association between RoB and effect size is observational, and unmeasured confounders cannot be excluded. This study focused on the specific set of methodological risks captured by the most common RoB frameworks. Other factors that may lead to overestimation of treatment benefit or an incomplete picture of treatment effects, including weak control groups, researcher allegiance, impaired treatment integrity, lack of assessment of adverse events, and the inherent heterogeneity of interventions grouped under the umbrella of psychotherapy, fall outside the scope of RoB assessment but are equally relevant for a comprehensive evaluation of the evidence base (Coyne & Kok, 2014; Cuijpers & Cristea, 2015; Leichsenring et al., 2017; Staines & Cleland, 2007).

Another important caveat concerns comparability with research using other RoB tools. The tool used here followed standard approaches in the field, but involved adaptations in the interpretation and operationalization of certain bias domains to reflect the particular challenges of psychotherapy trials. These adaptations make direct comparisons with studies using other RoB instruments difficult, though they are precisely what makes the ratings more valid for this context. The challenge of applying generic tools to specific research areas is not unique to psychotherapy. For example, in osteoarthritis research, Bolvig et al. (2018) found that not all RoB domains were equally important, and in nutrition RCTs, Bantle et al. (2026) found that most RoB 2 domains did not systematically inflate effect estimates.

Finally, the prevalence of risk of bias, its trends over time, and its association with trial characteristics and effect sizes were examined specifically in psychotherapy trials for depression, and these findings may not generalize to other mental health conditions or intervention types.

The present study has several strengths. The findings are based on a complete and systematically maintained cohort rather than a selected sample, providing a comprehensive picture of RoB across the full literature. The tool was specifically adapted for psychotherapy trials, addressing recurring interpretive decisions that generic frameworks do not fully resolve. The availability of inter-rater reliability estimates across ten independent rater pairs suggests that the ratings are sufficiently reproducible to support the meta-epidemiological analyses conducted here. The living nature of the underlying database also means that these analyses can be updated as new trials are published, providing an ongoing picture of how methodological quality in the field evolves over time.

### Implications

Several practical changes to the design, registration, and reporting of psychotherapy trials could substantially improve both trial conduct and RoB assessment, and with it the usability and trustworthiness of the evidence. For Domain 1, reporting not only that allocation was concealed but also how it was concealed would allow more confident ratings. For Domain 2, including a dedicated section in trial protocols and reports describing concomitant interventions in both study arms, specifying what was permitted, what constituted a deviation from the protocol, and how the protocol was monitored, would directly address one of the most common sources of uncertainty. For Domain 3, reporting not only the number of participants with missing outcome data but also the reasons for missingness, ideally separately by treatment arm, would allow more meaningful assessment of the likely direction of attrition bias and its impact. For Domain 4, more detailed reporting of how self-reports were administered would be informative. Treating clinicians may inadvertently influence participants’ responses, for example when questionnaires are completed in the presence of the therapist, without necessarily being aware of this risk. Prospective trial registration has improved, but there is still considerable room for improvement in Domain 5. For example, it is very difficult to verify whether analyses were truly pre-specified, as trial registries often do not provide sufficient space for detailed statistical analysis plans and full study protocols are rarely published before unblinded outcome data become available. We recognize that some of these changes concern reporting practices that are difficult to implement within the space constraints of standard trial reports, although they could be addressed through supplementary material, while others require more fundamental changes in how trials are designed and registered.

The findings have direct implications for how psychotherapy evidence is synthesized and used. Systematic reviews and clinical guidelines that pool all trials regardless of RoB may overestimate the benefits of psychotherapy for depression, given how effect estimates differ across risk of bias categories. Effect sizes from low-risk trials should be routinely presented alongside overall pooled estimates in abstracts, conclusions, and summary tables, rather than reserved for sensitivity analyses where they may receive little attention. In practice, however, this is often difficult: many meta-analyses in this field do not include enough low-risk trials to support a reliable analysis, which itself highlights the urgency of improving methodological quality at the level of primary trials. Importantly, even in low-risk trials, the pooled effect (*g* = 0.48, 95% CI [0.34, 0.62]) indicates that psychotherapy for depression, evaluated under the highest methodological standards, still probably shows a meaningful treatment benefit.

Because clinicians and patients rely on clinical guidelines informed by systematic reviews, the reliability of those recommendations depends not only on how rigorously the underlying trials were conducted but also on how consistently risk of bias is assessed across the reviews that inform them. The explicit decision rules, structured guidance, algorithms, and digital tools developed for this project are intended to reduce that inconsistency, supporting more transparent and reproducible RoB assessments in future psychotherapy evidence syntheses and, in turn, a more reliable evidence base for clinical decision-making.

## Conclusion

This study provides a systematic and empirically grounded account of risk of bias across the full body of psychotherapy trials for depression, supported by a purpose-built assessment tool whose initial reliability and construct validity have been empirically evaluated. High risk of bias is widespread in this literature and is associated with substantial overestimation of treatment effects, highlighting the importance of accounting for risk of bias when interpreting psychotherapy evidence. Even under the most rigorous standards, however, psychotherapy for depression probably shows a meaningful treatment benefit. The findings and the tool developed for this project contribute to more consistent, transparent, and reproducible psychotherapy research, from the design and reporting of primary trials to the synthesis of evidence in systematic reviews.

## Conflicts of Interest

MS is employed in the Department of Neurodevelopmental Disorders, Nagoya City University Graduate School of Medicine, which is an endowment department supported by the City of Nagoya and has received a personal fee from Daiichi-Sankyo outside the submitted work. AT received personal fees from Shionogi and Otsuka outside the submitted work. All other authors report no conflicts of interest.

**Publication Status** Preprint – changes resulting from peer review are possible.

**Preregistration**: doi.org/10.17605/OSF.IO/3BSX8

**Open Material** Metapsy RoB handbook, rating assistant, and digital tools available at metapsy.org/rob.

## Supplementary Materials

**A**: Metapsy depression database. **B**: Rating manual. **C**: Rating sheet and digital tools. **D**: Development and rationale of Metapsy RoB. **E**: Inter-rater reliability. **F**: Deviations from preregistration. **G**: PRISMA flowchart and included studies. **H**: Study characteristics. **I**: Post-CONSORT sensitivity analyses. **J**: Trends over time. **K**: RoB and effect sizes.

## Data Availability

All data produced in the present work are contained in the manuscript.

