## Supplement for "Risk of Bias in Psychotherapy Trials for Depression A large-scale empirical examination using a structured assessment tool developed for psychotherapy research"

[1] Department of Clinical, Neuro & Developmental Psychology, Amsterdam Public Health Research Institute, Vrije Universiteit Amsterdam, Amsterdam, the Netherlands; [2] Population Health and Policy Research Unit, Center for Medical Education and Internationalization, Graduate School of Medicine, Kyoto University, Kyoto, Japan; [3] Department of Neurodevelopmental Medicine, Nagoya City University Graduate School of Medical Sciences, Nagoya, Japan; [4] International Institute for Integrative Sleep Medicine (WPI-IIIS), Tsukuba Institute for Advanced Research (TIAR), University of Tsukuba, Tsukuba, Japan; [5] Department of Health Promotion and Behavioral Sciences, Kyoto University Graduate School of Medicine/School of Public Health, Kyoto, Japan; [6] Kyoto University Office of Institutional Advancement and Communications, Kyoto, Japan; [7] CORE Data Lab, Centre for Outcomes Research and Effectiveness, Research Department of Clinical, Educational and Health Psychology, UCL, London, UK; [8] Department of Clinical Psychology, University of Amsterdam, Amsterdam, the Netherlands

**Publication Status**
Preprint – changes resulting from peer review are possible.

**Correspondence to**
Clara Miguel


**Preregistration**
doi.org/10.17605/OSF.IO/3BSX8

**Open Material**
Metapsy RoB handbook, rating assistant, rating template, and digital tools available at metapsy.org/rob.

#### Supplement A. Additional information on the Metapsy depression psychotherapy database

*Overview*

The studies included in the current study were identified through the Metapsy living database of randomized trials on psychological treatments for depression (Cuijpers et al., 2023). This database is part of the Metapsy initiative (www.metapsy.org; osf.io/825c6/), which encompasses a series of open, living meta-analytic databases of interventions for mental health problems following a unified methodology and infrastructure (Harrer et al., 2025). The database is updated every four months and covers trials from 1966 onwards, with the last update on January 1st, 2026 used for the current study.

*Search strategy*

Systematic searches are conducted every four months in four major bibliographical databases, from their inception up to the latest search (in this case, January 1st, 2026): PubMed, PsycINFO, Embase, and the Cochrane Library (the latter was last searched in Sept 2022), combining index and free-text terms indicative of depression and psychotherapies, with filters for randomized controlled trials. The full search strings are available at the project website (www.metapsy.org; docs.metapsy.org/databases), and the search strings used in PubMed are presented below. In addition, reference lists of earlier meta-analyses on psychological treatments of depression are hand-searched to identify additional eligible trials.

All identified records are routinely screened by two independent researchers. Papers that could possibly meet inclusion criteria according to at least one researcher are retrieved as full texts. The decision to include or exclude a study is made by two independent researchers, and disagreements are resolved through discussion or consultation with a third researcher when necessary.

*Subsets available in the database*

The database includes trials organized into four main comparison categories: (1) psychotherapy versus control conditions (waitlist, care as usual, attention placebo, pill placebo, and other inactive controls such as a leaflet with general psychoeducation); (2) psychotherapy versus pharmacotherapy or combined treatment; (3) psychotherapy versus another psychotherapy; and (4) other comparisons (including inpatient settings, unguided treatments, dismantling studies, and other subcategories). Moreover, the database contains two main subgroups of trials that examine these comparisons: adults (mean age of the trial equal to or above 18) and children/adolescents (mean age of the trial below 18).

For the current study, only trials from adults and psychotherapy vs control were included.

*Inclusion Criteria*

Trials are eligible for inclusion in the database if they: (1) are randomized controlled trials; (2) include participants with depression, defined as either elevated depressive symptoms based on an established cut-off score on a validated self-report or clinician-rated instrument, or a clinical diagnosis of a depressive disorder based on a structured or semi-structured interview; (3) examine the effects of a psychological treatment as defined by Norcross (1990), that is, “the informed and intentional application of clinical methods and interpersonal stances derived from established psychological principles for the purpose of assisting people to modify their behaviors, cognitions, emotions, and/or other personal characteristics in directions that the participants deem desirable"; (4) include any type of psychotherapy (e.g., cognitive behavioural therapy, interpersonal psychotherapy, behavioural activation, psychodynamic therapy) delivered in any format (individual, group, guided self-help) as long as it includes human clinical support; and (5) report post-treatment outcomes on a validated measure of depressive symptomatology. Maintenance or relapse prevention trials are excluded, as well as collaborative care.

*Data Extraction*

For each included trial, data are extracted on five categories of variables (Cuijpers et al., 2023).

- *Participant characteristics:* target group, mean age, proportion of women, type of recruitment (community, clinical, primary care, other), and depression inclusion criterion (cut-off score vs. clinical diagnosis).
- *Intervention characteristics:* type of psychotherapy, delivery format (individual, group, guided self-help, other), and number of sessions.
- *General study characteristics:* publication year, country where the trial was conducted, income level of the country according to the World Bank classification matched to the publication year of each trial, and type of control condition.
- *Risk of bias:* The standard Metapsy database procedures include assessment of four items from the Cochrane Risk of Bias tool version 1 (Higgins et al., 2011): sequence generation, allocation concealment, blinding of outcome assessors, and intention-to-treat analysis. For the current study, all included trials were additionally assessed using Metapsy RoB, a structured tool developed specifically for psychotherapy research (see Supplement B).
- *Outcome data:* For the calculation of effect sizes, a priority hierarchy is used: (1) post-treatment means, standard deviations, and sample sizes; (2) change scores with standard deviations and sample sizes; (3) binary outcomes; and (4) other available statistics (t-values, F-values, p-values) converted to standardized mean differences. When both post-treatment scores and change scores are available for the same instrument, post-treatment scores are prioritized. Hedges' g is used as the primary effect size measure, appropriate given the small sample sizes of many included trials (Cuijpers et al., 2023).

*Review procedures*

All review stages, from abstract screening to full-text assessment, data extraction, and risk of bias assessment, are completed by different pairs of independent reviewers. Discrepancies are resolved through discussion or adjudication by a third reviewer.

*Search strings Metapsy depression psychotherapy database*

*PubMed*

```
Psychotherapy [MH] OR psychotherap*[All Fields] OR cbt[All Fields] OR "behavior therapies"[All Fields] OR "behavior therapy"[All Fields]
OR "behavior therapeutic"[All Fields] OR "behavior therapeutical"[All Fields] OR "behavior therapeutics"[All Fields] OR "behavior thera-
```

peutist"[all Fields] OR "behavior therapeutists"[All Fields] OR "behavior treatment"[All Fields] OR "behavior treatments"[All Fields] OR "behaviors therapies"[All Fields] OR "behaviors therapy"[All Fields] OR "behaviors therapeutics"[All Fields] OR "behaviors therapeutic"[All Fields] OR "behaviors therapeutical"[All Fields] OR "behaviors therapeutist"[All Fields] OR "behaviors therapeutists"[All Fields] OR "behaviors treatment"[All Fields] OR "behaviors treatments"[All Fields] OR "behavioral therapies"[All Fields] OR "behavioral therapy"[All Fields] OR "behavioral therapeutics"[All Fields] OR "behavioral therapeutic"[All Fields] OR "behavioral therapeutical"[All Fields] OR "behavioral therapeutist"[All Fields] OR "behavioral therapeutists"[All Fields] OR "behavioral treatment"[All Fields] OR "behavioral treatments"[All Fields] OR "behaviour therapies"[All Fields] OR "behaviour therapy"[All Fields] OR "behaviour therapeutic"[All Fields] OR "behaviour therapeutical"[All Fields] OR "behaviour therapeutics"[All Fields] OR "behaviour therapeutist"[all Fields] OR "behaviour therapeutists"[All Fields] OR "behaviour treatment"[All Fields] OR "behaviour treatments"[All Fields] OR "behaviours therapies"[All Fields] OR "behaviours therapy"[All Fields] OR "behaviours therapeutics"[All Fields] OR "behaviours therapeutic"[All Fields] OR "behaviours therapeutical"[All Fields] OR "behaviours therapeutist"[All Fields] OR "behaviours therapeutists"[All Fields] OR "behaviours treatment"[All Fields] OR "behaviours treatments"[All Fields] OR "behavioural therapies"[All Fields] OR "behavioural therapy"[All Fields] OR "behavioural therapeutics"[All Fields] OR "behavioural therapeutic"[All Fields] OR "behavioural therapeutical"[All Fields] OR "behavioural therapeutist"[All Fields] OR "behavioural therapeutists"[All Fields] OR "behavioural treatment"[All Fields] OR "behavioural treatments"[All Fields] OR "cognition therapies"[All Fields] OR "cognition therapie"[All Fields] OR "cognition therapy"[All Fields] OR "cognition therapeutical"[All Fields] OR "cognition therapeutic"[All Fields] OR "cognition therapeutics"[All Fields] OR "cognition therapeutist"[All Fields] OR "cognition therapeutists"[All Fields] OR "cognition treatment"[All Fields] OR "cognition treatments"[All Fields] OR psychodynamic[All Fields] OR Psychoanalysis[MH] OR psychoanalysis[All Fields] OR psychoanalytic*[All Fields] OR counselling[All Fields] OR counseling[All Fields] OR Counseling[MH] OR "problem-solving"[All Fields] OR mindfulness[All Fields] OR (acceptance[All Fields] AND commitment[All Fields] ) OR "assertiveness training"[All Fields] OR "behavior activation"[All Fields] OR "behaviors activation"[All Fields] OR "behavioral activation"[All Fields] OR "cognitive therapies"[All Fields] OR "cognitive therapy"[All Fields] OR "cognitive therapeutic"[All Fields] OR "cognitive therapeutics"[All Fields] OR "cognitive therapeutical"[All Fields] OR "cognitive therapeutist"[All Fields] OR "cognitive therapeutists"[All Fields] OR "cognitive treatment"[All Fields] OR "cognitive treatments"[All Fields] OR "cognitive restructuring"[All Fields] OR (("compassion-focused"[All Fields] OR "compassion-focussed"[All Fields]) AND (therapy[SH] OR therapies[All Fields] OR therapy[All Fields] OR therape*[All Fields] OR therapis*[All Fields] OR Therapeutics [All Fields] OR treatment*[All Fields])) OR ((therapy[SH] OR therapies[All Fields] OR therapy [All Fields] OR therape*[All Fields] OR therapis*[All Fields] OR Therapeutics[MH] OR treatment*[All Fields]) AND constructivist*[All Fields]) OR "metacognitive therapies"[All Fields] OR "metacognitive therapy"[All Fields] OR "metacognitive therapeutic"[All Fields] OR "metacognitive therapeutics"[All Fields] OR "metacognitive therapeutical"[All Fields] OR "metacognitive therapeutist"[All Fields] OR "metacognitive therapeutists"[All Fields] OR "metacognitive treatment"[All Fields] OR "metacognitive treatments"[All Fields] OR "meta-cognitive therapies"[All Fields] OR "meta-cognitive therapy"[All Fields] OR "meta-cognitive therapeutic"[All Fields] OR "meta-cognitive therapeutics"[All Fields] OR "meta-cognitive therapeutical"[All Fields] OR "meta-cognitive therapeutist"[All Fields] OR "meta-cognitive therapeutists"[All Fields] OR "meta-cognitive treatment"[All Fields] OR "meta-cognitive treatments"[All Fields] OR "solution-focused therapies"[All Fields] OR "solution-focused therapy"[All Fields] OR "solution-focused therapeutic"[All Fields] OR "solution-focused therapeutics"[All Fields] OR "solution-focused therapeutical"[All Fields] OR "solution focused therapies"[All Fields] OR "solution focused therapy"[All Fields] OR "solution focused therapeutic"[All Fields] OR "solution focused therapeutics"[All Fields] OR "solution focused therapeutical"[All Fields]OR "solution-focussed therapies"[All Fields] OR "solution-focussed therapy"[All Fields] OR "solution-focussed therapeutic"[All Fields] OR "solution-focussed therapeutics"[All Fields] OR "solution-focussed therapeutical"[All Fields]OR "solution focussed therapies"[All Fields] OR "solution focussed therapy"[All Fields] OR "solution focussed therapeutic"[All Fields] OR "solution focussed therapeutics"[All Fields] OR "solution focussed therapeutical"[All Fields] OR "self-control therapies"[All Fields] OR "self-control therapy"[All Fields] OR "self-control therapeutics"[All Fields] OR "self-control therapeutical"[All Fields] OR "self-control therapeutic"[All Fields] OR "self-control training"[All Fields] OR "self-control trainings"[All Fields] OR "self control therapies"[All Fields] OR "self control therapy"[All Fields] OR "self control therapeutics"[All Fields] OR "self control therapeutical"[All Fields] OR "self control therapeutic"[All Fields] OR "self control training"[All Fields] OR "self control trainings"[All Fields]

AND

Depressive Disorder[MH] OR Depression[MH]OR dysthymi*[All Fields] OR "affective disorder"[All Fields]OR "affective disorders"[All Fields] OR "mood disorder"[All Fields] OR "mood disorders"[All Fields] OR depression*[All Fields] OR depressive*[All Fields] OR "dysthymic disorder"[MeSH Terms]

Limits: RCTs

#### *References*

Cuijpers, P., Miguel, C., Harrer, M., Plessen, C. Y., Ciharova, M., Papola, D., Ebert, D., & Karyotaki, E. (2023). Psychological treatment of depression: A systematic overview of a 'Meta-Analytic Research Domain.' *Journal of Affective Disorders*, *335*, 141–151.

Harrer, M., Miguel, C., van Ballegooijen, W., Ciharova, M., Plessen, C. Y., Kuper, P., Sprenger, A. A., Buntrock, C., Papola, D., Cristea, I. A., de Ponti, N., Bašić, Đ., Pauley, D., Driessen, E., Quero, S., Grimaldos, J., Buendía, S. F., Botella, C., Hamblen, J. L., ... Cuijpers, P. (2025). Effectiveness of psychotherapy: Synthesis of a "meta-analytic research domain" across world regions and 12 mental health problems. *Psychological Bulletin*, *151*(5), 600–667. https://doi.org/10.1037/bul0000465

Higgins, J. P. T., Altman, D. G., Gøtzsche, P. C., Jüni, P., Moher, D., Oxman, A. D., Savović, J., Schulz, K. F., Weeks, L., & Sterne, J. A. C. (2011). The Cochrane Collaboration's tool for assessing risk of bias in randomised trials. *BMJ*, *343*, d5928. https://doi.org/10.1136/bmj.d5928

Norcross, J. C. (1990). An eclectic definition of psychotherapy. *What Is Psychotherapy*, *4*, 218–220.

#### Supplement B. Rating manual

##### Metapsy RoB

The questionnaire below aims to facilitate the conduct of risk of bias assessments in the context of psychotherapy trials. The 24 questions evaluate risk of bias arising from five bias domains consistent with current risk of bias frameworks for randomized controlled trials, including Cochrane's RoB 2 (Higgins et al., 2019), with specifications tailored to psychological intervention research. For a correct application, please consult the accompanying manual with all the specifications for each item. For an interactive version of this questionnaire, please visit https://www.metapsy.org/rob/assistant/.

Response options: every question can be responded with Yes/PY (Probably Yes), No/PN (Probably No), or NI (No Information)

| Item | Question |
|---|---|
| **Domain 1: Randomization process** | |
| 1 | Was a random method used to generate the allocation sequence? |
| 2 | Was the allocation sequence kept concealed from those enrolling and assigning participants to interventions? |
| 3 | Is the number of randomized participants balanced between groups? |
| 4 | Is baseline clinical severity balanced between groups? |
| **Domain 2: Deviations from intended interventions** | |
| 5 | Were participants masked to the intervention? |
| 6 | Were carers and people delivering the interventions masked? |
| 7 | If "No/PN" or "NI" to #5 or #6, did the interventions closely align with the intended plan, without any significant deviations that could affect the results? |
| 8 | Were the statistical analyses appropriate for estimating the intention-to-treat effect? |
| 9 | If "No/PN" or "NI" to #8, is the inappropriate analysis unlikely to have an impact on the result? |
| **Domain 3: Missing outcome data** | |
| 10 | Were outcome data obtained from nearly all participants who were originally randomized? |
| 11 | If "No/PN" or "NI" to #10, was an appropriate analysis used for handling the impact of missing data? |
| 12 | If "No/PN" or "NI" to #11, is the trial of moderate to large size (minimum 40 participants per arm reported data)? |
| 13 | If "No/PN" or "NI" to #11, did at least 70% of the randomized participants complete the assessments at the post-treatment? |
| 14 | If "No/PN" or "NI" to #11, is there indirect evidence indicating that missingness is unrelated to the outcome? |
| **Domain 4: Measurement of the outcome** | |
| 15 | Is the method of measurement appropriate and applied using similar procedures for both groups? |
| 16 | Were self-report measures used? |
| 17 | Were assessor-rated instruments used? |
| 18 | If "Yes" to #17, were the assessors masked to treatment allocation? |
| **Domain 5: Selective outcome reporting** | |
| 19 | Is there a trial registration, protocol, or statistical analysis plan (SAP) available? |
| 20 | Write down the registration number, protocol, or link to SAP |
| 21 | If "Yes" to #19, were all outcomes of potential interest for the meta-analysis (scale and time point) pre-specified before unmasked outcome data were available? |

| | |
|---|---|
| 22 | If "Yes" to #21, are all outcomes of potential interest for the meta-analysis fully reported in the paper or available through other means? (e.g., contact with authors) |
| 23 | If "Yes" to #19, was there an analysis plan pre-specified? |
| 24 | If "Yes" to #23, was data analyzed in accordance with the pre-specified plan? |

Higgins, J. P., Savović, J., Page, M. J., Elbers, R. G., & Sterne, J. A. (2019). Assessing risk of bias in a randomized trial. *Cochrane handbook for systematic reviews of interventions*, 205-228.

*Metapsy RoB: Extended manual*

An online version of this manual is available at: https://www.metapsy.org/rob/handbook/

*Introduction to the tool*

The team behind the Metapsy initiative (an initiative for open-access meta-analytical databases, www.metapsy.org) has developed Metapsy RoB to facilitate risk of bias assessment in psychotherapy trials and in large meta-analytic projects. The tool organizes 24 structured items around the five bias domains of Cochrane's RoB 2 (Higgins et al., 2019), which represent the current standard for assessing risk of bias in randomized controlled trials, and accompanies them with psychotherapy-specific guidance developed through expert consensus and meta-epidemiological evidence. This guidance addresses the particular challenges of evaluating risk of bias in psychotherapy research, where standard blinding procedures are unfeasible and context-specific judgment is required. The tool is also designed for large meta-analytic projects involving multiple rater pairs, where inter-rater reliability is a priority. To further facilitate the conduct of ratings, we developed a suite of digital helper tools, all accessible at www.metapsy.org/rob.

*The tool can be used in several ways:*

The most user-friendly option is the **RoB Assistant** (https://www.metapsy.org/rob/assistant/), in which each of the items can be directly answered as an online questionnaire. Brief guidance is provided with each item, and extended guidance can be found in the online handbook or in the current pdf version of the handbook. Using this assistant, reviewers can address all relevant signalling questions step by step. Based on the given answers, the assistant will automatically skip questions that are not required to complete the rating algorithm. Once the questions are completed, the scores per domain and the overall scores are directly calculated through the algorithms that run behind the web-app. The results for the rated study can be downloaded in various formats.

For raters that prefer to use an **Excel version of the tool**, the studies can be rated using the rob-template. This format offers more flexibility to perform the ratings compared to the online RoB Assistant, and may also be more efficient for experienced raters. An important advantage is that it allows users to automatically calculate some items (e.g., unbalanced clinical severity) based on data that has already been collected as part of the usual data extraction [i.e., baseline means and standard deviations for the outcome of interest, type of rating (clinician vs self-report), randomized participants and attrition in each arm, and randomization ratio]. For a correct linkage, the data extraction sheet should be populated according to the Metapsy data standard ("Metapsy database", see an example here). The automatic calculation of some items can be achieved through the Database Application or through the [createRobRatings] function from the metapsyTools R package. Finally, to calculate the scores per domain and the overall RoB score for all the trials, reviewers can upload their rob-template in the Database Application. Optionally, if a Metapsy database is uploaded, the created scores are automatically appended to it. Alternatively, the [createRobRatings] function in R can be used for the same purpose.

We recommend that ratings are performed by two independent reviewers, discussing discrepancies until a consensus is reached, or, where necessary, consulting a senior reviewer. Discrepancies between two independent RoB ratings (either downloaded from the RoB Assistant or performed through the rob-template) can be automatically flagged by using the function [checkRobDiscrepancies] from the metapsyTools R package, which also provides Cohen's kappa as a measure of inter-rater agreement.

Further documentation about the Metapsy RoB tool and the digital helper tools can be found in our documentation page.

*Preliminary considerations*

Metapsy RoB comprises 24 items that assist the conduct of risk of bias assessments in the context of psychological interventions and large meta-analytic projects. All items are formulated in the same direction, in which "Yes/PY" answers are indicative of lower risk of bias.

The current recommendations of this guidance document are applicable for individually randomized parallel-group trials. Some of the core specifications can be applied for cluster randomized trials or crossover trials; however, these types of designs pose specific challenges that should be further evaluated. Further guidance for these types of designs is available in Cochrane's RoB 2 documentation (Higgins et al., 2019).

As recommended in the risk of bias literature (e.g., Cochrane's RoB 2 guidelines), assessment of risk of bias should be specific to a particular result (e.g., standardized mean difference between psychotherapy and control condition) for a particular outcome (e.g., depression severity) and measured at a particular time (e.g., post-test), although some biases may apply to the whole study.

The recommendations of this document are aimed at examining the **effect of assignment to intervention**, given that this is the type of assessment of primary choice by clinical guideline developers and policy makers, and also typically of Metapsy databases. However, if reviewers wish to assess the effect of adhering to intervention, Cochrane's RoB 2 tool documentation contains a section specific to that aim.

*[createRobRatings]*

Variables marked with the tag [createRobRatings] do not need to be answered manually by the reviewers if there is a Metapsy formatted dataset ("Metapsy database", see an example here) that has already collected required variables for these items: baseline means and standard deviations, type of rating (clinician vs self-report), randomized participants and attrition in each arm, and randomization ratio (baseline_m_arm1/arm2, baseline_sd_arm1/arm2, baseline_n_arm1/arm2, rand_ratio, rand_arm1/rand_arm2, attr_arm1/attr_arm2 and rating). The items that can be pre-filled automatically via createRobRatings when a Metapsy database is available are: 3, 4, 10, 12, 13, 16, and 17.

#### Domain 1: Randomization process

1) **Was a random method used to generate the allocation sequence?**

Answer 'Yes/PY' if the trial used any method involving chance to generate the allocation sequence, such as computerized randomization, random number tables, or similar approaches.

2) **Was the allocation sequence kept concealed from those enrolling and assigning participants to interventions?**

Answer 'Yes/PY' if the trial used a system ensuring that those enrolling participants could not predict or influence upcoming assignments, for example through central telephone or web-based allocation systems, or properly sealed opaque envelopes.

3) **Is the number of randomized participants balanced between groups? [createRobRatings]**

Imbalance in the number of randomized participants can be examined through the proportion assigned to one group and its 99% confidence intervals. If the intervals do not cover the expected proportion (e.g., 50% in a 1:1 allocation), there is suspicion of baseline imbalance. The calculations needed for this item can also be performed at https://www.metapsy.org/rob/assistant/.

4) **Is baseline clinical severity balanced between groups? [createRobRatings]**

Imbalance between groups in baseline severity is examined by the standardized mean difference between groups at baseline and its 99% confidence intervals. An SMD > 0.2 (with 99% CI lower limit > 0) will be suggestive of baseline imbalance. We focus on baseline severity only because it is consistently reported across trials, can be examined objectively, and it is a known predictor of the outcome. P-values and confidence intervals resulting from multiple significance testing (e.g., due to multiple clinical severity measures) can be adjusted using Bonferroni corrections. Depending on the clinical field and mental health disorder, there might be key prognostic factors that should be added to this examination. In that case, the judgment of the reviewers can override the algorithm, and the proposed items and further variables can be examined for baseline imbalances. The calculations needed for this item can also be performed at https://www.metapsy.org/rob/assistant/.

#### Domain 2: Deviations from intended interventions

5) **Were participants masked to the nature of the intervention?**

In almost all psychotherapy trials, this will be No. Exceptions can be specifically studied for the trials comparing two active psychological interventions (e.g., both active interventions are presented to participants as equally effective, the trial is designed as a non-inferiority trial, etc.).

6) **Were carers and people delivering the interventions masked?**

In almost all psychotherapy trials, this will be No. Exceptions can be specifically studied for the trials comparing two active psychological interventions (e.g., both active interventions are presented to participants as equally effective, the trial is designed as a non-inferiority trial, etc.).

7) **<u>If "No/PN" or "NI" to #5 or #6</u>, did the interventions closely align with the intended plan, without any significant deviations that could affect the results?**

When participants and carers are not blinded, there could be deviations from the intended interventions associated with this lack of blinding. An example of such a type of deviation is when participants who are told that they were on the waitlist seek the intervention of the experimental arm outside the trial or other interventions. Another example could be that, due to the trial context, a substantially larger number of participants in the psychotherapy group make use of treatment as usual than the treatment as usual control group. For example, participants in the psychotherapy group have easier access to the prescription of antidepressant medication than participants in the control group. Deviations from the intended interventions that would arise even if the intervention took place outside the trial do not constitute a risk of bias with regard to the intention-to-treat analysis. An example could be drop-outs due to side effects of antidepressants: such drop-outs are deviations from the intended intervention but would occur even if the patients were taking antidepressants outside the trial context.

To pose a risk, these deviations should be influential for the outcome. For example, participants taking benzodiazepines in a depression psychotherapy trial might not be influential, but receiving the psychotherapy of interest or antidepressant medication is influential.

If there are deviations related to the outcome, but they are balanced between the groups, bias will be less likely. However, **if these influential deviations are unbalanced between the groups,** it is more likely that the intervention effect estimate is biased. For example, when a substantially larger proportion of participants allocated to the psychotherapy group are taking antidepressants compared to the control group. Reviewers should make agreements on how an imbalance between groups is defined. In the previous example, an imbalance of 20% could be taken as a rough indication. In these cases, the trial can be **judged as high risk,** by giving a “No/PN” answer to this item.

8) **Were the statistical analyses appropriate for estimating the intention-to-treat effect?**

This item evaluates whether trialists report to have adhered to the intention-to-treat principles, analyzing participants according to the group to which they were originally assigned (regardless of the intervention finally received) and including all randomized participants in the analysis (regardless of the actual adherence of the participants).

Approaches such as “per-protocol” analyses (exclusion of those who did not receive the allocated intervention) or “as treated” analysis (analysis of participants according to the intervention received, rather than the originally assigned) should be considered inappropriate. For example, it is not appropriate if participants are excluded due to not receiving a minimum number of treatment sessions. In some cases, it might be sensible to assume that trialists adhered to ITT principles even if it is not explicitly stated (e.g., when authors used statistical analyses that are implicitly associated with the ITT principle).

9) **If “No/PN” or “NI” to #8, is the inappropriate analysis unlikely to have an impact on the result?**

If no appropriate analysis was conducted (or there is not enough information), reviewers should examine whether there was likely an impact of not using an appropriate analysis to estimate the effect of assignment to intervention. In cases in which excluded participants (e.g., due to not receiving a minimum number of sessions) or participants analyzed in the wrong group (e.g., participants randomized to the intervention group are analyzed in the waitlist group because in the end they did not receive the intervention) are less than 5%, this item should be rated as “Yes/PY” (leading to some concerns). Reviewers should answer “NI” if there is not enough information, and answer “No/PN” for thresholds above 5%.

#### Domain 3: Missing outcome data

**10) Were outcome data obtained from nearly all participants who were originally randomized? [createRobRatings]**

For meta-analyses focused on **continuous outcomes:** if the proportion of available data is above 95% for both groups, then domain 3 is directly rated as low risk and it is not needed to answer the next questions. Note that this refers to the proportion of participants who have endpoint data (i.e., who completed the questionnaires), and not the LOCF or imputed data.

For meta-analyses focused on **dichotomous outcomes**: For dichotomous outcomes, the proportion should be directly related to the risk of the event. If the observed number of events is much larger than the number of participants with missing data, the risk of bias will be less likely.

**11) If "No/PN" or "NI" to #10, was an appropriate analysis used for handling the impact of missing data?**

For example, the following approaches are considered appropriate for handling the impact of missing data:

- MMRM (mixed models for repeated measures, also known as mixed models, growth curve analyses) based on two or more measurements after baseline (e.g., mid-treatment, post-test, follow-ups, etc.).
- Multiple imputation, if the following criteria are met:
  - "Rubin's rules" or other methods to account for imputation uncertainty were applied.
  - Important auxiliary variables were used in the imputation model, such as intermediate outcome assessments.
  - Imputations were generated separately by RCT groups ("bygroup" imputation).
  - Controlled/reference-based imputation (e.g., "jump-to-reference") is used, using appropriate external information.
  - Sensitivity analyses corresponding with a range of plausible reasons for missingness to confirm the primary analyses.

"Last observation carried forward" is not considered an appropriate approach for handling missing data.

*If no appropriate analyses or unclear/no information, answer the next questions.*

**12) If "No/PN" or "NI" in #11, is the trial of moderate to large size (minimum 40 participants per arm reported data)? [createRobRatings]**

**13) If "No/PN" or "NI" in #11, did at least 70% of the randomized participants in each of the arms complete the assessments at the post-treatment? [createRobRatings]**

**14) If "No/PN" or "NI" in #11, is there indirect evidence indicating that missingness is unrelated to the outcome?**

This item examines the following sources of indirect evidence that might signal risk of bias:

a) whether the overall *reported reasons for missing data indicate that missingness might depend on its true value* (e.g., the general reason for missing data is termination of funding would not be indicative of risk of bias),

b) *large imbalances in the proportion of missing data between the groups*: As a rule of thumb, the difference in study drop-out between the groups should be less than 20% (e.g., the intervention arm has a study drop-out of 10% and the waitlist control has a drop-out of 20%),

c) *differing reasons for missing data between the groups* (e.g., more participants from the intervention group abandon the trial due to being more critically symptomatic could be an indicator of missingness being related to the outcome).

If one of the three is present, reviewers should answer with a "No/PN", given that missing data might be related to the outcome. To answer with a "Yes/PY" there should be enough information to evaluate these possible sources. Reviewers may answer this question with "NI" if there is not enough information to assess the sources.

#### Domain 4: Measurement of the outcome

**15) Is the method of measurement appropriate and applied using similar procedures for both groups?**

Included instruments are valid and reliably measure the outcome domain of interest in the meta-analysis.

**16) Were self-report measures used? [createRobRatings]**

In the context of psychological interventions, blinding is often not feasible or even impossible to achieve. Empirical evidence shows that self-reports are not associated with overestimated treatment effects despite the lack of participant blinding (Miguel et al., 2025; Cuijpers et al., 2010). Therefore, if there is no evidence that participants under-reported or there is a safeguard to prevent this (e.g., therapists did not have access to the self-reports), reviewers may consider self-reports as low risk of bias. If it is likely that patients under-reported their severity (for example, to please the therapists or the researchers when self-reports are filled out in front of the therapist at the last session), reviewers should consider assessing item #16 as "No/PN", which will lead to high risk of bias for domain 4.

**17) Were assessor-rated instruments used? [createRobRatings]**

Assessor-rated instruments are filled out by the therapist, independent evaluators, or research personnel. These are usually administered as a structured interview with the participant or are completed by the evaluator through observation of the participant's behavior.

**18) If yes to #17, were the assessors masked to treatment allocation?**

This item evaluates whether the assessors evaluating the outcome were masked to treatment allocation. This item should be rated as "Yes" when trialists clearly report that the assessors were masked. If this is not reported, it might be sensible to assume that no masking procedures were implemented ("No/PN").

*For meta-analyses including multiple outcome measures from the same trial:*

Meta-analysts might be interested in including all available results for one outcome domain within a trial. For example, in a depression trial, there might be post-test data from the Hamilton Depression Rating Scale (assessor-rated) and the Beck Depression Inventory (self-report). There could be two strategies for these cases, depending on the meta-analysis protocol and analysis approach:

1) Separate risk of bias scores could be performed for each outcome and numeric result, or

2) Two or more ratings could be combined into an aggregated score, conservatively rating the aggregated score as high risk when one of the measurements is at high risk.

For the Metapsy depression database we use the second strategy, combining ratings into an aggregated score.

#### Domain 5: Selective outcome reporting

Similarly to the previous domain, the evaluation of this domain will depend on the meta-analysts' protocol regarding the choice of outcome measures. Some meta-analysts might decide to include all available measurements for one outcome domain (e.g., depression symptoms) (even if this entails including multiple measurements from within a study), while others might decide on more restrictive approaches (e.g., including only assessor-rated depression severity). To appropriately evaluate the selection of the reported result, reviewers should take into account the meta-analysis protocol for inclusion of outcome measures and results.

**19) Is there a trial registration, protocol, or statistical analysis plan (SAP) available?**

If there is no access to documents describing the trial's protocol pre-defined outcome measures or analysis plan reviewers should answer "NI" in this item. The trial can then be rated as "Some concerns" for domain 5 and the ratings stop here.

If reviewers have access to such documents, the next questions should be answered.

**20) Please write down the registration number, protocol, or link to SAP.**

**21) If "Yes/PY" to #19, were outcomes (scale and time point) pre-specified before unmasked outcome data were available?**

This item evaluates whether the outcome measures in a trial were pre-specified before the start of data collection (i.e., within one month of the start of participant enrolment). For example, when the trial was registered (and specified the outcomes) before the start of data collection, which is known as prospective registration. To evaluate whether registration of outcomes was prospective, reviewers can compare the date in which recruitment started and the date in which the registration was posted (e.g., in ClinicalTrials.gov). This differs per trial registry, as some registries already give the information of whether the registration was prospective (e.g., ISRCTN Registry). It is important to also examine the "history of changes" that is available in some of the registries, as unjustified changes throughout the trial after unmasked data was available could have occurred.

Outcome domain, scale, time point, metric, and analysis should ideally be specified, although reviewers can answer this item as "Yes" if at least scale and time point were prospectively specified.

Retrospective registrations (registering the trial after start of data collection) and lack of information (no scale/time specified) in a prospective registration should lead to "NI" (respectively) in this item, both resulting in "Some concerns" in domain 5.

**22) If "Yes/PY" to #21, are all outcomes of interest for the meta-analysis fully reported in the paper or available through other means?**

This will partly depend on the meta-analysis protocol. If meta-analysts aim to include all available instruments for a given outcome (e.g., depression severity), the article should report all instruments in full (with enough data to be entered in the meta-analysis). If meta-analysts have a pre-specified hierarchy for instrument inclusion (e.g., select HAM-D over BDI), then selective reporting will be judged regarding the availability of outcomes based on the pre-specified hierarchy. It will also be rated as high risk when a non-registered outcome is added in the publication of the paper. Reviewers should answer this question with “No” in this case. Reviewers should consider whether data from the outcome of interest can be made available through other means (e.g., contact with authors).

**23) If “Yes/PY” to #19, was there an analysis plan pre-specified?**

Pre-specified plan = before unblinded outcome data is available.

Dates of publication of the analysis plan should precede participant enrollment. Published protocols: Submission date of the published protocol precedes participant enrollment. Reviewers should answer with “NI” if it is not clear whether the protocol reflects a pre-specified plan.

**24) If “Yes/PY” to #23, was data analyzed in accordance with the pre-specified plan?**

Any deviation from the original plan should be reported in the final publication and should be justified. If properly justified, meta-analysts can rate #24 as low risk, after careful assessment of how this deviation can affect the studied meta-analytic result.

We will not consider item #24 in the algorithm for domain 5 unless there is evidence of unjustified deviations from a pre-specified analysis plan, in which case the meta-analysts should consider downgrading the domain to high risk when necessary.

*Algorithms for calculating scores*

The scores per domain and the overall score for each of the rated trials can be easily calculated using the *metapsyTools* package in RStudio. For this, reviewers should import the rating template (rob-template) in RStudio and use the [createRobRatings] function. Alternatively, the scores can also be calculated by uploading the (rob-template) in our user-friendly web application at: https://www.metapsy.org/rob/database-app/.

Note: “No” also includes “PN” (probably No) response option, and “Yes” also includes “PY” (probably Yes) response option.

**Proposed algorithm domain 1**

2 = No → High risk
2 = NI, and either 3 or 4 = No → High risk
2 = NI, and both 3 and 4 = Yes or NI → Some concerns
2 = Yes, 1 = No → Some concerns
2 = Yes, 1 = Yes or NI, and either 3 or 4 = No → Some concerns
2 = Yes, 1 = Yes or NI, and both 3 and 4 = Yes or NI → Low risk

**Proposed algorithm domain 2**

5 AND 6= Yes → Low risk
7= NI or Yes, and 8= Yes → Low risk
7= NI or Yes, 8= No/NI, and 9= Yes → Some concerns
7= NI or Yes, 8= No/NI, and 9= No/NI → High risk
7= No, any answer for 8/9 → High risk

*Note. Item 7 only downgrades the assessment when there is clear evidence of deviations that might have influenced the outcome.*

##### Proposed algorithm domain 3

10 = Yes → Low risk
10= No/NI, 11= Yes → Low risk
11= No/NI (analysis), 12= Yes (larger trial), 13= Yes (70% data), 14= Yes (missing unrelated) → Some concerns
11= No/NI (analysis), 12= Yes (larger trial), 13= No/NI (70% data), 14= Yes (missing unrelated) → Some concerns
11= No/NI (analysis), 12= Yes (larger trial), 13= Yes (70% data), 14= No/NI (missing unrelated) → Some concerns
11= No/NI (analysis), 12= Yes (larger trial), 13= No/NI (70% data), 14= No/NI (missing unrelated) → High risk
11= No/NI (analysis), 12= No/NI (smaller), 13= Yes (70% data), 14= Yes (missing unrelated) → Some concerns
11= No/NI (analysis), 12= No/NI (smaller), 13= No/NI (70% data), 14= Yes (missing unrelated) → High risk
11= No/NI (analysis), 12= No/NI (smaller), 13= Yes (70% data), 14= No/NI (missing unrelated) → High risk
11= No/NI (analysis), 12= No/NI (smaller), 13= No/NI (70% data), 14= No/NI (missing unrelated) → High risk

##### Proposed algorithm domain 4

15=No → High risk
15= Yes, 16= Yes, 17= No/NI → Low risk
15= Yes, 16= No/NI, 17= Yes, 18= Yes → Low risk
15= NI, 16= Yes, 17= No/NI → Some concerns
15= NI, 16= No/NI, 17= Yes, 18= Yes → Some concerns
15= Yes/NI, 16= No/NI, 17= Yes, 18= No/NI → High risk
(For meta-analyses including trials with multiple instruments/types of assessments)
15=No → High risk
15= Yes, 16= Yes, 17= Yes, 18= Yes → Low risk
15= Yes, 16= Yes, 17= No → Low risk
15= NI, 16= Yes, 17= Yes, 18= Yes → Some concerns
15= Yes/NI/No, 16= Yes, 17= Yes, 18= No/NI → High risk

##### Proposed algorithm domain 5

19= NI/No → Some concerns
21 = No/NI → Some concerns
22= Yes → Low risk
22= NI → Some concerns
22= No → High risk
22= NI, 24= No → High risk
22= Yes, 24= No → High risk

*Note. Item 24 only downgrades the score in the algorithm if the answer is "No" (i.e., when there is evidence of unjustified deviations from the analysis plan).*

##### Overall score

**Low risk** requires all domains rated as low risk.
**High risk** is assigned when at least one domain is rated as high risk, or when three or more domains have some concerns.
**Some concerns** is assigned if at least one domain has some concerns and there is a maximum of 2 domains with some concerns.

#### *References*

Higgins, J. P., Savović, J., Page, M. J., Elbers, R. G., & Sterne, J. A. (2019). Assessing risk of bias in a randomized trial. *Cochrane handbook for systematic reviews of interventions*, 205-228.
Miguel, C., Harrer, M., Karyotaki, E., Plessen, C. Y., Ciharova, M., Furukawa, T. A., ... & Cuijpers, P. (2025). Self-reports vs clinician ratings of efficacies of psychotherapies for depression: a meta-analysis of randomized trials. *Epidemiology and Psychiatric Sciences*, 34, e15.
Cuijpers, P., Li, J., Hofmann, S. G., & Andersson, G. (2010). Self-reported versus clinician-rated symptoms of depression as outcome measures in psychotherapy research on depression: a meta-analysis. *Clinical psychology review,* 30(6), 768-778.

#### Supplement C. Rating sheet and digital tools

The full suite of digital tools associated with Metapsy RoB can be found here: https://www.metapsy.org/rob.

##### Screenshots of the Excel rating sheet

(https://www.metapsy.org/assets/files/rob-template.xlsx)

| **RoB ratings specifications** | | | | **1 ) Randomization process** | | | | |
|---|---|---|---|---|---|---|---|---|
| **metapsyTools** | | **[createRobRatings]** | | | | **[createRobRatings]** | | |
| **Question** | | **comparisons** | **instruments** | **#1** | **#2** | **#3** | **#4** | |
| **Rater ID or name** | **Study's unique ID** | Comparisons for which RoB is rated | Instruments for which RoB is rated | Was a random method used to generate the allocation sequence? | Was the allocation sequence kept concealed from those enrolling and assigning participants to interventions? | Is the number of randomized participants balanced between groups? | Is baseline clinical severity balanced between groups? | Notes for domain 1 |
| **rater** | **study** | **comparisons** | **instruments** | **d1_1** | **d1_2** | **d1_3** | **d1_4** | **d1_notes** |
| CM | Author, 2022 | | | Yes/PY | Yes/PY | | | |

| **2) Deviations from intended interventions** | | | | | |
|---|---|---|---|---|---|
| **#5** | **#6** | **#7** | **#8** | **#9** | |
| Were participants masked to the intervention? | Were carers and people delivering the interventions masked? | **If "No/PN" or "NI" to #5 or #6,** Did the interventions closely align with the intended plan, without any significant deviations that could affect the results? | Were the statistical analyses appropriate for estimating the intention-to-treat effect? | **If "No/PN" or "NI" to #8,** is the inappropriate analysis unlikely to have an impact on the result? | Notes for domain 2 |
| **d2_5** | **d2_6** | **d2_7** | **d2_8** | **d2_9** | **d2_notes** |
| No/PN | No/PN | NI | Yes/PY | | |

| **3) Missing outcome data** | | | | | |
|---|---|---|---|---|---|
| **[createRobRatings]** | | **[createRobRatings]** | **[createRobRatings]** | | |
| **#10** | **#11** | **#12** | **#13** | **#14** | |
| Were outcome data obtained from nearly all participants who were originally randomized? | **If "No/PN" or "NI" to #10,** was an appropriate analysis used for handling the impact of missing data? | **If "No/PN" or "NI" to #11,** is the trial of moderate to large size (minimum 40 participants per arm reported data)? | **If "No/PN" or NI to #11,** did at least 70% of the randomized participants in each of the arms complete the assessments at the post-treatment? | **If "No/PN" or "NI" to #11,** is there indirect evidence indicating that missingness is unrelated to the outcome? | Notes for domain 3 |
| **d3_10** | **d3_11** | **d3_12** | **d3_13** | **d3_14** | **d3_notes** |
| | NI | | | Yes/PY | |

| 4) Measurement of the outcome | | | | |
|---|---|---|---|---|
| | [createRobRatings] | [createRobRatings] | | |
| **#15** | **#16** | **#17** | **#18** | |
| Is the method of measurement appropriate? | Were self-report measures used? | Were assessor-rated instruments used? | **If "Yes" to #17,** were the assessors masked to treatment allocation? | Notes for domain 4 |
| **d4_15** | **d4_16** | **d4_17** | **d4_18** | **d4_notes** |
| Yes/PY | | | | |

| 5) Selective outcome reporting | | | | | | |
|---|---|---|---|---|---|---|
| **#19** | **#20** | **#21** | **#22** | **#23** | **#24** | |
| Is there a trial registration, a protocol, or statistical analysis plan (SAP) available? | Write down the registration number, protocol, or link to SAP | **If "Yes/PY" to #19,** were outcomes (scale and time point) pre-specified before unmasked outcome data were available? | **If "Yes/PY" to #21,** are all outcomes of interest for the meta-analysis fully reported or available through other means? | **If "Yes/PY" to #19,** Was there an analysis plan pre-specified? | **If "Yes/PY" to #23,** was data analyzed in accordance with the pre-specified plan? | Notes for domain 5 |
| **d5_19** | **d5_20** | **d5_21** | **d5_22** | **d5_23** | **d5_24** | **d5_notes** |
| NI | | | | | | |

*Screenshots of the digital applications*

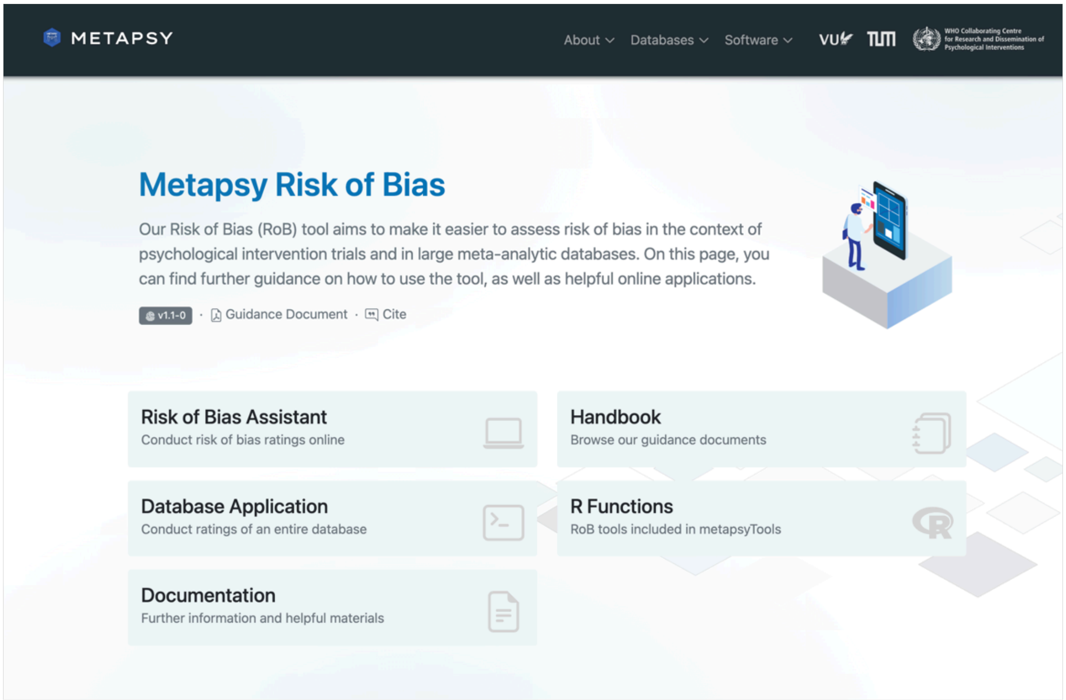
METAPSY
About
Databases
Software
Metapsy Risk of Bias
Our Risk of Bias (RoB) tool aims to make it easier to assess risk of bias in the context of psychological intervention trials and in large meta-analytic databases. On this page, you can find further guidance on how to use the tool, as well as helpful online applications.
v1.1-0 · Guidance Document · Cite
Risk of Bias Assistant
Conduct risk of bias ratings online
Handbook
Browse our guidance documents
Database Application
Conduct ratings of an entire database
R Functions
RoB tools included in metapsyTools
Documentation
Further information and helpful materials

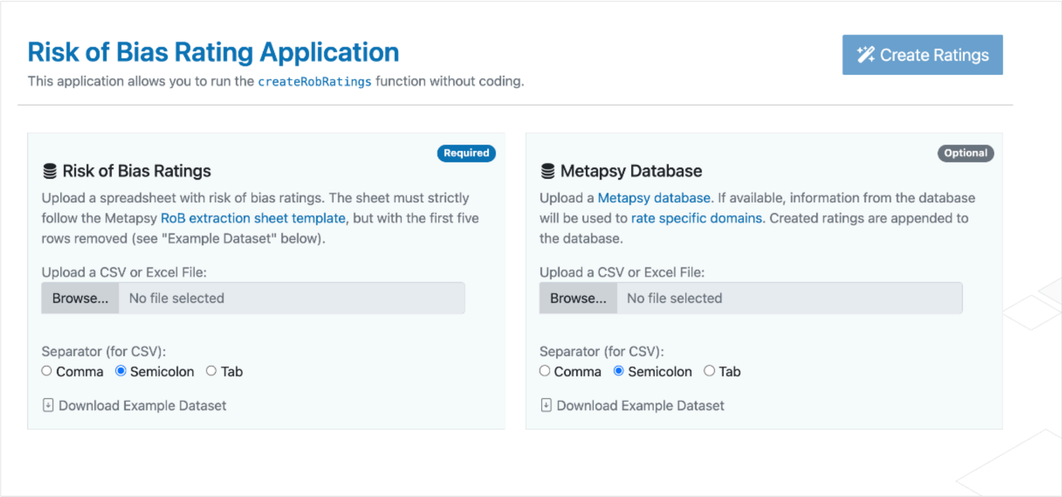
Risk of Bias Rating Application
Create Ratings
This application allows you to run the createRobRatings function without coding.
Required
Risk of Bias Ratings
Upload a spreadsheet with risk of bias ratings. The sheet must strictly follow the Metapsy RoB extraction sheet template, but with the first five rows removed (see "Example Dataset" below).
Upload a CSV or Excel File:
Browse... No file selected
Separator (for CSV):
Comma Semicolon Tab
Download Example Dataset
Optional
Metapsy Database
Upload a Metapsy database. If available, information from the database will be used to rate specific domains. Created ratings are appended to the database.
Upload a CSV or Excel File:
Browse... No file selected
Separator (for CSV):
Comma Semicolon Tab
Download Example Dataset

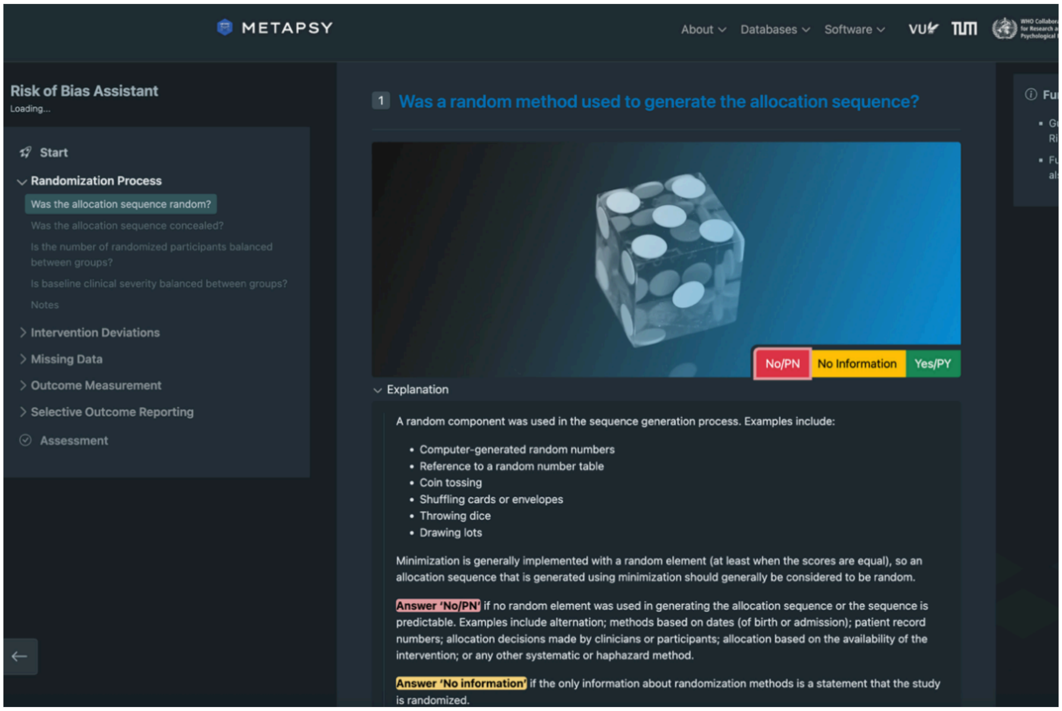
METAPSY
About
Databases
Software
Risk of Bias Assistant
Loading...
Start
Randomization Process
Was the allocation sequence random?
Was the allocation sequence concealed?
Is the number of randomized participants balanced between groups?
Is baseline clinical severity balanced between groups?
Notes
Intervention Deviations
Missing Data
Outcome Measurement
Selective Outcome Reporting
Assessment
1 Was a random method used to generate the allocation sequence?
No/PN
No Information
Yes/PY
Explanation
A random component was used in the sequence generation process. Examples include:
Computer-generated random numbers
Reference to a random number table
Coin tossing
Shuffling cards or envelopes
Throwing dice
Drawing lots
Minimization is generally implemented with a random element (at least when the scores are equal), so an allocation sequence that is generated using minimization should generally be considered to be random.
Answer 'No/PN' if no random element was used in generating the allocation sequence or the sequence is predictable. Examples include alternation; methods based on dates (of birth or admission); patient record numbers; allocation decisions made by clinicians or participants; allocation based on the availability of the intervention; or any other systematic or haphazard method.
Answer 'No information' if the only information about randomization methods is a statement that the study is randomized.

#### Supplement D. Extended Background on the Development and Rationale for Metapsy RoB

This supplement provides an extended account of the conceptual and empirical background that shaped Metapsy RoB, the structured risk of bias assessment tool used in the present study. It summarizes the rationale, the developmental process, and the domain-by-domain reasoning behind the tool's specifications for psychotherapy trials. The operational content of the tool itself, its 24 items, scoring algorithms, and rating instructions, is presented separately in Supplement B. This supplement instead focuses on why each specification was chosen and on the empirical and theoretical evidence that supports it.

##### *1. Background and rationale for Metapsy RoB*

Systematic reviews and meta-analyses of randomized controlled trials are widely regarded as providing the most trustworthy evidence for clinical decision making and policy (Group, 1992; Steinberg, Greenfield, Wolman, Mancher, & Graham, 2011). The credibility of their conclusions depends directly on the quality of the primary trials on which they are based. Flaws in trial design or execution, such as improper randomization or selective reporting, can distort effect estimates in either direction, so evaluating this risk of bias is an essential step in any systematic review.

Cochrane's Risk of Bias tool is the most widely used method for this purpose across disciplines (Jørgensen et al., 2016). Its current version, RoB 2 (Sterne et al., 2019), evaluates bias across five domains, namely the randomization process, deviations from intended interventions, missing outcome data, measurement of the outcome, and selection of the reported result, using a series of signalling questions that feed into domain- and study-level algorithms. These five domains are broadly consistent with other current risk of bias frameworks for randomized trials.

Independent evaluations of RoB 2 have described substantial practical difficulties in its application, including low inter-rater agreement, long assessment times of around 30 minutes per study, and considerable overall complexity (Minozzi, Cinquini, Gianola, Gonzalez-Lorenzo, & Banzi, 2020). A study focused on complex interventions reached similar conclusions, finding that several signalling questions were too theoretical to apply without further practical guidance. Missing outcome data, measurement bias in unblinded trials, and deviations from intended interventions were identified as the domains most consistently difficult to rate (Crocker et al., 2023; Minozzi et al., 2020).

These general difficulties are compounded in psychotherapy research. Unlike pharmacological trials, psychotherapy trials cannot achieve double blinding. Therapists cannot be masked to the treatment they deliver, and there is no placebo equivalent that would allow participants to remain unaware of their allocation (Cuijpers, 2025). This creates specific uncertainty around deviations from intended interventions and the use of self-reported outcomes, and in practice has led reviewers to apply markedly different, sometimes idiosyncratic, criteria for these domains, or to omit them from assessment altogether (Munder & Barth, 2018). Because this inconsistency can produce divergent risk of bias conclusions for the same trial across different reviews, it has direct downstream consequences for the coherence of clinical guidelines that rely on this evidence.

The methodological literature has suggested that developing a review-specific implementation document can facilitate consistent application, reduce assessment time, and increase inter-rater reliability (Crocker et al., 2023; Minozzi, Dwan, Borrelli, & Filippini, 2022; Tomlinson et al., 2024). At the time this work was carried out, there were general recommendations about how to consider RoB in the context of psychotherapy trials (Munder & Barth, 2018), but there was no prior published work with specific recommendations and a structured risk of bias assessment developed for psychotherapy trials. This gap, together with the practical need to rate risk of bias reliably and reproducibly across the large, living Metapsy meta-analytic database of psychotherapy trials for depression (Cuijpers, Harrer, Miguel, Ciharova, & Karyotaki, 2023), motivated the development of Metapsy RoB.

*2. Development process*

The development of Metapsy RoB proceeded in two stages.

**Preliminary assessment.** A sample of 35 trials comparing psychotherapies for depression with control conditions, drawn from the Metapsy database, was independently rated by pairs of reviewers, all members of the author team of the original development work, using Cochrane's RoB 2 documentation and guidance as a reference point. Reviewers systematically documented the items and domains that generated the most discrepancies, uncertainty, and difficulties specific to the nature of psychological interventions. This exercise identified challenges across all five domains, including inconsistent handling of baseline imbalances (Domain 1), general difficulty defining and rating blinding and deviations from intended interventions (Domain 2), inconsistent choices of appropriate analyses for missing data (Domain 3), divergent interpretation of self-report instruments (Domain 4), and inconsistent procedures for evaluating pre-specified outcomes (Domain 5). These findings formed the empirical starting point for the tool's development.

**Expert consensus process.** Following the preliminary assessment, experienced psychotherapy researchers from the authorship team (PC, TAF, EK, ES, MS, CM) held monthly videoconference meetings over one year, working through the challenges identified for each domain, with particular attention to the items that had generated the most difficulty. Additional input was sought from experts in the wider research network for specific indicators. Several rounds of piloting were conducted between meetings, in which pairs of experts independently rated samples of trials using draft versions of the proposed criteria. Piloting continued iteratively until consensus was reached. The result of this process was a final tool built around 24 standardized items, all scored in the same direction so that Yes or Probably Yes always indicates lower risk of bias, with harmonized scoring algorithms.

Metapsy RoB's five domains are consistent with current risk of bias frameworks for randomized controlled trials, including Cochrane's RoB 2, and its items and guidance were developed with specifications tailored to psychological intervention research. It was developed primarily for individually randomized, parallel-group trials assessing the intention-to-treat effect, as these represent the most common design in the psychotherapy literature. Most items remain applicable to cluster randomized trials with the addition of design-specific signalling questions, but cluster and crossover designs raise further considerations not fully addressed here.

*3. General preliminary considerations*

Two general decisions shaped the interpretation of several domains and are described here before turning to domain-specific rationale.

**Effect of interest.** A distinction is commonly drawn between the effect of assignment to intervention, or intention-to-treat effect, and the effect of adhering to intervention, or per-protocol effect. This choice affects the assessment of Domain 2 in particular (Higgins et al., 2019). Metapsy RoB focuses on the effect of assignment to intervention, as this is the effect typically prioritized by clinical guideline developers and policymakers.

**Level of assessment.** As is standard practice in current risk of bias frameworks, assessment is in principle carried out for a specific result, outcome, and time point, although some biases apply to the whole study (Sterne et al., 2019).

##### 4. Domain-specific rationale

*Domain 1. Randomization process*

Randomization is intended to balance prognostic factors across groups and thereby prevent confounding. Two mechanisms protect this balance. The first is allocation sequence generation through a genuinely random process. The second is allocation sequence concealment, which prevents trial staff or participants from anticipating and thereby influencing the next assignment (Higgins et al., 2019). Current risk of bias frameworks also examine baseline imbalances as an indirect signal of problems with randomization, for example unexpected differences in group size relative to the intended allocation ratio, an excess of statistically significant baseline differences, or imbalance in key prognostic factors exceeding what would be expected by chance.

A key consideration for psychotherapy trials concerns which prognostic factors to examine, since this must be tailored to the intervention and clinical population under study. Two aspects were selected for Metapsy RoB. The first is imbalance in the number of randomized participants, evaluated through the proportion allocated to one group and its 99% confidence interval, with intervals excluding the expected proportion, for example 50% under a 1:1 allocation, considered suggestive of imbalance. The second is imbalance in baseline clinical severity, evaluated through the standardized mean difference between groups at baseline and its 99% confidence interval, with a standardized mean difference above 0.2 and a lower 99% confidence interval limit above 0 considered suggestive of imbalance. Baseline severity was selected as the key prognostic factor because it is reported consistently across trials, can be assessed objectively, and has repeatedly been shown to be a robust predictor of treatment outcome in psychological interventions for depression (Driessen, Cuijpers, Hollon, & Dekker, 2010; Bower et al., 2013; Karyotaki et al., 2018; Reins et al., 2021; Kuyken et al., 2016; Furukawa et al., 2018; Furukawa et al., 2021; Cuijpers et al., 2022), anxiety (Rosenfield et al., 2019; Scholten et al., 2023), and post-traumatic stress disorder (de Haan et al., 2024; Wright et al., 2024). Other prognostic factors may be more relevant depending on the mental disorder and clinical context under review, in which case Metapsy RoB should be adjusted accordingly.

*Domain 2. Deviations from intended interventions*

Because Metapsy RoB targets the effect of assignment to intervention, this domain assesses bias arising when the interventions actually delivered depart from what was pre-specified in the trial protocol. Current risk of bias frameworks direct reviewers to evaluate three elements, namely whether participants and personnel were blinded, whether deviations occurred that were attributable to the trial context, and whether an appropriate analysis was used to estimate the effect of assignment (Higgins et al., 2019).

Deviations are only relevant insofar as they depart from what was pre-specified, including any protocol-permitted adjustments in response to participant status, and this requires a precise definition of both the active and control conditions. In practice, control conditions such as care as usual are frequently vague or inconsistently described in psychotherapy trials, which complicates this assessment (Watts, Turnell, Kladnitski, Newby, & Andrews, 2015; Cuijpers, Quero, Papola, Cristea, & Karyotaki, 2021). A key type of deviation is the receipt of non-protocol interventions, also called co-interventions, that were not part of the specified plan. Whether these introduce bias depends on whether they are related to the outcome and on the review's research question, and even interventions not explicitly listed in a trial's own protocol may be acceptable if they are consistent with the broader definition used across the review (Higgins et al., 2019).

Blinding is central to this domain because unsuccessful blinding is a common route through which deviations arise. In psychotherapy, blinding of participants and personnel is not merely difficult but, in most conceptualizations of therapeutic change, undesirable. Participant awareness of and engagement with the intervention is considered a necessary mechanism of change rather than a threat to validity (Baskin, Tierney, Minami, & Wampold, 2003; Munder & Barth, 2018; Doering, Glombiewski, & Rief, 2018; Wampold & Imel, 2015). The absence of masking can nonetheless introduce bias through subtle psychological mechanisms, for instance reduced motivation to engage in problem-solving among waitlist participants who know they will eventually receive the active intervention. This mechanism is empirically supported, as waitlist-controlled trials are consistently associated with overestimated psychotherapy effects relative to trials using more robust control conditions (Furukawa et al., 2014; Cuijpers, Miguel, Harrer, Ciharova, & Karyotaki, 2024; Michopoulos et al., 2021). Because these subtle effects are difficult to observe directly, Metapsy RoB instructs reviewers to infer them indirectly through unbalanced deviations from the protocolized interventions arising specifically from the trial context, as opposed to deviations that would have occurred equally in routine care, rather than through the presence or absence of masking as such (Higgins et al., 2019). This reasoning is also informed by the broader literature on assessing the success or failure of blinding in clinical trials more generally (Lin et al., 2022; Tajika et al., 2023).

Finally, an appropriate intention-to-treat analysis should include all randomized participants, analyzed according to their assigned rather than received group, regardless of adherence (Hollis & Campbell, 1999; Detry & Lewis, 2014; Higgins et al., 2019). Per-protocol or as-treated analyses are considered inappropriate. Reviewers are directed to judge whether analytic departures, for example excluding participants who did not complete a minimum number of sessions, are likely to have had a substantial impact on the effect estimate. A 5% exclusion threshold can serve as a rough rule of thumb, though smaller exclusions may still matter substantially when outcomes are rare or exclusions are related to prognostic factors, so a case-by-case assessment of the available information remains necessary (Higgins et al., 2019).

*Domain 3. Missing outcome data*

Missing outcome data threatens the core logic of randomization, which depends on prognostic factors remaining balanced between groups after allocation. Whether missingness introduces bias depends critically on whether the reasons for missing data are related to the outcome itself, for example worsening symptoms or side effects, which raise concern, as opposed to unrelated causes, for example funding termination or technical assessment problems, which do not.

Following standard practice in current risk of bias frameworks, bias is considered unlikely when outcome data are available for at least 95% of randomized participants, with particular caution warranted for rare binary outcomes even below this threshold (Higgins et al., 2019). Where missingness exceeds this threshold, Metapsy RoB specifies which statistical approaches are considered appropriate for handling its impact. These include mixed models for repeated measures, also described as growth curve approaches using multiple assessment points, multiple imputation following current best-practice standards such as application of Rubin's combination rules, inclusion of relevant auxiliary variables, arm-specific imputation, or controlled or reference-based imputation, and sensitivity analyses across a plausible range of assumptions about the missingness mechanism that converge on consistent conclusions (Barnard & Rubin, 1999; von Hippel & Bartlett, 2021; Sullivan, White, Salter, Ryan, & Lee, 2018; Carpenter, Roger, & Kenward, 2013; Cro, Morris, Kenward, & Carpenter, 2020). Generalized estimating equations are also considered acceptable in some contexts (Little et al., 2012). Methods such as last observation carried forward imputation are explicitly considered inappropriate.

When no appropriate correction method was used or reporting is unclear, Metapsy RoB directs reviewers to three sources of indirect evidence, consistent with the general approach in current frameworks of examining indirect indicators when a direct judgment is not possible (Higgins et al., 2019). The first is whether the reported reasons for missingness suggest a relationship with the outcome's true value. The second is whether there is a large imbalance in the proportion of missing data between groups, using a difference greater than 20 percentage points as a rule-of-thumb indicator. The third is whether the reasons for missingness differ systematically between groups, for example missingness linked to severity in the intervention arm but not the control arm.

*Domain 4. Measurement of the outcome*

Current risk of bias frameworks evaluate this domain by asking whether the measurement method is appropriate and applied consistently between groups, who assesses the outcome, whether that assessor is blinded, and whether the assessment is plausibly influenced by knowledge of treatment allocation. In psychotherapy trials, appropriateness and consistency of measurement are generally straightforward to evaluate. The identity of the assessor, however, requires specific consideration, since essentially all mental health outcomes are patient-reported, whether through a self-rated questionnaire such as the Beck Depression Inventory II, or through a structured clinical interview conducted by a clinician. Clinician administered assessment offers, in principle, an additional layer of protection through masking of the assessor, since masked clinicians can interpret behavioral, facial, or verbal cues in a way that is thought to provide more objective information than unmasked assessment. In practice, this has led reviewers to treat masked clinician-rated assessments as consistently low risk, while self-reports have been rated inconsistently, ranging from low to high risk, across different reviews.

To resolve this inconsistency, an empirical comparison was conducted across psychotherapy trials for depression, examining whether self-reports and clinician-rated instruments produce systematically different effect estimates (Miguel et al., 2025; Cuijpers, Li, Hofmann, & Andersson, 2010). This work found that self-reports produced somewhat smaller effect sizes than clinician-rated instruments overall, but that this difference was not clinically meaningful when the clinician-administered assessments were themselves masked. In contrast, depression scales administered by unmasked clinicians produced substantially larger effect estimates than self-reports, with a differential standardized mean difference of 0.20. Because a fully objective benchmark for depression severity does not exist, this comparison cannot completely rule out an effect of unmasking on self-reports, but the absence of any meaningful gap between self-reports and the gold standard of masked clinician ratings, combined with the premise that participants must be aware of receiving a psychotherapeutic intervention in the first place, was taken as evidence against a general assumption of bias arising from self-report use. Accordingly, in Metapsy RoB self-report measures are, as a default, not treated as introducing risk of bias due to lack of blinding, whereas unmasked clinician-administered assessments are downgraded. Some safeguards are still required for self-reports specifically. Reviewers are asked to consider whether there is evidence, or plausible risk, of participants underreporting symptom severity, for example to please a therapist or researcher when a questionnaire is completed in that person's presence, as this could still introduce bias.

*Domain 5. Selective outcome reporting*

This domain concerns bias introduced when the results reported from a trial are chosen based on their direction, magnitude, or statistical significance, for example by selectively reporting one time point, one instrument, or one analytic approach, such as continuous versus binary, from among several that were measured or planned.

Safeguards against this bias remain underdeveloped in psychotherapy research. An in-depth examination of selective reporting across psychotherapy trials for depression found that only 21% of trials, 75 of 353, were prospectively registered, and that among the subset of prospectively registered trials in which selective reporting could be reliably examined, 17% showed evidence of selecting the reported result. Trials with evidence of selective reporting showed effect sizes 0.27 standardized mean differences larger than trials without such evidence (Miguel, Karyotaki, Cuijpers, & Cristea, 2021). Low rates of prospective registration are not unique to depression trials, having also been documented in psychotherapy research for anxiety disorders (Papola et al., 2022), obsessive-compulsive disorder (Wang et al., 2024), borderline personality disorder (Storebø et al., 2020), and post-traumatic stress disorder (Mavranezouli et al., 2020).

Consistent with current risk of bias frameworks, selection is assessed with reference to a specific outcome measurement and a specific analysis, and the appropriate level of assessment depends on the meta-analytic protocol's own approach to outcome selection (Higgins et al., 2019). In practice, reviewers are directed to check for a trial registration, protocol, or statistical analysis plan and, where available, to establish whether the outcome measures of interest, at minimum scale and time point, were specified before unblinded outcome data became available. An analysis plan should ideally also have been pre-specified, though this remains rare in practice. Where such prospective information exists, reviewers can then judge whether all pre-specified outcomes were reported and whether the reported analysis matches the plan, considering whether any apparent selection could plausibly have been influenced by the direction, magnitude, or significance of the results, and whether unreported outcome data might still be obtainable through other means such as contacting study authors. Because assessment is carried out at the outcome level, evidence of selective reporting for one outcome domain, for example omission of a pre-specified anxiety outcome, does not automatically imply high risk of bias for a different outcome domain, for example depression, if the omitted outcome falls outside the scope of the review in question.

*5. Design choices, scope, and boundaries*

The certainty of evidence synthesized in clinical guidelines, for example through GRADE assessment, depends in part on the risk of bias evaluated in the underlying systematic reviews (Guyatt et al., 2008). Inconsistent standards for risk of bias assessment across reviews can therefore translate directly into inconsistent or misleading guideline conclusions. Structured, context-specific implementation tools have been proposed as a way to improve both the efficiency and the consistency of risk of bias assessment (Minozzi, Dwan, Borrelli, & Filippini, 2022; Minozzi et al., 2020), and this was the central motivation for developing Metapsy RoB as a standardized tool rather than relying on case-by-case judgment alone. This motivation, in turn, shaped several specific design choices, described below, together with the scope within which the tool is intended to be applied and the issues it deliberately does not address.

*5.1 Standardization across many reviewers and a large number of trials*

Current guidance generally favors avoiding fixed cut-offs in favor of individualized, trial-by-trial judgment. Metapsy RoB nonetheless introduced numerical thresholds for several items, for example 95% data availability for Domain 3, a standardized mean difference above 0.2 for baseline severity imbalance in Domain 1, and a difference greater than 20 percentage points between groups as an indirect indicator of attrition bias in Domain 3. This departure from a purely individualized approach was considered a necessary and justifiable compromise given the scale of the intended application, a living database of over 1,000 trials rated by multiple reviewers, where predefined cut-offs support inter-reviewer agreement and enable the accompanying digital assistant tools to compute scores automatically. For the same reason, all 24 items were formulated so that Yes or Probably Yes responses consistently indicate lower risk of bias, which eases both manual and automated score calculation.

*5.2 Scope of application*

Metapsy RoB was developed with a primary focus on individually randomized, parallel-group trials assessing the intention-to-treat effect, as these represent the most common design in the psychotherapy literature. Most items remain applicable to cluster randomized trials with the addition of design-specific signalling questions, but cluster and crossover designs raise further considerations not fully addressed by this tool. Reviewers interested in the effect of adhering to intervention rather than the intention-to-treat effect should consult the corresponding guidance for Domain 2 in current risk of bias frameworks directly (Higgins et al., 2019).

*5.3 Issues outside the scope of risk of bias assessment*

Metapsy RoB does not address other well-documented problems in psychotherapy trial design that fall outside the scope of risk of bias assessment as conventionally defined. These include the use of weak control conditions such as waitlist, imprecise definitions of treatment as usual, researcher allegiance effects, small sample sizes, inadequate assessment of adverse events, and poor reporting more broadly (Cuijpers, 2025; Cuijpers, 2019; Cuijpers & Cristea, 2015; Watts, Turnell, Kladnitski, Newby, & Andrews, 2015; Cuijpers, Quero, Papola, Cristea, & Karyotaki, 2021). Aspects of treatment implementation quality, such as therapist training and supervision, manualization, and treatment fidelity, are similarly relevant to evaluating the effect of adhering to an intervention but were not incorporated into this intention-to-treat-focused tool.

*5.4 Remaining evidence gaps*

During the consensus process through which the 24 items and their guidance were developed, discrepancies between reviewers on the trials used for piloting were resolved through group discussion rather than formally tracked, since this developmental phase was not originally conceived as a preplanned methodological study. This is distinct from the inter-rater reliability of the ratings produced by applying the finished tool, which was formally assessed in the present study across a subsample of 237 trials and is reported in the main text. Separately, some domains, particularly deviations from intended interventions, remain limited by a broader lack of empirical research within psychotherapy trials to inform decision rules, a gap that itself partly reflects poor reporting in primary trials. Much of the meta-epidemiological evidence available to inform the tool's development derives specifically from depression trials, although the tool was not intended to be limited to this condition.

*References*

Barnard, J., & Rubin, D. (1999). Miscellanea. Small-sample degrees of freedom with multiple imputation. Biometrika, 86(4), 948-955. doi:10.1093/biomet/86.4.948

Baskin, T. W., Tierney, S. C., Minami, T., & Wampold, B. E. (2003). Establishing specificity in psychotherapy. A meta-analysis of structural equivalence of placebo controls. Journal of Consulting and Clinical Psychology, 71(6), 973-979. doi:10.1037/0022-006X.71.6.973

Bower, P., Kontopantelis, E., Sutton, A., Kendrick, T., Richards, D. A., Gilbody, S., . . . Liu, E. T. (2013). Influence of initial severity of depression on effectiveness of low intensity interventions. Meta-analysis of individual patient data. Bmj, 346, f540. doi:10.1136/bmj.f540

Carpenter, J. R., Roger, J. H., & Kenward, M. G. (2013). Analysis of longitudinal trials with protocol deviation. A framework for relevant, accessible assumptions, and inference via multiple imputation. J Biopharm Stat, 23(6), 1352-1371. doi:10.1080/10543406.2013.834911

Cro, S., Morris, T. P., Kenward, M. G., & Carpenter, J. R. (2020). Sensitivity analysis for clinical trials with missing continuous outcome data using controlled multiple imputation. A practical guide. Statistics in Medicine, 39(21), 2815-2842. doi:10.1002/sim.8569

Crocker, T. F., Lam, N., Jordão, M., Brundle, C., Prescott, M., Forster, A., . . . Clegg, A. (2023). Risk of bias assessment using Cochrane's revised tool for randomized trials (RoB 2) was useful but challenging and resource intensive. Observations from a systematic review. Journal of Clinical Epidemiology, 161, 39-45. doi:10.1016/j.jclinepi.2023.06.015

Cuijpers, P. (2019). Targets and outcomes of psychotherapies for mental disorders. An overview. World Psychiatry, 18(3), 276-285. doi:10.1002/wps.20661

Cuijpers, P. (2025). Has the time come to stop using control groups in trials of psychosocial interventions? World Psychiatry.

Cuijpers, P., & Cristea, I. A. (2015). How to prove that your therapy is effective, even when it is not. A guideline. Epidemiology and Psychiatric Sciences, 25(5), 428-435. doi:10.1017/S2045796015000864

Cuijpers, P., Ciharova, M., Quero, S., Miguel, C., Driessen, E., Harrer, M., . . . Karyotaki, E. (2022). The contribution of individual participant data meta-analyses of psychotherapies for depression to the development of personalized treatments. A systematic review. J Pers Med, 12(1). doi:10.3390/jpm12010093

Cuijpers, P., Harrer, M., Miguel, C., Ciharova, M., & Karyotaki, E. (2023). Five decades of research on psychological treatments of depression. A historical and meta-analytic overview. American Psychologist. doi:10.1037/amp0001250

Cuijpers, P., Li, J., Hofmann, S. G., & Andersson, G. (2010). Self-reported versus clinician-rated symptoms of depression as outcome measures in psychotherapy research on depression. A meta-analysis. Clin Psychol Rev, 30(6), 768-778. doi:10.1016/j.cpr.2010.06.001

Cuijpers, P., Miguel, C., Harrer, M., Ciharova, M., & Karyotaki, E. (2024). The overestimation of the effect sizes of psychotherapies for depression in waitlist controlled trials. A meta-analytic comparison with usual care controlled trials. Epidemiology and Psychiatric Sciences, 33, e56.

Cuijpers, P., Quero, S., Papola, D., Cristea, I. A., & Karyotaki, E. (2021). Care as usual control groups across different settings in randomized trials on psychotherapy for adult depression. A meta-analysis. Psychol Med, 51(4), 634-644. doi:10.1017/s0033291719003581

de Haan, A., Meiser-Stedman, R., Landolt, M. A., Kuhn, I., Black, M. J., Klaus, K., . . . Dalgleish, T. (2024). Efficacy and moderators of efficacy of cognitive behavioural therapies with a trauma focus in children and adolescents. An individual participant data meta-analysis of randomised trials. Lancet Child Adolesc Health, 8(1), 28-39. doi:10.1016/s2352-4642(23)00253-5

Detry, M. A., & Lewis, R. J. (2014). The intention to treat principle. How to assess the true effect of choosing a medical treatment. Jama, 312(1), 85-86.

Doering, B. K., Glombiewski, J. A., & Rief, W. (2018). Chapter Fourteen. Expectation-focused psychotherapy to improve clinical outcomes. In L. Colloca (Ed.), International Review of Neurobiology (Vol. 138, pp. 257-270). Academic Press.

Driessen, E., Cuijpers, P., Hollon, S. D., & Dekker, J. J. (2010). Does pretreatment severity moderate the efficacy of psychological treatment of adult outpatient depression? A meta-analysis. J Consult Clin Psychol, 78(5), 668-680. doi:10.1037/a0020570

Furukawa, T. A., Efthimiou, O., Weitz, E. S., Cipriani, A., Keller, M. B., Kocsis, J. H., . . . Schramm, E. (2018). Cognitive behavioral analysis system of psychotherapy, drug, or their combination for persistent depressive disorder. Personalizing the treatment choice using individual participant data network metaregression. Psychother Psychosom, 87(3), 140-153. doi:10.1159/000489227

Furukawa, T. A., Noma, H., Caldwell, D. M., Honyashiki, M., Shinohara, K., Imai, H., . . . Churchill, R. (2014). Waiting list may be a nocebo condition in psychotherapy trials. A contribution from network meta-analysis. Acta Psychiatr Scand, 130(3), 181-192. doi:10.1111/acps.12275

Furukawa, T. A., Suganuma, A., Ostinelli, E. G., Andersson, G., Beevers, C. G., Shumake, J., . . . Cuijpers, P. (2021). Dismantling, optimising, and personalising internet cognitive behavioural therapy for depression. A systematic review and component network meta-analysis using individual participant data. Lancet Psychiatry, 8(6), 500-511. doi:10.1016/s2215-0366(21)00077-8

Group, E.-B. M. W. (1992). Evidence based medicine. A new approach to teaching the practice of medicine. Jama, 268(17), 2420-2425. doi:10.1001/jama.1992.03490170092032

Guyatt, G. H., Oxman, A. D., Vist, G. E., Kunz, R., Falck-Ytter, Y., Alonso-Coello, P., & Schünemann, H. J. (2008). GRADE. An emerging consensus on rating quality of evidence and strength of recommendations. Bmj, 336(7650), 924-926. doi:10.1136/bmj.39489.470347.AD

Higgins, Savović, Page, Sterne, & Group, T. R. D. (2019). Revised Cochrane risk of bias tool for randomized trials (RoB 2). Retrieved from https://sites.google.com/site/riskofbiastool/welcome/rob-2-0-tool/current-version-of-rob-2

Higgins, J. P., Savović, J., Page, M. J., Elbers, R. G., & Sterne, J. A. (2019). Assessing risk of bias in a randomized trial. Cochrane Handbook for Systematic Reviews of Interventions, 205-228.

Higgins, J. P. T., Thomas, J., Chandler, J., Cumpston, M., Li, T., Page, M. J., & Welch, V. A. (2019). Cochrane Handbook for Systematic Reviews of Interventions (Second ed.). Chichester (UK). John Wiley & Sons.

Hollis, S., & Campbell, F. (1999). What is meant by intention to treat analysis? Survey of published randomised controlled trials. Bmj, 319(7211), 670-674. doi:10.1136/bmj.319.7211.670

Jørgensen, L., Paludan-Müller, A. S., Laursen, D. R., Savović, J., Boutron, I., Sterne, J. A., . . . Hróbjartsson, A. (2016). Evaluation of the Cochrane tool for assessing risk of bias in randomized clinical trials. Overview of published comments and analysis of user practice in Cochrane and non-Cochrane reviews. Systematic Reviews, 5, 1-13.

Karyotaki, E., Ebert, D. D., Donkin, L., Riper, H., Twisk, J., Burger, S., . . . Cuijpers, P. (2018). Do guided internet-based interventions result in clinically relevant changes for patients with depression? An individual participant data meta-analysis. Clin Psychol Rev, 63, 80-92. doi:10.1016/j.cpr.2018.06.007

Kuyken, W., Warren, F. C., Taylor, R. S., Whalley, B., Crane, C., Bondolfi, G., . . . Dalgleish, T. (2016). Efficacy of mindfulness based cognitive therapy in prevention of depressive relapse. An individual patient data meta-analysis from randomized trials. JAMA Psychiatry, 73(6), 565-574. doi:10.1001/jamapsychiatry.2016.0076

Lin, Y.-H., Sahker, E., Shinohara, K., Horinouchi, N., Ito, M., Lelliott, M., . . . Furukawa, T. A. (2022). Assessment of blinding in randomized controlled trials of antidepressants for depressive disorders 2000 to 2020. A systematic review and meta-analysis. eClinicalMedicine, 50. doi:10.1016/j.eclinm.2022.101505

Little, R. J., D'Agostino, R., Cohen, M. L., Dickersin, K., Emerson, S. S., Farrar, J. T., . . . Stern, H. (2012). The prevention and treatment of missing data in clinical trials. N Engl J Med, 367(14), 1355-1360. doi:10.1056/NEJMsr1203730

Mavranezouli, I., Megnin-Viggars, O., Daly, C., Dias, S., Welton, N. J., Stockton, S., . . . Pilling, S. (2020). Psychological treatments for post-traumatic stress disorder in adults. A network meta-analysis. Psychological Medicine, 50(4), 542-555. doi:10.1017/S0033291720000070

Michopoulos, I., Furukawa, T. A., Noma, H., Kishimoto, S., Onishi, A., Ostinelli, E. G., . . . Cuijpers, P. (2021). Different control conditions can produce different effect estimates in psychotherapy trials for depression. J Clin Epidemiol, 132, 59-70. doi:10.1016/j.jclinepi.2020.12.012

Miguel, C., Harrer, M., Karyotaki, E., Plessen, C. Y., Ciharova, M., Furukawa, T. A., . . . Cuijpers, P. (2025). Self reports vs clinician ratings of efficacies of psychotherapies for depression. A meta-analysis of randomized trials. Epidemiology and Psychiatric Sciences, 34, e15. doi:10.1017/S2045796025000095

Miguel, C., Karyotaki, E., Cuijpers, P., & Cristea, I. A. (2021). Selective outcome reporting and the effectiveness of psychotherapies for depression. World Psychiatry, 20(3), 444-445. doi:10.1002/wps.20900

Minozzi, S., Cinquini, M., Gianola, S., Gonzalez-Lorenzo, M., & Banzi, R. (2020). The revised Cochrane risk of bias tool for randomized trials (RoB 2) showed low interrater reliability and challenges in its application. Journal of Clinical Epidemiology, 126, 37-44. doi:10.1016/j.jclinepi.2020.06.015

Minozzi, S., Dwan, K., Borrelli, F., & Filippini, G. (2022). Reliability of the revised Cochrane risk of bias tool for randomised trials (RoB2) improved with the use of implementation instruction. Journal of Clinical Epidemiology, 141, 99-105.

Munder, T., & Barth, J. (2018). Cochrane's risk of bias tool in the context of psychotherapy outcome research. Psychother Res, 28(3), 347-355. doi:10.1080/10503307.2017.1411628

Papola, D., Ostuzzi, G., Tedeschi, F., Gastaldon, C., Purgato, M., Del Giovane, C., . . . Barbui, C. (2022). Comparative efficacy and acceptability of psychotherapies for panic disorder with or without agoraphobia. Systematic review and network meta-analysis of randomised controlled trials. The British Journal of Psychiatry, 221(3), 507-519. doi:10.1192/bjp.2021.148

Reins, J. A., Buntrock, C., Zimmermann, J., Grund, S., Harrer, M., Lehr, D., . . . Ebert, D. D. (2021). Efficacy and moderators of internet-based interventions in adults with subthreshold depression. An individual participant data meta-analysis of randomized controlled trials. Psychother Psychosom, 90(2), 94-106. doi:10.1159/000507819

Rosenfield, D., Smits, J. A. J., Hofmann, S. G., Mataix-Cols, D., de la Cruz, L. F., Andersson, E., . . . Otto, M. W. (2019). Changes in dosing and dose timing of D-cycloserine explain its apparent declining efficacy for augmenting exposure therapy for anxiety related disorders. An individual participant data meta-analysis. Journal of Anxiety Disorders, 68, 102149. doi:10.1016/j.janxdis.2019.102149

Scholten, W., Seldenrijk, A., Hoogendoorn, A., Bosman, R., Muntingh, A., Karyotaki, E., . . . Batelaan, N. (2023). Baseline severity as a moderator of the waiting list controlled association of cognitive behavioral therapy with symptom change in social anxiety disorder. A systematic review and individual patient data meta-analysis. JAMA Psychiatry, 80(8), 822-831. doi:10.1001/jamapsychiatry.2023.1291

Steinberg, E., Greenfield, S., Wolman, D. M., Mancher, M., & Graham, R. (2011). Clinical practice guidelines we can trust. National Academies Press.

Sterne, J. A. C., Savović, J., Page, M. J., Elbers, R. G., Blencowe, N. S., Boutron, I., . . . Higgins, J. P. T. (2019). RoB 2. A revised tool for assessing risk of bias in randomised trials. Bmj, 366, l4898. doi:10.1136/bmj.l4898

Storebø, O. J., Stoffers-Winterling, J. M., Völlm, B. A., Kongerslev, M. T., Mattivi, J. T., Jørgensen, M. S., . . . Simonsen, E. (2020). Psychological therapies for people with borderline personality disorder. Cochrane Database Syst Rev, 5(5), Cd012955. doi:10.1002/14651858.CD012955.pub2

Sullivan, T. R., White, I. R., Salter, A. B., Ryan, P., & Lee, K. J. (2018). Should multiple imputation be the method of choice for handling missing data in randomized trials? Stat Methods Med Res, 27(9), 2610-2626. doi: 10.1177/0962280216683570

Tajika, A., Furukawa, T. A., Shinohara, K., Kikuchi, S., Toyomoto, R., Furukawa, Y., . . . Leucht, S. (2023). Blinding successfulness in antipsychotic trials of acute treatment for schizophrenia. A systematic review. BMJ Ment Health, 26(1). doi:10.1136/bmjment-2023-300654

Tomlinson, E., Cooper, C., Davenport, C., Rutjes, A. W. S., Leeflang, M., Mallett, S., & Whiting, P. (2024). Common challenges and suggestions for risk of bias tool development. A systematic review of methodological studies. Journal of Clinical Epidemiology, 171. doi:10.1016/j.jclinepi.2024.111370

von Hippel, P. T., & Bartlett, J. W. (2021). Maximum likelihood multiple imputation. Faster imputations and consistent standard errors without posterior draws. Statistical Science, 36(3), 400-420, 421. doi:10.1214/20-STS793

Wampold, B. E., & Imel, Z. E. (2015). The great psychotherapy debate. The evidence for what makes psychotherapy work (2nd ed.). Routledge.

Wang, Y., Miguel, C., Ciharova, M., Amarnath, A., Lin, J., Zhao, R., . . . Cuijpers, P. (2024). The effectiveness of psychological treatments for obsessive compulsive disorders. A meta-analysis of randomized controlled trials published over last 30 years. Psychological Medicine, 54(11), 2838-2851. doi:10.1017/S0033291724001375

Watts, S. E., Turnell, A., Kladnitski, N., Newby, J. M., & Andrews, G. (2015). Treatment as usual (TAU) is anything but usual. A meta-analysis of CBT versus TAU for anxiety and depression. Journal of Affective Disorders, 175, 152-167. doi:10.1016/j.jad.2014.12.025

Wright, S. L., Karyotaki, E., Cuijpers, P., Bisson, J., Papola, D., Witteveen, A., . . . Sijbrandij, M. (2024). EMDR v. other psychological therapies for PTSD. A systematic review and individual participant data meta-analysis. Psychological Medicine, 54(8), 1580-1588. doi:10.1017/S0033291723003446

#### Supplement E. Inter-rater reliability

A sample of 237 studies was rated independently by two raters across 10 rater pairs (n = 7–35 studies per pair). Prior to independent rating, all raters participated in a training session and calibration exercise consisting of a review of the risk of bias rating manual and group discussion of example studies to ensure a shared understanding of rating criteria. Following independent ratings, discrepancies were discussed and resolved by consensus before final ratings were assigned. The inter-rater reliability estimates reported here reflect agreement between raters prior to consensus discussion. It should be noted that some raters participating in the reliability subsample were involved in developing the risk of bias rating manual and had extensive prior experience with the rating criteria (CM, TF, ES, MS, EK, PC). For pairs where both raters had this level of experience, reliability estimates may reflect above-average familiarity with the instrument rather than what would be expected in typical applications. However, most pairs were mixed in terms of experience, which limits this concern, and four pairs (AS & PK, LT & EsK, RT & AT, RT & KY) consisted entirely of raters not involved in manual development. Reliability estimates for these four pairs were strong and broadly comparable to those of pairs with greater prior experience, suggesting that the rating instrument can yield reliable results following adequate training.

The sensitivity analysis restricting to the 16 manually rated items yielded somewhat lower estimates (kappa: 0.55–0.90; observed agreement: 77.6%–95.1%), reflecting the exclusion of seven pre-filled items identical across raters by construction. Score-level Gwet's AC2 varied more across pairs, particularly for Domain 2 (0.30–1.00) and Domain 3 (0.23–1.00), and for pairs with smaller sample sizes. Pair-level estimates are reported in Tables E1 and E2.

**Table E1**

*Pair-level item-level inter-rater reliability: Cohen's kappa and observed agreement on individual RoB items*

| Rater pair | n studies | Cohen's kappa (all items) | Observed agreement – all items (%) | Cohen's kappa (manual items only) | Observed agreement – manual items (%) |
|---|---|---|---|---|---|
| CM & TF | 32 | 0.82 [0.78, 0.87] | 91.3 | 0.71 [0.63, 0.78] | 85.8 |
| CM & EsK | 7 | 0.92 [0.85, 0.98] | 95.8 | 0.87 [0.77, 0.97] | 93.6 |
| AS & PK | 33 | 0.92 [0.89, 0.95] | 96.1 | 0.90 [0.85, 0.94] | 95.1 |
| EK & OP | 33 | 0.87 [0.83, 0.91] | 93.6 | 0.79 [0.73, 0.85] | 89.6 |
| ES & MS | 24 | 0.81 [0.75, 0.86] | 90.3 | 0.73 [0.65, 0.81] | 86.4 |
| LT & EsK | 30 | 0.82 [0.77, 0.87] | 90.8 | 0.73 [0.66, 0.80] | 86.6 |
| PC & YW | 35 | 0.83 [0.78, 0.87] | 91.6 | 0.67 [0.59, 0.76] | 85.8 |
| RT & AT | 7 | 0.73 [0.61, 0.85] | 86.4 | 0.55 [0.36, 0.74] | 77.6 |
| RT & KY | 14 | 0.93 [0.88, 0.97] | 96.4 | 0.87 [0.79, 0.95] | 94.0 |
| CM & MC | 22 | 0.92 [0.88, 0.96] | 95.9 | 0.86 [0.79, 0.93] | 93.8 |

**Table E2**

*Pair-level score-level inter-rater reliability: Gwet's AC2 with 95% confidence intervals and observed agreement by RoB domain and overall score*

| Domain | Rater pair | n studies | AC2 [95% CI] | Observed agreement (%) |
|---|---|---|---|---|
| Overall RoB score | AS & PK | 33 | 0.87 [0.73, 1.00] | 83.3 |
| Overall RoB score | CM & EsK | 7 | 0.79 [0.23, 1.00] | 85.7 |
| Overall RoB score | CM & MC | 22 | 0.86 [0.70, 1.00] | 77.8 |
| Overall RoB score | CM & TF | 32 | 0.82 [0.65, 0.99] | 73.1 |

| | | | | |
|---|---|---|---|---|
| Overall RoB score | EK & OP | 33 | 0.60 [0.35, 0.86] | 56.0 |
| Overall RoB score | ES & MS | 24 | 0.64 [0.37, 0.92] | 60.9 |
| Overall RoB score | LT & EsK | 30 | 0.88 [0.74, 1.00] | 82.8 |
| Overall RoB score | PC & YW | 35 | 0.72 [0.55, 0.90] | 58.6 |
| Overall RoB score | RT & AT | 7 | 0.70 [0.09, 1.00] | 60.0 |
| Overall RoB score | RT & KY | 14 | 0.89 [0.73, 1.00] | 84.6 |
| Domain 1 - Randomization | AS & PK | 33 | 0.92 [0.84, 1.00] | 87.9 |
| Domain 1 - Randomization | CM & EsK | 7 | 1.00 [1.00, 1.00] | 100.0 |
| Domain 1 - Randomization | CM & MC | 22 | 0.88 [0.76, 1.00] | 81.8 |
| Domain 1 - Randomization | CM & TF | 32 | 0.78 [0.59, 0.96] | 75.0 |
| Domain 1 - Randomization | EK & OP | 33 | 0.92 [0.83, 1.00] | 87.9 |
| Domain 1 - Randomization | ES & MS | 24 | 0.73 [0.51, 0.96] | 66.7 |
| Domain 1 - Randomization | LT & EsK | 30 | 0.93 [0.85, 1.00] | 90.0 |
| Domain 1 - Randomization | PC & YW | 35 | 0.83 [0.67, 0.99] | 82.9 |
| Domain 1 - Randomization | RT & AT | 7 | 0.91 [0.69, 1.00] | 85.7 |
| Domain 1 - Randomization | RT & KY | 14 | 0.91 [0.78, 1.00] | 85.7 |
| Domain 2 - Deviations | AS & PK | 33 | 0.81 [0.62, 1.00] | 84.8 |
| Domain 2 - Deviations | CM & EsK | 7 | 1.00 [1.00, 1.00] | 100.0 |
| Domain 2 - Deviations | CM & MC | 22 | 0.86 [0.62, 1.00] | 90.0 |
| Domain 2 - Deviations | CM & TF | 32 | 0.53 [0.20, 0.85] | 62.1 |
| Domain 2 - Deviations | EK & OP | 33 | 0.92 [0.80, 1.00] | 93.3 |
| Domain 2 - Deviations | ES & MS | 24 | 0.57 [0.20, 0.95] | 65.2 |
| Domain 2 - Deviations | LT & EsK | 30 | 0.61 [0.31, 0.91] | 73.3 |
| Domain 2 - Deviations | PC & YW | 35 | 0.30 [−0.02, 0.61] | 62.9 |
| Domain 2 - Deviations | RT & AT | 7 | 0.92 [0.66, 1.00] | 83.3 |
| Domain 2 - Deviations | RT & KY | 14 | 0.74 [0.34, 1.00] | 84.6 |
| Domain 3 - Missing data | AS & PK | 33 | 0.94 [0.87, 1.00] | 93.5 |
| Domain 3 - Missing data | CM & EsK | 7 | 1.00 [1.00, 1.00] | 100.0 |
| Domain 3 - Missing data | CM & MC | 22 | 0.74 [0.36, 1.00] | 90.0 |
| Domain 3 - Missing data | CM & TF | 32 | 0.52 [0.18, 0.86] | 69.0 |
| Domain 3 - Missing data | EK & OP | 33 | 0.57 [0.29, 0.84] | 59.4 |
| Domain 3 - Missing data | ES & MS | 24 | 0.71 [0.41, 1.00] | 79.2 |
| Domain 3 - Missing data | LT & EsK | 30 | 0.80 [0.59, 1.00] | 83.3 |
| Domain 3 - Missing data | PC & YW | 35 | 0.23 [−0.18, 0.64] | 53.1 |
| Domain 3 - Missing data | RT & AT | 7 | 0.32 [−0.69, 1.00] | 57.1 |
| Domain 3 - Missing data | RT & KY | 14 | 0.94 [0.80, 1.00] | 92.9 |
| Domain 4 - Measurement | AS & PK | 33 | 0.92 [0.79, 1.00] | 93.8 |
| Domain 4 - Measurement | CM & EsK | 7 | 1.00 [1.00, 1.00] | 100.0 |
| Domain 4 - Measurement | CM & MC | 22 | 1.00 [1.00, 1.00] | 100.0 |
| Domain 4 - Measurement | CM & TF | 32 | 1.00 [1.00, 1.00] | 100.0 |
| Domain 4 - Measurement | EK & OP | 33 | 0.96 [0.89, 1.00] | 97.0 |
| Domain 4 - Measurement | ES & MS | 24 | 0.95 [0.85, 1.00] | 95.8 |
| Domain 4 - Measurement | LT & EsK | 30 | 0.96 [0.88, 1.00] | 96.7 |
| Domain 4 - Measurement | PC & YW | 35 | 0.97 [0.90, 1.00] | 97.1 |
| Domain 4 - Measurement | RT & AT | 7 | 0.55 [−0.40, 1.00] | 66.7 |
| Domain 4 - Measurement | RT & KY | 14 | 1.00 [1.00, 1.00] | 100.0 |
| Domain 5 – Selective reporting | AS & PK | 33 | 0.96 [0.91, 1.00] | 90.9 |
| Domain 5 – Selective reporting | CM & EsK | 7 | 0.62 [−0.30, 1.00] | 85.7 |
| Domain 5 – Selective reporting | CM & MC | 22 | 0.98 [0.94, 1.00] | 95.5 |
| Domain 5 – Selective reporting | CM & TF | 32 | 0.96 [0.92, 1.00] | 90.6 |

| | | | | |
|---|---|---|---|---|
| Domain 5 – Selective reporting | EK & OP | 33 | 0.93 [0.86, 1.00] | 86.2 |
| Domain 5 – Selective reporting | ES & MS | 24 | 0.86 [0.75, 0.98] | 75.0 |
| Domain 5 – Selective reporting | LT & EsK | 30 | 0.76 [0.50, 1.00] | 79.3 |
| Domain 5 – Selective reporting | PC & YW | 35 | 0.97 [0.93, 1.00] | 90.9 |
| Domain 5 – Selective reporting | RT & AT | 7 | 0.83 [0.51, 1.00] | 71.4 |
| Domain 5 – Selective reporting | RT & KY | 14 | 0.94 [0.84, 1.00] | 85.7 |

Notes on Tables E1 and E2:
a) Two rater pairs (CM & EsK; RT & AT) rated only 7 studies each, resulting in wide confidence intervals and unstable point estimates. These pairs are included in all summary statistics for completeness but their estimates should be interpreted cautiously.
b) Negative lower confidence interval bounds for Gwet's AC2 are possible when sample sizes are small and observed agreement is moderate. They do not indicate negative reliability, but rather reflect high estimation uncertainty.

#### Supplement F. Deviations from Preregistered Analysis Plan

*Inter-rater Reliability*

The preregistered analysis plan (Miguel et al., 2026) specified that inter-rater reliability at the score level would be estimated using Cohen's weighted kappa with linear weights (cohen.kappa function, psych package; Revelle, 2026). This was replaced with Gwet's AC2 with ordinal weights (irrCAC package; Gwet, 2026). This decision was made after inspecting the score distributions, prior to interpreting any reliability estimates. For several domains, particularly Domain 4 (measurement of the outcome) and Domain 5 (selective outcome reporting), the large majority of trials received the same rating category, resulting in near-zero variance in the score distributions. Under these conditions, kappa-family coefficients produce paradoxical estimates: weighted kappa can equal zero or take negative values despite high observed agreement, because the chance-correction term becomes unstable when marginal distributions are highly skewed (Feinstein & Cicchetti, 1990; Cicchetti & Feinstein, 1990).

Gwet's AC2 uses a different estimator of expected agreement that remains stable and interpretable under skewed distributions (Gwet, 2014), and has been used for this purpose in comparable risk-of-bias reliability studies (Vidor et al., 2026). Ordinal weights were applied, consistent with the intent of the original preregistered approach. Observed percentage agreement is additionally reported for each domain.

The preregistered protocol specified an inter-rater reliability subsample of 250 studies. The final subsample comprised 237 studies, a reduction of 13. Three studies were excluded because they were identified as cluster randomized controlled trials after IRR ratings had been completed, and five studies were excluded because they were not included in the final Metapsy depression database at the time of analysis due to not meeting eligibility criteria (e.g., not an RCT, effect size cannot be calculated, pdf not available, etc.). A further five studies had been assigned to rater pairs but ratings were not available in the final data file and could not be recovered. These exclusions are not expected to meaningfully affect the reliability estimates.

*Sensitivity analysis for trends objective*

The preregistered analysis plan specified that all meta-epidemiological analyses (objectives 2 to 5) would be repeated restricting the sample to trials published from 2001 onwards, the year the revised CONSORT statement was published (Moher et al., 2001). This sensitivity analysis was conducted for objectives 2, 4, and 5, but not for objective 3 (trends in RoB over time). Restricting the trends analysis to post-2001 trials would remove the baseline period necessary to assess change over time, which would make the analysis uninformative. Results for objective 3 are therefore based on the full sample (N = 514).

*References*

Cicchetti, D. V., & Feinstein, A. R. (1990). High agreement but low kappa: II. Resolving the paradoxes. *Journal of Clinical Epidemiology*, *43*(6), 551–558. https://doi.org/10.1016/0895-4356(90)90159-M

Feinstein, A. R., & Cicchetti, D. V. (1990). High agreement but low kappa: I. The problems of two paradoxes. *Journal of Clinical Epidemiology*, *43*(6), 543–549. https://doi.org/10.1016/0895-4356(90)90158-l

Gwet, K. L. (2014). *Handbook of inter-rater reliability: The definitive guide to measuring the extent of agreement among raters (4th ed.)*. Advanced Analytics, LLC.

Gwet, K. L. (2026). *irrCAC: Computing the Extent of Agreement among Raters with Chance-Corrected Agreement Coefficient (CAC)* (Version 1.4) [Computer software]. https://CRAN.R-project.org/package=irrCAC

Miguel, C., Harrer, M., Sahker, E., Sakata, M., Karyotaki, E., Furukawa, T. A., & Cuijpers, P. (2026, June 3). *Risk of bias in psychotherapy trials for depression: An empirical examination on a complete cohort of RCTs.* https://doi.org/10.17605/OSF.IO/3BSX8

Moher, D., Schulz, K. F., & Altman, D. G. (2001). The CONSORT statement: Revised recommendations for improving the quality of reports of parallel-group randomised trials. *The Lancet*, *357*(9263), 1191–1194. https://doi.org/10.1016/S0140-6736(00)04337-3

Revelle, W. (2026). *psych: Procedures for Psychological, Psychometric, and Personality Research* (Version 2.6.4) [Computer software]. https://CRAN.R-project.org/package=psych

Vidor, P. R., Casiraghi, Y., de Souza, A. M., & Schmidt, M. I. (2026). Assessing the risk of bias of clinical trials with large language models and ROBUST-RCT: A feasibility study. *Scientific Reports*, *16*(1), 13723. https://doi.org/10.1038/s41598-026-44303-z

#### Supplement G. PRISMA flowchart and references of included studies

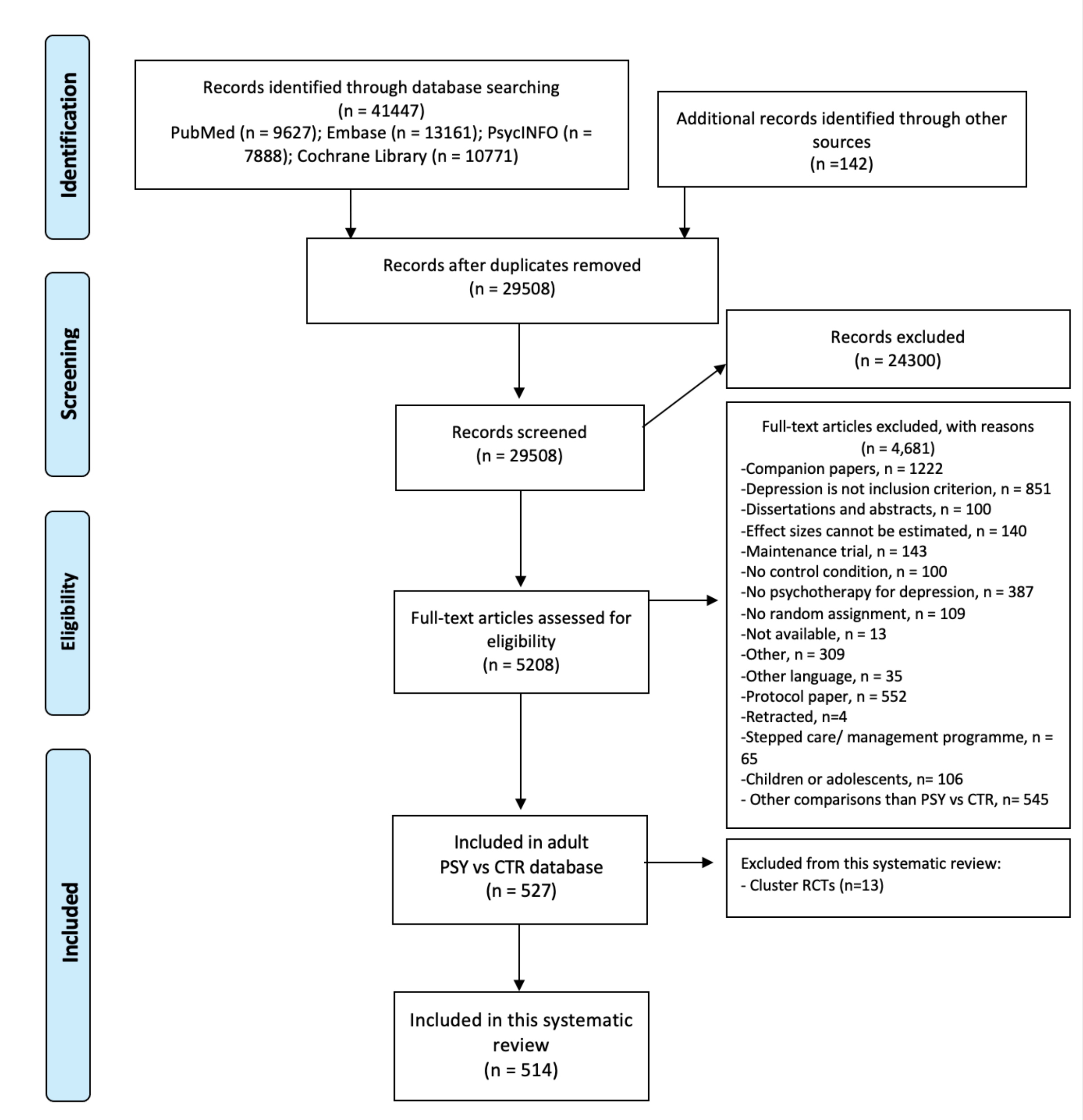

**Figure G1.** PRISMA flowchart (searches up to 1st January 2026)

##### *Reference list of included studies*

Aagaard, J., Foldager, L., Makki, A., Hansen, V., & Møller-Nielsen, K. (2017). The efficacy of psychoeducation on recurrent depression: a randomized trial with a 2-year follow-up. Nord J Psychiatry, 71(3), 223-229.

Abas M, Nyamayaro P, Bere T, et al. Feasibility and acceptability of a task-shifted intervention to enhance adherence to HIV medication and improve depression in people living with HIV in Zimbabwe, a low income country in sub-Saharan Africa. AIDS and Behavior 2018; 22(1): 86-101.

Abbas, Q., Latif, S., Ayaz Habib, H., Shahzad, S., Sarwar, U., Shahzadi, M., . . . Washdev, W. (2023). Cognitive behavior therapy for diabetes distress, depression, health anxiety, quality of life and treatment adherence among patients with Type-II diabetes mellitus: A randomized control trial. BMC Psychiatry, 23. doi:10.1186/s12888-023-04546-w

Abbas, Q., Nisa, M., Khan, M. U., Anwar, N., Aljhani, S., Ramzan, Z., & Shahzadi, M. (2023). Brief cognitive behavior therapy for stigmatization, depression, quality of life, social support and adherence to treatment among patients with HIV/AIDS: a randomized control trial. BMC Psychiatry, 23(1), 539.

Abujilban, S., Al-Omari, H., Issa, E., Alhamdan, A., Al-Nabulsi, L., Mrayan, L., . . . Kernohan, W. G. (2024). Effectiveness of Telephone-Based Interpersonal Psychotherapy on Antenatal Depressive Symptoms: A Prospective Randomized Controlled Trial in The Kingdom of Jordan. Journal of the American Psychiatric Nurses Association, 30(3), 635-645.

Afonso, R., & Bueno, B. (2009). Efectos de un programa de reminiscencia sobre la sintomatología depresiva en una muestra de población mayor portuguesa. Revista Española de Geriatría y Gerontología, 44(6), 317-322.

Aguilar-Raab C, Winter F, Warth M, Stoffel M, Moessner M, Hernández C, et al. A compassion-based treatment for couples with the female partner suffering from current depressive disorder: A randomized-controlled trial. Journal of Affective Disorders. 2023;342:127-38.

Ahmadpanah, M., Paghale, S. J., Bakhtyari, A., Kaikhavani, S., Aghaei, E., Nazaribadie, M., . . . Brand, S. (2016). Effects of psychotherapy in combination with pharmacotherapy, when compared to pharmacotherapy only on blood pressure, depression, and anxiety in female patients with hypertension.

Ahmed N, Zavala GA, Siddiqui F, Aslam F, Keding A, Halmkan S, et al. A randomised controlled feasibility trial of Behavioural activation as a treatment for people with diabetes and depression: (DiaDeM feasibility trial). J Affect Disord. 2025;372:333-46.

Alexopoulos GS, Raue PJ, McCulloch C, Kanellopoulos D, Seirup JK, Sirey JA, et al. Clinical case management versus case management with problem-solving therapy in low-income, disabled elders with major depression: A randomized clinical trial. American Journal of Geriatric Psychiatry. 2016;24(1):50-9.

Alhusen, J. L., Hayat, M. J., & Borg, L. (2021, Feb). A pilot study of a group-based perinatal depression intervention on reducing depressive symptoms and improving maternal-fetal attachment and maternal sensitivity. Arch Womens Ment Health, 24(1), 145-154. https://doi.org/10.1007/s00737-020-01032-0

Allart-Van Dam E, Hosman CMH, Hoogduin CAL, Schaap CPDR. The coping with depression course: Short-term outcomes and mediating effects of a randomized controlled trial in the treatment of subclinical depression. Behavior Therapy. 2003;34(3):381-96.

Amani, B., Merza, D., Savoy, C., Streiner, D., Bieling, P., Ferro, M. A., & Van Lieshout, R. J. (2021, Nov 9). Peer-Delivered Cognitive-Behavioral Therapy for Postpartum Depression: A Randomized Controlled Trial. J Clin Psychiatry, 83(1).

Amano, M., Katayama, N., Umeda, S., Terasawa, Y., Tabuchi, H., Kikuchi, T., . . . Nakagawa, A. (2023). The effect of cognitive behavioral therapy on future thinking in patients with major depressive disorder: A randomized controlled trial. Frontiers in Psychiatry, 14. doi:10.3389/fpsyt.2023.997154

Ammerman RT, Putnam FW, Altaye M, Stevens J, Teeters AR, Van Ginkel JB. A clinical trial of in-home CBT for depressed mothers in home visitation. Behavior Therapy. 2013;44(3):359-72.

Andersson G, Bergström J, Holländare F, Carlbring P, Kaldo V, Ekselius L. Internet-based self-help for depression: Randomised controlled trial. British Journal of Psychiatry. 2005;187(5):456-61.

Araya, R., Menezes, P. R., Claro, H. G., Brandt, L. R., Daley, K. L., Quayle, J., Diez-Canseco, F., Peters, T. J., Vera Cruz, D., Toyama, M., Aschar, S., Hidalgo-Padilla, L., Martins, H., Cavero, V., Rocha, T., Scotton, G., de Almeida Lopes, I. F., Begale, M., Mohr, D. C., & Miranda, J. J. (2021, May 11). Effect of a Digital Intervention on Depressive Symptoms in Patients With Comorbid Hypertension or Diabetes in Brazil and Peru: Two Randomized Clinical Trials. Jama, 325(18), 1852-1862.

Arean PA, Perri MG, Nezu AM, Schein RL, Christopher F, Joseph TX. Comparative effectiveness of social problem-solving therapy and reminiscence therapy as treatments for depression in older adults. Journal of Consulting and Clinical Psychology. 1993;61(6): 1003-10.

Arjadi R, Nauta MH, Scholte WF, et al. Internet-based behavioural activation with lay counsellor support versus online minimal psychoeducation without support for treatment of depression: a randomised controlled trial in Indonesia. The lancet psychiatry 2018; 5(9): 707-16.

Au, A., Nan, H., Sum, R., Ng, F., Kwong, A., & Wong, S. (2022). Cognitive behavioural therapy for adherence and sub-clinical depression in type 2 diabetes: a randomised controlled trial (abridged secondary publication). Hong Kong Med J, 28 Suppl 3(3), 21-23.

Ayen I, Hautzinger M. Cognitive behavior therapy for depression in menopausal women. A controlled, randomized treatment study. Zeitschrift fur Klinische Psychologie und Psychotherapie. 2004;33(4):290-9.

Babiy, Z., Layton, H., Savoy, C. D., Xie, F., Brown, J. S. L., Bieling, P. J., . . . Van Lieshout, R. J. (2024). One-Day Peer-Delivered Cognitive Behavioral Therapy-Based Workshops for Postpartum Depression: A Randomized Controlled Trial. Psychotherapy and Psychosomatics, 93(2), 129-140. doi:https://doi.org/10.1159/000536040

Baker AL, Kavanagh DJ, Kay-Lambkin FJ, Hunt SA, Lewin TJ, Carr VJ, et al. Randomized controlled trial of cognitive-behavioural therapy for coexisting depression and alcohol problems: short-term outcome. Addiction (Abingdon, England). 2010;105(1):87-99.

Barber JP, Barrett MS, Gallop R, Rynn MA, Rickels K. Short-term dynamic psychotherapy versus pharmacotherapy for major depressive disorder: A randomized, placebo-controlled trial. Journal of Clinical Psychiatry. 2012;73(1):66-73.

Barnhofer T, Crane C, Hargus E, Amarasinghe M, Winder R, Williams JM. Mindfulness-based cognitive therapy as a treatment for chronic depression: A preliminary study. Behaviour Research and Therapy. 2009;47(5):366-73.

Barnhofer, T., Dunn, B. D., Strauss, C., Ruths, F. A., Barrett, B., Ryan, M., . . . Warren, F. C. (2025). Mindfulness-based cognitive therapy versus treatment as usual after non-remission with NHS Talking Therapies high-intensity psychological therapy for depression: a UK-based clinical effectiveness and cost-effectiveness randomised, controlled, superiority trial. Lancet Psychiatry, 12(6), 433-446.

Barrett JE, Williams Jr JW, Oxman TE, Frank E, Katon W, Sullivan M, et al. Treatment of dysthymia and minor depression in primary care: A ramdomized trial in patients aged 18 to 59 years. Journal of Family Practice. 2001;50(5):405-12.

Bashir, M., Saleem, T., & Arouj, K. (2025). Religiously Integrated Forgiveness Therapy: A Psychotherapeutic Tool for Depression and Pain Among Pakistani Patients with Chronic Widespread Pain. J Relig Health, 64(3), 2136-2151.

Basirat, Z., Kheirkhah, F., Faramarzi, M., Esmaelzadeh, S., Khafri, S., & Tajali, Z. (2022). Pharmacotherapy or Psychotherapy? Selective Treatment Depression in The Infertile Women with Recurrent

Pregnancy Loss: A Triple-Arm Randomized Controlled Trial. International Journal of Fertility and Sterility, 16(3), 211-219. doi: 10.22074/ijfs.2021.529258.1124

Baumeister, H., Paganini, S., Sander, L. B., Lin, J., Schlicker, S., Terhorst, Y., Moshagen, M., Bengel, J., Lehr, D., & Ebert, D. D. (2021). Effectiveness of a Guided Internet- and Mobile-Based Intervention for Patients with Chronic Back Pain and Depression (WARD-BP): A Multicenter, Pragmatic Randomized Controlled Trial. Psychother Psychosom, 90(4), 255-268.

Baumgartner, C., Schaub, M. P., Wenger, A., Malischnig, D., Augsburger, M., Lehr, D., . . . Haug, S. (2021). "Take Care of You" - Efficacy of integrated, minimal-guidance, internet-based self-help for reducing co-occurring alcohol misuse and depression symptoms in adults: Results of a three-arm randomized controlled trial. Drug Alcohol Depend, 225, 108806. doi:10.1016/j.drugalcdep.2021.108806

Beach SR, O'Leary KD. Treating depression in the context of marital discord: Outcome and predictors of response of marital therapy versus cognitive therapy. Behavior Therapy. 1992;23(4):507-28.

Bedard M, Felteau M, Marshall S, Cullen N, Gibbons C, Dubois S, et al. Mindfulness-based cognitive therapy reduces symptoms of depression in people with a traumatic brain injury: Results from a randomized controlled trial. Journal of Head Trauma Rehabilitation. 2014;29(4):E13-E22.

Beeber LS, Holditch-Davis D, Perreira K, Schwartz TA, Lewis V, Blanchard H, et al. Short-term in-home intervention reduces depressive symptoms in Early Head Start Latina mothers of infants and toddlers. Research in Nursing and Health. 2010;33(1): 60-76.

Behnammoghadam, M., Alamdari, A. K., Behnammoghadam, A., & Darban, F. (2015). Effect of Eye Movement Desensitization and Reprocessing (EMDR) on Depression in Patients With Myocardial Infarction (MI). Global journal of health science, 7(6), 258-262. doi:10.5539/gjhs.v7n6p258

Bendig, E., Bauereiß, N., Buntrock, C., Habibović, M., Ebert, D. D., & Baumeister, H. (2021). Lessons learned from an attempted randomized-controlled feasibility trial on "WIDeCAD" - An internet-based depression treatment for people living with coronary artery disease (CAD) [Article]. Internet Interventions, 24. https://doi.org/10.1016/j.invent.2021.100375

Bengtson AM, Filipowicz TR, Mphonda S, Udedi M, Kulisewa K, Meltzer-Brody S, et al. An Intervention to Improve Mental Health and HIV Care Engagement Among Perinatal Women in Malawi: A Pilot Randomized Controlled Trial. AIDS and behavior. 2023.

Berger T, Hämmerli K, Gubser N, Andersson G, Caspar F. Internet-based treatment of depression: A randomized controlled trial comparing guided with unguided self-help. Cognitive Behaviour Therapy. 2011;40(4):251-66.

Berman, M. I., Park, J., Kragenbrink, M. E., & Hegel, M. T. (2022). Accept Yourself! A Pilot Randomized Controlled Trial of a Self-Acceptance-Based Treatment for Large-Bodied Women With Depression. Behav Ther, 53(5), 913-926.

Beutel ME, Weissflog G, Leuteritz K, Wiltink J, Haselbacher A, Ruckes C, et al. Efficacy of short-term psychodynamic psychotherapy (STPP) with depressed breast cancer patients: Results of a randomized controlled multicenter trial. Annals of Oncology. 2014;25(2):378-84.

Bilich, L. L., Deane, F. P., Phipps, A. B., Barisic, M., & Gould, G. (2008). Effectiveness of bibliotherapy self-help for depression with varying levels of telephone helpline support. Clin Psychol Psychother, 15(2), 61-74. doi:10.1002/cpp.562

Bitew, T., Keynejad, R., Sorsdahl, K., Myers, B., Honikman, S., Medhin, G., . . . Hanlon, C. (2025). Brief problem-solving therapy (PST) for women experiencing antenatal depressive symptoms: a randomised, controlled feasibility trial in (an) antenatal care setting in rural Ethiopia. Pilot and Feasibility Studies, 11(1).

Blanco, V., Otero, P., & Vázquez, F. L. (2023). A pilot study for a smartphone app for the prevention of depression in non-professional caregivers. Aging Ment Health, 27(1), 166-175. doi: 10.1080/13607863.2022.2056878

Boele FW, Klein M, Verdonck-de Leeuw IM, et al. Internet-based guided self-help for glioma patients with depressive symptoms: a randomized controlled trial. Journal of neuro-oncology 2018; 137(1): 191-203.

Boele, F. W., et al. (2021). "The effects of SmartCare© on neuro-oncology family caregivers' distress: a randomized controlled trial." Supportive Care in Cancer.

Boeschoten, R. E., Dekker, J., Uitdehaag, B. M. J., Beekman, A. T. F., Hoogendoorn, A. W., Collette, E. H., . . . Van Oppen, P. (2017). Internet-based treatment for depression in multiple sclerosis: A randomized controlled trial. Multiple Sclerosis, 23(8), 1112-1122.

Bohlmeijer ET, Fledderus M, Rokx TA, Pieterse ME. Efficacy of an early intervention based on acceptance and commitment therapy for adults with depressive symptomatology: Evaluation in a randomized controlled trial. Behaviour research and therapy. 2011;49(1):62-7.

Bower, J. E., Partridge, A. H., Wolff, A. C., Thorner, E. D., Irwin, M. R., Joffe, H., Petersen, L., Crespi, C. M., & Ganz, P. A. (2021, Nov 1). Targeting Depressive Symptoms in Younger Breast Cancer Survivors: The Pathways to Wellness Randomized Controlled Trial of Mindfulness Meditation and Survivorship Education. J Clin Oncol, 39(31), 3473-3484.

Bowman D, Scogin F, Lyrene B. The efficacy of self-examination therapy and cognitive bibliotherapy in the treatment of mild to moderate depression. Psychotherapy Research. 1995;5(2): 131-40.

Braun, L., Titzler, I., Terhorst, Y., Freund, J., Thielecke, J., Ebert, D. D., & Baumeister, H. (2021, Jan 1). Effectiveness of guided internet-based interventions in the indicated prevention of depression in green professions (PROD-A): Results of a pragmatic randomized controlled trial. J Affect Disord, 278, 658-671. https://doi.org/10.1016/j.jad.2020.09.066

Brown RA, Lewinsohn PM. A psychoeducational approach to the treatment of depression: Comparison of group, individual, and minimal contact procedures. Journal of Consulting and Clinical Psychology. 1984;52(5):774.

Buhrman M, Syk M, Burvall O, Hartig T, Gordh T, Andersson G. Individualized Guided Internet-delivered Cognitive Behaviour Therapy for Chronic Pain Patients with Comorbid Depression and Anxiety: A Randomized Controlled Trial. Clinical Journal of Pain 2014.

Buntrock C, Ebert D, Lehr D, Riper H, Smit F, Cuijpers P, et al. Effectiveness of a web-based cognitive behavioural intervention for subthreshold depression: Pragmatic randomised controlled trial. Psychotherapy and Psychosomatics. 2015;84(6):348-58.

Burns A, Banerjee S, Morris J, Woodward Y, Baldwin R, Proctor R, et al. Treatment and prevention of depression after surgery for hip fracture in older people: Randomized, controlled trials. Journal of the American Geriatrics Society. 2007;55(1):75-80.

Burns A, O'Mahen H, Baxter H, Bennert K, Wiles N, Ramchandani P, et al. A pilot randomised controlled trial of cognitive behavioural therapy for antenatal depression. BMC psychiatry. 2013;13:33.

Carlbring P, Hagglund M, Luthstrom A, Dahlin M, Kadowaki A, Vernmark K, et al. Internet-based behavioral activation and acceptance-based treatment for depression: A randomized controlled trial. Journal of Affective Disorders. 2013;148(2-3): 331-7.

Carpenter KM, Smith JL, Aharonovich E, Nunes EV. Developing therapies for depression in drug dependence: Results of a stage 1 therapy study. American Journal of Drug and Alcohol Abuse. 2008;34(5):642-52.

Carr, A., Finnegan, L., Griffin, E., Cotter, P., & Hyland, A. (2017). A Randomized Controlled Trial of the Say Yes to Life (SYTL) Positive Psychology Group Psychotherapy Program for Depression: An Interim Report. Journal of Contemporary Psychotherapy, 47(3), 153-161.

Carta MG, Petretto D, Adamo S, Bhat KM, Lecca ME, Mura G, et al. Counseling in primary care improves depression and quality of life. Clinical Practice and Epidemiology in Mental Health. 2012;8.

Casanas R, Catalan R, del Val JL, Real J, Valero S, Casas M. Effectiveness of a psycho-educational group program for major depression in primary care: A randomized controlled trial. BMC psychiatry. 2012;12:230.

Chan AS, Wong QY, Sze SL, Kwong PP, Han YM, Cheung MC. A Chinese Chan-based mind-body intervention for patients with depression. Journal of Affective Disorders. 2012;142(1-3):283-9.

Chan MF, Ng SE, Tien A, Man Ho RC, Thayala J. A randomised controlled study to explore the effect of life story review on depression in older Chinese in Singapore. Health and Social Care in the Community. 2013;21(5):545-53.

Chen CH, Tseng YF, Chou FH, Wang SY. Effects of support group intervention in postnatally distressed women. A controlled study in Taiwan. Journal of psychosomatic research. 2000;49(6): 395-9.

Chesney MA, Chambers DB, Taylor JM, Johnson LM, Folkman S. Coping effectiveness training for men living with HIV: Results from a randomized clinical trial testing a group-based intervention. Psychosomatic Medicine. 2003;65(6):1038-46.

Chiang KJ, Chen TH, Hsieh HT, Tsai JC, Ou KL, Chou KR. One-year follow-up of the effectiveness of cognitive behavioral group therapy for patients' depression: A randomized, single-blinded, controlled study. Scientific World Journal. 2015;2015:Article ID 373149.

Cho HJ, Kwon JH, Lee JJ. Antenatal cognitive-behavioral therapy for prevention of postpartum depression: A pilot study. Yonsei medical journal. 2008;49(4):553-62.

Choi I, Zou J, Titov N, Dear BF, Li S, Johnston L, et al. Culturally attuned Internet treatment for depression amongst Chinese Australians: A randomised controlled trial. Journal of Affective Disorders. 2012;136(3):459-68.

Choi, N. G., Hegel, M. T., Marti, C. N., Marinucci, M. L., Sirrianni, L., & Bruce, M. L. (2014). Telehealth problem-solving therapy for depressed low-income homebound older adults. American Journal of Geriatric Psychiatry, 22(3), 263-271.

Choi NG, Marti CN, Wilson NL, et al. Effect of Telehealth Treatment by Lay Counselors vs by Clinicians on Depressive Symptoms Among Older Adults Who Are Homebound: A Randomized Clinical Trial. JAMA Netw Open. 2020;3(8):e2015648.

Chowdhary N, Anand A, Dimidjian S, Shinde S, Weobong B, Balaji M, et al. The Healthy Activity Program lay counsellor delivered treatment for severe depression in India: Systematic development and randomised evaluation. British Journal of Psychiatry. 2016;208(4):381-8.

Choy JC, Lou VW. Effectiveness of the modified instrumental reminiscence intervention on psychological well-being among community-dwelling chinese older adults: A randomized controlled trial. American Journal of Geriatric Psychiatry. 2016;24(1):60-9.

Christ, C., van Schaik, D. J. F., Kikkert, M. J., de Waal, M. M., Dozeman, E., Hulstijn, H. L., . . . Dekker, J. J. M. (2024). Internet-based emotion regulation training aimed at reducing violent revictimization and depressive symptoms in victimized depressed patients: Results of a randomized controlled trial. Journal of Affective Disorders, 355, 95-103. doi:https://doi.org/10.1016/j.jad.2024.03.028

Cladder-Micus, M. B., Speckens, A. E. M., Vrijsen, J. N., A.R, T. D., Becker, E. S., & Spijker, J. (2018). Mindfulness-based cognitive therapy for patients with chronic, treatment-resistant depression: A pragmatic randomized controlled trial. Depress Anxiety, 35(10), 914-924.

Clark R, Tluczek A, Brown R. A mother–infant therapy group model for postpartum depression. Infant Mental Health Journal. 2008;29(5):514-36.

Cohen S, O'Leary KD, Foran H. A randomized clinical trial of a brief, problem-focused couple therapy for depression. Behavior Therapy. 2010;41(4):433-46.

Cooper PJ, Murray L, Wilson A, Romaniuk H. Controlled trial of the short- and long-term effect of psychological treatment of post-partum depression. I. Impact on maternal mood. The British Journal of Psychiatry. 2003;182(5):412-9.

Cooper, Z. W., Mowbray, O., Ali, M. K., & Johnson, L. C. M. (2024). Addressing depression and comorbid health conditions through solution-focused brief therapy in an integrated care setting: a randomized clinical trial. BMC Prim Care, 25(1), 313.

Cramer H, Salisbury C, Conrad J, Eldred J, Araya R. Group cognitive behavioural therapy for women with depression: Pilot and feasibility study for a randomised controlled trial using mixed methods. BMC psychiatry. 2011;11:82.

Cuijpers, P., Heim, E., Abi Ramia, J., Burchert, S., Carswell, K., Cornelisz, I., . . . El Chammay, R. (2022). Effects of a WHO-guided digital health intervention for depression in Syrian refugees in Lebanon: A randomized controlled trial. PLoS Med, 19(6), e1004025. doi:10.1371/journal.pmed.1004025

Cuijpers, P., Heim, E., Ramia, J. A., Burchert, S., Carswell, K., Cornelisz, I., . . . El Chammay, R. (2022). Guided digital health intervention for depression in Lebanon: randomised trial. Evidence Based Mental Health, 25(e1), e34.

Cukor, D., Ver Halen, N., Asher, D. R., Coplan, J. D., Weedon, J., Wyka, K. E., . . . Kimmel, P. L. (2014). Psychosocial intervention improves depression, quality of life, and fluid adherence in hemodialysis. Journal of the American Society of Nephrology, 25(1), 196-206. doi:https://doi.org/10.1681/asn.2012111134

Dai, J., Huang, Y., Liu, J., Sun, Y., Fan, X., Tu, Y., . . . Liu, Y. (2025). Effect of the thinking healthy programme-based internet intervention model for maternal perinatal depression: A pilot study. Journal of Affective Disorders, 374, 294-302.

Davoudi M, Taheri AA, Foroughi AA, Ahmadi SM, Heshmati K. Effectiveness of acceptance and commitment therapy (ACT) on depression and sleep quality in painful diabetic neuropathy: a randomized clinical trial. Journal of Diabetes and Metabolic Disorders. 2020.

De Groot, M., Shubrook, J. H., Hornsby, W. G., Pillay, Y., Mather, K. J., Fitzpatrick, K., . . . Saha, C. (2019). Program ACTIVE II: Outcomes from a randomized, multistate community-based depression treatment for rural and urban adults with type 2 diabetes. Diabetes care, 42(7), 1185-1193. doi:10.2337/dc18-2400

De Jong M, Peeters F, Gard T, et al. A randomized controlled pilot study on mindfulness-based cognitive therapy for unipolar depression in patients with chronic pain. Journal of clinical psychiatry 2018; 79(1): 26-34.

Dekker RL, Moser DK, Peden AR, Lennie TA. Cognitive therapy improves three-month outcomes in hospitalized patients with heart failure. Journal of cardiac failure. 2012;18(1):10-20.

Demir, S., & Ercan, F. (2022). The effectiveness of cognitive behavioral therapy-based group counseling on depressive symptomatology, anxiety levels, automatic thoughts, and coping ways Turkish nursing students: A randomized controlled trial. Perspectives in psychiatric care. doi:10.1111/ppc.13073

Dennis CL, Grigoriadis S, Zupancic J, Kiss A, Ravitz P. Telephone-based nurse-delivered interpersonal psychotherapy for postpartum depression: nationwide randomised controlled trial. Br J Psychiatry. 2020;216(4):189-196.

DeRubeis RJ, Hollon SD, Amsterdam JD, Shelton RC, Young PR, Salomon RM, et al. Cognitive therapy vs medications in the treatment of moderate to severe depression. Archives of General Psychiatry. 2005;62(4):409-16.

Desautels, C., Savard, J., Ivers, H., Savard, M. H., & Caplette-Gingras, A. (2018). Treatment of depressive symptoms in patients with breast cancer: a randomized controlled trial comparing cognitive therapy and bright light therapy [Journal Article; Randomized Controlled Trial]. Health Psychology, 37(1), 1-13. https://doi.org/10.1037/hea0000539

Dimidjian S, Hollon SD, Dobson KS, Schmaling KB, Kohlenberg RJ, Addis ME, et al. Randomized trial of behavioral activation, cognitive therapy, and antidepressant medication in the acute treatment of adults with major depression. Journal of Consulting and Clinical Psychology. 2006;74(4):658-70.

Dimidjian, S., Goodman, S., Sherwood, N., Simon, G., Ludman, E., Gallop, R., . . . Beck, A. (2017). A Pragmatic Randomized Clinical Trial of Behavioral Activation for Depressed Pregnant Women. Journal of Consulting and Clinical Psychology, 85(1), 26-36

Dindo, L. N., Recober, A., Calarge, C. A., Zimmerman, B. M., Weinrib, A., Marchman, J. N., & Turvey, C. (2019). One-Day Acceptance and Commitment Therapy Compared to Support for Depressed Migraine Patients: a Randomized Clinical Trial. Neurotherapeutics. doi:10.1007/s13311-019-00818-0

Dobkin RD, Menza M, Allen LA, Gara MA, Mark MH, Tiu J, et al. Cognitive-behavioral therapy for depression in Parkinson's disease: A randomized, controlled trial. American Journal of Psychiatry. 2011;168(10):1066-74.

Dobkin RD, Mann SL, Gara MA, Interian A, Rodriguez KM, Menza M. Telephone-based cognitive behavioral therapy for depression in Parkinson disease: A randomized controlled trial. Neurology. 2020;94(16):e1764-e1773.

Dobkin, R. D., Mann, S. L., Weintraub, D., Rodriguez, K. M., Miller, R. B., St. Hill, L., King, A., Gara, M. A., & Interian, A. (2021). Innovating Parkinson's Care: A Randomized Controlled Trial of Telemedicine Depression Treatment [Article]. Movement disorders, 36(11), 2549-2558.

Doering LV, Chen B, Cross Bodan R, Magsarili MC, Nyamathi A, Irwin MR. Early cognitive behavioral therapy for depression after cardiac surgery. Journal of Cardiovascular Nursing. 2013;28(4): 370-9.

Dong, X., Sun, G., Zhan, J., Liu, F., Ma, S., Li, P., . . . Liu, Y. (2019). Telephone-based reminiscence therapy for colorectal cancer patients undergoing postoperative chemotherapy complicated with depression: a three-arm randomised controlled trial. Supportive care in cancer : official journal of the Multinational Association of Supportive Care in Cancer, 27(8), 2761-2769.

Dowrick, C., Dunn, G., Ayuso-Mateos, J. L., Dalgard, O. S., Page, H., Lehtinen, V., ... Wilkinson, G. (2000). Problem solving treatment and group psychoeducation for depression: multicentre randomised controlled trial Outcomes of Depression International Network [ODIN] Group. Bmj, 321(7274), 1450-1454.

Duan, C. C., Zhang, C., Xu, H. L., Tao, J., Yu, J. L., Zhang, D., . . . Wu, Y. T. (2025). Internet-Based Cognitive Behavioral Therapy for Preventing Postpartum Depressive Symptoms Among Pregnant Individuals With Depression: Multicenter Randomized Controlled Trial in China. Journal of Medical Internet Research, 27. doi:https://doi.org/10.2196/67386

Duarte PS, Miyazaki MC, Blay SL, Sesso R. Cognitive-behavioral group therapy is an effective treatment for major depression in hemodialysis patients. Kidney international. 2009;76(4):414-21.

Dunn NJ, Rehm LP, Schillaci J, Souchek J, Mehta P, Ashton CM, et al. A randomized trial of self-management and psychoeducational group therapies for comorbid chronic posttraumatic stress disorder and depressive disorder. Journal of traumatic stress. 2007;20(3):221-37.

Dwight-Johnson M, Aisenberg E, Golinelli D, Hong S, O'Brien M, Ludman E. Telephone-based cognitive-behavioral therapy for Latino patients living in rural areas: A randomized pilot study. Psychiatric Services. 2011;62(8):936-42.

Ebert, D. D., Lehr, D., Boß, L., Riper, H., Cuijpers, P., Andersson, G., . . . Berking, M. (2014). Efficacy of an internet-based problem-solving training for teachers: results of a randomized controlled trial. Scandinavian journal of work, environment & health, 582-596.

Ebert DD, Buntrock C, Lehr D, et al. Effectiveness of Web- and Mobile-Based Treatment of Subthreshold Depression With Adherence-Focused Guidance: a Single-Blind Randomized Controlled Trial. Behavior therapy 2018; 49(1): 71-83.

Ede MO, Igbo JN, Eseadi C, et al. Effect of group cognitive behavioural therapy on depressive symptoms in a sample of college adolescents in Nigeria. Journal of Rational-Emotive & Cognitive-Behavior Therapy. 2020;38(3):306-318.

Eimontas, J., Medeišienė, J., Baranova, K., Biliūnaitė, I., Pakalniškienė, V., Zrumaitė, S., . . . Andersson, G. (2025). Tailored internet-delivered cognitive behavioral therapy with therapist support for depressed older adults: Results from a randomized controlled trial with a one-year follow-up. J Affect Disord, 385, 119406.

Ekers D, Richards D, McMillan D, Bland JM, Gilbody S. Behavioural activation delivered by the non-specialist: Phase II randomised controlled trial. The British Journal of Psychiatry. 2011;198(1): 66-72.

Ekkers W, Korrelboom K, Huijbrechts I, Smits N, Cuijpers P, Gaag M. Competitive memory training for treating depression and rumination in depressed older adults: A randomized controlled trial. Behaviour Research and Therapy. 2011;49(10):588-96.

El-Haj-Mohamad R, Böttche M, Vöhringer M, Specht F, Stammel N, Nesterko Y, et al. An internet-based cognitive behavioural intervention for adults with depression in Arabic-speaking countries: A randomized controlled trial. Stress Health. 2024;40(5):e3432.

El-Haj-Mohamad R, Stein J, Stammel N, Nesterko Y, Wagner B, Böttche M, et al. Efficacy of internet-based cognitive behavioral and interpersonal treatment for depression in Arabic speaking countries: A randomized controlled trial. J Affect Disord. 2025;368:573-83.

El-Monshed, A. H., Khonji, L. M., Altheeb, M., Saad, M. T. E., Elsheikh, M. A., Loutfy, A., . . . Zoromba, M. A. (2024). Does a program-based cognitive behavioral therapy affect insomnia and depression in menopausal women? A randomized controlled trial. Worldviews Evid Based Nurs, 21(2), 202-215. doi:https://doi.org/10.1111/wvn.12707

Elkin I, Shea MT, Watkins JT, Imber SD, Sotsky SM, Collins JF, et al. National institute of mental health treatment of depression collaborative research program. General effectiveness of treatments. Archives of General Psychiatry. 1989;46(11):971-82; discussion 83.

Embling S. The effectiveness of cognitive behavioural therapy in depression. Nursing Standard. 2002;17(14-15):33-41.

Eseadi C, Obidoa MA, Ogbuabor SE, Ikechukwu-Ilomuanya AB. Effects of Group-Focused Cognitive-Behavioral Coaching Program on Depressive Symptoms in a Sample of Inmates in a Nigerian Prison. International journal of offender therapy and comparative criminology 2018; 62(6): 1589-602.

Eseadi, C., Ilechukwu, L. C., Victor-Aigbodion, V., Sewagegn, A. A., & Amedu, A. N. (2022). Intervention for depression among undergraduate religious education students: A randomized controlled trial. Medicine (Baltimore), 101(41), e31034.

Euteneuer, F., Dannehl, K., Del Rey, A., Engler, H., Schedlowski, M., & Rief, W. (2017). Immunological effects of behavioral activation with exercise in major depression: An exploratory randomized controlled trial. Translational Psychiatry, 7(5).

Euteneuer, F., Neuert, M., Salzmann, S., Fischer, S., Ehlert, U., & Rief, W. (2022). Does psychological treatment of major depression reduce cardiac risk biomarkers? An exploratory randomized controlled trial. Psychological medicine, 1-15.

Ewais, T., et al. (2021). "Mindfulness based cognitive therapy for youth with inflammatory bowel disease and depression - Findings from a pilot randomised controlled trial." J Psychosom Res 149: 110594.

Fann JR, Bombardier CH, Vannoy S, Dyer J, Ludman E, Dikmen S, et al. Telephone and in-person cognitive behavioral therapy for major depression after traumatic brain injury: A randomized controlled trial. Journal of neurotrauma. 2015;32(1):45-57.

Faramarzi M, Alipor A, Esmaelzadeh S, Kheirkhah F, Poladi K, Pash H. Treatment of depression and anxiety in infertile women: Cognitive behavioral therapy versus fluoxetine. Journal of Affective Disorders. 2008;108(1-2):159-64.

Fereydouni, S., & Forstmeier, S. (2022). An Islamic Form of Logotherapy in the Treatment of Depression, Anxiety and Stress Symptoms in University Students in Iran. J Relig Health, 61(1), 139-157. doi:10.1007/s10943-021-01495-0

Fissler, M., Winnebeck, E., Schroeter, T. A., Gummbersbach, M., Huntenburg, J. M., Gärtner, M., & Barnhofer, T. (2017). Brief training in mindfulness may normalize a blunted error-related negativity in chronically depressed patients. Cognitive, Affective & Behavioral Neuroscience, 17(6), 1164-1175. doi:10.3758/s13415-017-0540-x

Fledderus M, Bohlmeijer ET, Pieterse ME, Schreurs KM. Acceptance and commitment therapy as guided self-help for psychological distress and positive mental health: A randomized controlled trial. Psychological Medicine. 2012;42(3):485-95.

Floyd M, Scogin F, McKendree-Smith NL, Floyd DL, Rokke PD. Cognitive therapy for depression: A comparison of individual psychotherapy and bibliotherapy for depressed older adults. Behavior modification. 2004;28(2):297-318.

Flygare AL, Engström I, Hasselgren M, et al. Internet-based CBT for patients with depressive disorders in primary and psychiatric care: Is it effective and does comorbidity affect outcome? Internet Interventions. 2020;19.

Folke F, Parling T, Melin L. Acceptance and commitment therapy for depression: A preliminary randomized clinical trial for unemployed on long-term sick leave. Cognitive and Behavioral Practice. 2012;19(4):583-94.

Fonagy P, Rost F, Carlyle JA, McPherson S, Thomas R, Pasco Fearon RM, et al. Pragmatic randomized controlled trial of long-term psychoanalytic psychotherapy for treatment-resistant depression: The Tavistock Adult Depression Study (TADS). World Psychiatry. 2015;14(3):312-21.

Fonagy P, Lemma A, Target M, O'Keeffe S, Constantinou MP, Ventura Wurman T, et al. Dynamic interpersonal therapy for moderate to severe depression: a pilot randomized controlled and feasibility trial. Psychological medicine. 2019:1-10.

Forand NR, Barnett JG, Strunk DR, Hindiyeh MU, Feinberg JE, Keefe JR. Efficacy of Guided iCBT for Depression and Mediation of Change by Cognitive Skill Acquisition. Behavior Therapy 2018; 49(2): 295-307.

Forman-Hoffman, V. L., Sihvonen, S., Wielgosz, J., Kuhn, E., Nelson, B. W., Peiper, N. C., & Gould, C. E. (2024). Therapist-supported digital mental health intervention for depressive symptoms: A randomized clinical trial. Journal of Affective Disorders, 349, 494-501. doi:https://doi.org/10.1016/j.jad.2024.01.057

Forsell E, Bendix M, Holländare F, et al. Internet delivered cognitive behavior therapy for antenatal depression: A randomised controlled trial. Journal of Affective Disorders 2017; 221: 56-64.

Franco, P., Olhaberry, M., Kelders, S., Muzard, A., & Cuijpers, P. (2024). Guided web app intervention for reducing symptoms of depression in postpartum women: Results of a feasibility randomized controlled trial. Internet Interventions, 36, 100744. doi:https://doi.org/10.1016/j.invent.2024.100744

Frangou, E., Bertelli, G., Love, S., Mackean, M. J., Glasspool, R. M., Fotopoulou, C., Cook, A., Nicum, S., Lord, R., Ferguson, M., Roux, R. L., Martinez, M., Butcher, C., Hulbert-Williams, N., Howells, L.,

& Blagden, S. P. (2021). OVPSYCH2: A randomized controlled trial of psychological support versus standard of care following chemotherapy for ovarian cancer. Gynecol Oncol, 162(2), 431-439.

Freedland KE, Skala JA, Carney RM, Rubin EH, Lustman PJ, D·vila-Rom·n VG, et al. Treatment of depression after coronary artery bypass surgery: A randomized controlled trial. Archives of General Psychiatry. 2009;66(4):387-96.

Freedland KE, Carney RM, Rich MW, Steinmeyer BC, Rubin EH. Cognitive behavior therapy for depression and self-care in heart failure patients: A randomized clinical trial. JAMA Internal Medicine. 2015;175(11):1773-82.

Fry P. Structured and unstructured reminiscence training and depression among the elderly. Clinical Gerontologist. 1983;1(3): 15-37.

Fuhr, D. C., Weobong, B., Lazarus, A., Vanobberghen, F., Weiss, H. A., Singla, D. R., . . . Patel, V. (2019). Delivering the Thinking Healthy Programme for perinatal depression through peers: An individually randomised controlled trial in India. The Lancet Psychiatry, 6(2), 115-127. doi:10.1016/S2215-0366(18)30466-8

Funderburk, J. S., Pigeon, W. R., Shepardson, R. L., Wade, M., Acker, J., Fivecoat, H., Wray, L. O., & Maisto, S. A. (2021). Treating depressive symptoms among veterans in primary care: A multi-site RCT of brief behavioral activation. J Affect Disord, 283, 11-19.

Furukawa TA, Horikoshi M, Kawakami N, Kadota M, Sasaki M, Sekiya Y, et al. Telephone cognitive-behavioral therapy for subthreshold depression and presenteeism in workplace: A randomized controlled trial. PLoS One. 2012;7(4):e35330.

García-Peña C, Vázquez-Estupiñan F, Avalos-Pérez F, Jiménez LVR, Sánchez-Garcia S, Juárez-Cedillo T. Clinical effectiveness of group cognitive-behavioural therapy for depressed older people in primary care: A randomised controlled trial. Salud Mental. 2015;38(1):33-9.

Garcia, A., Yáñez, A. M., Bennasar-Veny, M., Navarro, C., Salva, J., Ibarra, O., . . . Garcia-Toro, M. (2023). Efficacy of an adjuvant non-face-to-face multimodal lifestyle modification program for patients with treatment-resistant major depression: A randomized controlled trial. Psychiatry Res, 319, 114975. doi:10.1016/j.psychres.2022.114975

Gawrysiak M, Nicholas C, Hopko DR. Behavioral activation for moderately depressed university students: Randomized controlled trial. Journal of Counseling Psychology. 2009;56(3): 468-75.

Gellis ZD, McGinty J, Tierney L, Jordan C, Burton J, Misener E. Randomized controlled trial of problem-solving therapy for minor depression in home care. Research on Social Work Practice. 2008;18(6):596-606.

Gellis ZD, Bruce ML. Problem solving therapy for subthreshold depression in home healthcare patients with cardiovascular disease. The American Journal of Geriatric Psychiatry. 2010;18(6):464-74.

Geraedts AS, Kleiboer AM, Wiezer NM, van Mechelen W, Cuijpers P. Short-term effects of a web-based guided self-help intervention for employees with depressive symptoms: Randomized controlled trial. Journal of Medical Internet Research. 2014;16(5):e121.

Ghorbani, V., Zanjani, Z., Omidi, A., & Sarvizadeh, M. (2021). Efficacy of acceptance and commitment therapy (ACT) on depression, pain acceptance, and psychological flexibility in married women with breast cancer: a pre- and post-test clinical trial. Trends Psychiatry Psychother, 43(2), 126-133. https://doi.org/10.47626/2237-6089-2020-0022

Gibbons MB, Thompson SM, Scott K, Schauble LA, Mooney T, Thompson D, et al. Supportive-expressive dynamic psychotherapy in the community mental health system: A pilot effectiveness trial for the treatment of depression. Psychotherapy (Chicago, Ill). 2012;49(3):303-16.

Gilbody, S., Littlewood, E., McMillan, D., Atha, L., Bailey, D., Baird, K., . . . Ekers, D. (2024). Behavioural activation to mitigate the psychological impacts of COVID-19 restrictions on older people in England and Wales (BASIL+): a pragmatic randomised controlled trial. The Lancet Healthy Longevity, 5(2), e97-e107. doi:https://doi.org/10.1016/s2666-7568(23)00238-6

Gitlin LN, Harris LF, McCoy MC, Chernett NL, Pizzi LT, Jutkowitz E, et al. A home-based intervention to reduce depressive symptoms and improve quality of life in older African Americans: A randomized trial. Annals of Internal Medicine. 2013;159(4): 243-52.

Gold SM, Friede T, Meyer B, Moss-Morris R, Hudson J, Asseyer S, et al. Internet-delivered cognitive behavioural therapy programme to reduce depressive symptoms in patients with multiple sclerosis: a multicentre, randomised, controlled, phase 3 trial. The Lancet Digital Health. 2023;5(10):e668-e78.

Goodman JH, Prager J, Goldstein R, Freeman M. Perinatal Dyadic Psychotherapy for postpartum depression: A randomized controlled pilot trial. Archives of Women's Mental Health. 2015;18(3):493-506.

Greenberg J, Datta T, Shapero BG, Sevinc G, Mischoulon D, Lazar SW. Compassionate hearts protect against wandering minds: Self-compassion moderates the effect of mind-wandering on depression. Spirituality in Clinical Practice 2018; 5(3): 155-69.

Grote NK, Swartz HA, Geibel SL, Zuckoff A, Houck PR, Frank E. A randomized controlled trial of culturally relevant, brief interpersonal psychotherapy for perinatal depression. Psychiatric Services. 2009;60(3):313-21.

Haakana, R., Rosenström, T., Parkkinen, L., Tuomisto, M. T., & Isometsä, E. (2024). Effectiveness of an add-on brief group behavioral activation treatment for depression in psychiatric care: a randomized clinical trial. Frontiers in Psychiatry, 15. doi:https://doi.org/10.3389/fpsyt.2024.1284363

Hagen, R., Hjemdal, O., Solem, S., Kennair, L. E. O., Nordahl, H. M., Fisher, P., & Wells, A. (2017). Metacognitive therapy for depression in adults: A waiting list randomized controlled trial with six months follow-up. Frontiers in Psychology, 8.

Hallford DJ, Mellor D. Autobiographical memory-based intervention for depressive symptoms in young adults: A randomized controlled trial of cognitive-reminiscence therapy. Psychotherapy and Psychosomatics. 2016;85(4):246-9.

Hallgren M, Kraepelien M, Öjehagen A, Lindefors N, Zeebari Z, Kaldo V, et al. Physical exercise and internet-based cognitive-behavioural therapy in the treatment of depression: Randomised controlled trial. British Journal of Psychiatry. 2015;207(3):227-34.

Hamamci Z. Integrating psychodrama and cognitive behavioral therapy to treat moderate depression. Arts in Psychotherapy. 2006;33(3):199-207.

Hamdan-Mansour AM, Puskar K, Bandak AG. Effectiveness of cognitive-behavioral therapy on depressive symptomatology, stress and coping strategies among Jordanian university students. Issues in mental health nursing. 2009;30(3):188-96.

Han, A., Wilroy, J. D., Yuen, H. K., Jenkins, J., & Hawkins, J. (2025). Videoconference-delivered acceptance and commitment therapy for depressed individuals with spinal cord injury sustained within the past 5 years: A pilot randomized controlled trial. Rehabilitation Psychology.

Haringsma R, Engels G, Cuijpers P, Spinhoven P. Effectiveness of the Coping With Depression (CWD) course for older adults provided by the community-based mental health care system in the Netherlands: A randomized controlled field trial. International Psychogeriatrics. 2006;18(02):307-25.

Harley R, Sprich S, Safren S, Jacobo M, Fava M. Adaptation of dialectical behavior therapy skills training group for treatment-resistant depression. Journal of Nervous and Mental Disease. 2008;196(2):136-43.

Harrer, M., Apolinário-Hagen, J., Fritsche, L., Salewski, C., Zarski, A. C., Lehr, D., Baumeister, H., Cuijpers, P., & Ebert, D. D. (2021). Effect of an internet- and app-based stress intervention compared to online psychoeducation in university students with depressive symptoms: Results of a randomized controlled trial [Article]. Internet Interventions, 24.

Hasani, M., Zenoozian, S., Ahmadi, R., Khakpoor, S., Saberi, S., Pirzeh, R., & Saed, O. (2025). Evaluating the efficacy of rumination-focused cognitive-behavioral therapy in alleviating depression, negative affect, and rumination among patients with recurrent major depressive disorder: a randomized, multicenter clinical trial. BMC Psychiatry, 25(1), 626.

Hashemi, Z., Eyni, S., & Ebadi, M. (2022). Effectiveness of Acceptance and Commitment Therapy in Depression and Anxiety in People with Substance Use Disorder. Iranian Journal of Psychiatry and Behavioral Sciences, 16(1). doi:10.5812/ijpbs.110135

Hassanvandi, S., Mohammadzadegan, R., Saadat, S. H., Hatef, B., & Shahyad, S. (2025). Effectiveness of Mindfulness-Based Stress Reduction on Salivary Cortisol and α-amylase Level, Anxiety and Depression in Students: A Randomized and Parallel-Group Clinical Trial. Appl Psychophysiol Biofeedback, 50(3), 417-428.

Hassiotis A, Serfaty M, Azam K, Strydom A, Blizard R, Romeo R, et al. Manualised individual cognitive behavioural therapy for mood disorders in people with mild to moderate intellectual disability: A feasibility randomised controlled trial. Journal of Affective Disorders. 2013;151(1):186-95.

Hautzinger M, Welz S. Kognitive Verhaltenstherapie bei Depressionen im Alter: Ergebnisse einer kontrollierten Vergleichsstudie unter ambulanten Bedingungen an Depressionen mittleren Schweregrads. = Cognitive behavioral therapy for depressed older outpatients: A controlled, randomized trial. Zeitschrift für Gerontologie und Geriatrie. 2004;37(6):427-35.

Hayati, M. S., Shams, J., Meibodi, S. S., Shafighi, A. H., Shattell, M., & Ghadirian, F. (2024). The effects of a telenursing scheduled intervention of brief behavioral activation therapy on depression and anxiety symptoms of patients with mixed depression and anxiety disorder: A randomized controlled trial. Arch Psychiatr Nurs, 52, 39-44. doi:https://doi.org/10.1016/j.apnu.2024.07.014

Hayman PM, Cope CS. Effects of assertion training on depression. Journal of clinical psychology. 1980;36(2):534-43.

He, H. L., Zhang, M., Gu, C. Z., Xue, R. R., Liu, H. X., Gao, C. F., & Duan, H. F. (2019). Effect of Cognitive Behavioral Therapy on Improving the Cognitive Function in Major and Minor Depression. The Journal of nervous and mental disease, 207(4), 232-238.

Heckman TG, Sikkema KJ, Hansen N, Kochman A, Heh V, Neufeld S, et al. A randomized clinical trial of a coping improvement group intervention for HIV-infected older adults. Journal of behavioral medicine. 2011;34(2):102-11.

Heckman, T. G., Heckman, B. D., Anderson, T., Lovejoy, T. I., Mohr, D., Sutton, M., . . . Gau, J.-T. (2013). Supportive-expressive and coping group teletherapies for HIV-infected older adults: a randomized clinical trial. AIDS Behav, 17(9), 3034-3044.

Heckman, T. G., Heckman, B. D., Anderson, T., Lovejoy, T. I., Markowitz, J. C., Shen, Y., & Sutton, M. (2017). Tele-interpersonal psychotherapy acutely reduces depressive symptoms in depressed HIV-infected rural persons: A randomized clinical trial. Behavioral Medicine, 43(4), 285-295.

Hegerl U, Hautzinger M, Mergl R, Kohnen R, Sch¸tze M, Scheunemann W, et al. Effects of pharmacotherapy and psychotherapy in depressed primary-care patients: A randomized, controlled trial including a patients' choice arm. International Journal of Neuropsychopharmacology. 2010;13(1):31-44.

Heim, E., Ramia, J. A., Hana, R. A., Burchert, S., Carswell, K., Cornelisz, I., . . . van't Hof, E. (2021). Step-by-step: Feasibility randomised controlled trial of a mobile-based intervention for depression among populations affected by adversity in Lebanon. Internet Interventions, 24, 100380. doi:https://doi.org/10.1016/j.invent.2021.100380

Heinrich, M., Zagorscak, P., Kampisiou, C., Bohn, J., Schulze, L., Schaeuffele, C., . . . Knaevelsrud, C. (2025). A randomized controlled trial of a therapist-guided online intervention for depressed adults and its utility as an adjunctive to antidepressants and psychotherapy. BMC Psychiatry, 25(1). doi:https://doi.org/10.1186/s12888-025-06564-2

Hemanny, C., Carvalho, C., Maia, N., Reis, D., Botelho, A. C., Bonavides, D., . . . De Oliveira, I. R. (2019). Efficacy of trial-based cognitive therapy, behavioral activation and treatment as usual in the treatment of major depressive disorder: Preliminary findings from a randomized clinical trial. CNS Spectrums. doi:10.1017/S1092852919001457

Hermanns N, Schmitt A, Gahr A, Herder C, Nowotny B, Roden M, et al. The effect of a diabetes-specific cognitive behavioral treatment program (DIAMOS) for patients with diabetes and subclinical depression: Results of a randomized controlled trial. Diabetes care. 2015;38(4):551-60.

Herrmann-Lingen C, Beutel ME, Bosbach A, Deter HC, Fritzsche K, Hellmich M, et al. A stepwise psychotherapy intervention for reducing risk in coronary artery disease (SPIRR-CAD): Results of an observer-blinded, multicenter, randomized trial in depressed patients with coronary artery disease. Psychosomatic Medicine. 2016;78(6):704-15.

Heshmati R, Wienicke FJ, Driessen E. The effects of intensive short-term dynamic psychotherapy on depressive symptoms, negative affect, and emotional repression in single treatment-resistant depression: A randomized controlled trial. Psychotherapy. 2023;60(4):497-511.

Hoifodt RS, Lillevoll KR, Griffiths KM, et al. The clinical effectiveness of web-based cognitive behavioral therapy with face-to-face therapist support for depressed primary care patients: randomized controlled trial. Journal of medical Internet research 2013; 15(8): e153.

Holden JM, Sagovsky R, Cox JL. Counselling in a general practice setting: Controlled study of heath visitor intervention in treatment of postnatal depression. British Medical Journal. 1989;298(6668):223-6.

Honey KL, Bennett P, Morgan M. A brief psycho-educational group intervention for postnatal depression. British Journal of Clinical Psychology. 2002;41(4):405-9.

Horrell L, Goldsmith KA, Tylee AT, Schmidt UH, Murphy CL, Bonin E-M, et al. One-day cognitive–behavioural therapy self-confidence workshops for people with depression: Randomised controlled trial. The British Journal of Psychiatry. 2014;204(3):222-33.

Hou Y, Hu P, Zhang Y, Lu Q, Wang D, Yin L, et al. Cognitive behavioral therapy in combination with systemic family therapy improves mild to moderate postpartum depression. Revista Brasileira de Psiquiatria. 2014;36(1):47-52.

Howard, R., Cort, E., Rawlinson, C., Wiegand, M., Downey, A., Lawrence, V., . . . Gould, R. (2024). Adapted problem adaptation therapy for depression in mild to moderate Alzheimer's disease dementia: A randomized controlled trial. Alzheimers Dement, 20(4), 2990-2999. doi:https://doi.org/10.1002/alz.13766

Hsiao, F.-H., Lai, Y.-M., Chen, Y.-T., Yang, T.-T., Liao, S.-C., Ho, R. T., Ng, S.-M., Chan, C. L., & Jow, G.-M. (2014). Efficacy of psychotherapy on diurnal cortisol patterns and suicidal ideation in adjustment disorder with depressed mood. General hospital psychiatry, 36(2), 214-219.

Huang, C.-Y., Lai, H.-L., Chen, C.-I., Lu, Y.-C., Li, S.-C., Wang, L.-W., & Su, Y. (2016). Effects of motivational enhancement therapy plus cognitive behaviour therapy on depressive symptoms and health-related quality of life in adults with type II diabetes mellitus: A randomised controlled trial. Quality of Life Research: An International Journal of Quality of Life Aspects of Treatment, Care & Rehabilitation, 25(5), 1275-1283.

Huh, K., Layton, H., Savoy, C. D., Ferro, M. A., Bieling, P. J., Hicks, A., & Van Lieshout, R. J. (2023). Online Public Health Nurse-Delivered Group Cognitive Behavioral Therapy for Postpartum Depression: A Randomized Controlled Trial During the COVID-19 Pandemic. J Clin Psychiatry, 84(5).

Hum, K. M., Chan, C. J., Gane, J., Conway, L., McAndrews, M. P., & Smith, M. L. (2019). Do distance-delivery group interventions improve depression in people with epilepsy? Epilepsy & Behavior, 98, 153-160. doi:10.1016/j.yebeh.2019.06.037

Hummel, J., Weisbrod, C., Boesch, L., Himpler, K., Hauer, K., Hautzinger, M., . . . Kopf, D. (2017). AIDE–Acute Illness and Depression in Elderly Patients. Cognitive Behavioral Group Psychotherapy in Geriatric Patients With Comorbid Depression: A Randomized, Controlled Trial. Journal of the american medical directors association, 18(4), 341-349.

Hunter SB, Watkins KE, Hepner KA, Paddock SM, Ewing BA, Osilla KC, et al. Treating depression and substance use: A randomized controlled trial. Journal of substance abuse treatment. 2012;43(2):137-51.

Husain, N., Zulqernain, F., Carter, L.-A., Chaudhry, I., Fatima, B., Kiran, T., . . . Rahman, A. (2017). Treatment of maternal depression in urban slums of Karachi, Pakistan: a randomized controlled trial (RCT) of an integrated maternal psychological and early child development intervention. Asian journal of psychiatry, 29, 63-70.

Husain, N., Kiran, T., Shah, S., Rahman, A., Raza Ur, R., Saeed, Q., Naeem, S., Bassett, P., Husain, M., Haq, S. U., Jaffery, F., Cohen, N., Naeem, F., & Chaudhry, N. (2021). Efficacy of learning through play plus intervention to reduce maternal depression in women with malnourished children: A randomized controlled trial from Pakistan(☆). J Affect Disord, 278, 78-84.

Husain N, Lunat F, Lovell K, Sharma D, Zaidi N, Bokhari A, et al. Exploratory RCT of a group psychological intervention for postnatal depression in British mothers of South Asian origin - ROSHNI-D. Acta Psychol (Amst). 2023;238:103974.

Husain N, Lunat F, Lovell K, Miah J, Chew-Graham CA, Bee P, et al. Efficacy of a culturally adapted, cognitive behavioural therapy-based intervention for postnatal depression in British south Asian women (ROSHNI-2): a multicentre, randomised controlled trial. Lancet. 2024;404(10461):1430-43.

Iacob, E., Kausler, R., Williams, M., Dorn, U., Simonsen, S., Smid, M., & Latendresse, G. (2025). A Randomized Controlled Trial of a Telehealth Group Intervention to Reduce Perinatal Depressive Symptoms: A Mixed Methods Analysis. J Midwifery Womens Health, 70(3), 431-441.

Jalali, F., Hasani, A., Hashemi, S. F., Kimiaei, S. A., & Babaei, A. (2019). Cognitive Group Therapy Based on Schema-Focused Approach for Reducing Depression in Prisoners Living With HIV. International journal of offender therapy and comparative criminology, 63(2), 276-288. doi:10.1177/0306624X18784185

Jamison C, Scogin F. The outcome of cognitive bibliotherapy with depressed adults. Journal of Consulting and Clinical Psychology. 1995;63(4):644-50.

Jarrett RB, Schaffer M, McIntire D, Witt-Browder A, Kraft D, Risser RC. Treatment of atypical depression with cognitive therapy or phenelzine: A double-blind, placebo-controlled trial. Archives of General Psychiatry. 1999;56(5):431-7.

Jelinek L, Hauschildt M, Wittekind CE, Schneider BC, Kriston L, Moritz S. Efficacy of metacognitive training for depression: A randomized controlled trial. Psychotherapy and Psychosomatics. 2016;85(4):231-4.

Jesse DE, Gaynes BN, Feldhousen EB, Newton ER, Bunch S, Hollon SD. Performance of a culturally tailored cognitive-behavioral intervention integrated in a public health setting to reduce risk of antepartum depression: A randomized controlled trial. Journal of Midwifery and Women's Health. 2015;60(5):578-92.

Jiang L, Wang ZZ, Qiu LR, Wan GB, Lin Y, Wei Z. Psychological intervention for postpartum depression. Journal of Huazhong University of Science and Technology: Medical sciences. 2014;34(3):437-42.

Johansson R, Ekbladh S, Hebert A, Lindström M, Möller S, Petitt E, et al. Psychodynamic guided self-help for adult depression through the internet: A randomised controlled trial. PloS One. 2012;7(5):e38021.

Johansson R, Sjöberg E, Sjögren M, Johnsson E, Carlbring P, Andersson T, et al. Tailored vs. standardized internet-based cognitive behavior therapy for depression and comorbid symptoms: A randomized controlled trial. PloS One. 2012;7(5):e36905.

Johansson, O., Bjärehed, J., Andersson, G., Carlbring, P., & Lundh, L. G. (2019). Effectiveness of guided internet-delivered cognitive behavior therapy for depression in routine psychiatry: A randomized controlled trial. Internet Interventions, 17.

Johansson P, Westas M, Andersson G, Alehagen U, Broström A, Jaarsma T, Mourad G, Lundgren J An Internet-Based Cognitive Behavioral Therapy Program Adapted to Patients With Cardiovascular Disease and Depression: Randomized Controlled Trial JMIR Ment Health 2019;6(10):e14648

Johnson JE, Zlotnick C. Pilot study of treatment for major depression among women prisoners with substance use disorder. Journal of Psychiatric Research. 2012;46(9):1174-83.

Johnson, J. E., Stout, R. L., Miller, T. R., Zlotnick, C., Cerbo, L. A., Andrade, J. T., . . . Wiltsey-Stirman, S. (2019). Randomized cost-effectiveness trial of group interpersonal psychotherapy (IPT) for prisoners with major depression. Journal of consulting and clinical psychology, 87(4), 392-406. doi:10.1037/ccp0000379

Joling KJ, Hout HP, van't Veer-Tazelaar PJ, Horst HE, Cuijpers P, Ven PM, et al. How effective is bibliotherapy for very old adults with subthreshold depression? A randomized controlled trial. American Journal of Geriatric Psychiatry. 2011;19(3):256-65.

Jordans, M. J. D., Luitel, N. P., Garman, E., Kohrt, B. A., Rathod, S. D., Shrestha, P., . . . Patel, V. (2019). Effectiveness of psychological treatments for depression and alcohol use disorder delivered by community-based counsellors: Two pragmatic randomised controlled trials within primary healthcare in Nepal. British Journal of Psychiatry, 215(2), 485-493.

Käll A, Bäck M, Fahlroth O, Ekeflod E, Lundberg A, Viberg N, et al. Internet-based therapist-supported interpersonal psychotherapy for depression: A randomized controlled trial. J Affect Disord. 2025;369:188-94.

Kamga, H., McCusker, J., Yaffe, M., Sewitch, M., Sussman, T., Strumpf, E., . . . Freeman, E. (2017). Self-care tools to treat depressive symptoms in patients with age-related eye disease: a randomized controlled clinical trial. Clinical & experimental ophthalmology, 45(4), 371-378.

Kanter JW, Santiago-Rivera AL, Santos MM, Nagy G, López M, Hurtado GD, et al. A randomized hybrid efficacy and effectiveness trial of behavioral activation for latinos with depression. Behavior Therapy. 2015;46(2):177-92.

Kay-Lambkin, F. J., Baker, A. L., Lewin, T. J., & Carr, V. J. (2009). Computer-based psychological treatment for comorbid depression and problematic alcohol and/or cannabis use: A randomized controlled trial of clinical efficacy. Addiction, 104(3), 378-388. doi:10.1111/j.1360-0443.2008.02444.x

Kelly JA, Murphy DA, Bahr GR, Kalichman SC, Morgan MG, Stevenson LY, et al. Outcome of cognitive-behavioral and support group brief therapies for depressed, HIV-infected persons. American Journal of Psychiatry. 1993;150(11):1679-86.

Kemmeren, L. L., van Schaik, A., Draisma, S., Kleiboer, A., Riper, H., & Smit, J. H. (2023). Effectiveness of Blended Cognitive Behavioral Therapy Versus Treatment as Usual for Depression in Routine Specialized Mental Healthcare: E-COMPARED Trial in the Netherlands. Cognitive Therapy and Research. doi:10.1007/s10608-023-10363-y

Kenter, R. M. F., Cuijpers, P., Beekman, A., & van Straten, A. (2016). Effectiveness of a Web-based guided self-help intervention for outpatients with a depressive disorder: Short-term results from a randomized controlled trial. Journal of medical Internet research, 18(3).

Kessler D, Lewis G, Kaur S, Wiles N, King M, Weich S, et al. Therapist-delivered Internet psychotherapy for depression in primary care: A randomised controlled trial. Lancet. 2009;374(9690):628-34.

Khazraee, H., Bakhtiari, M., Kianimoghadam, A. S., & Ghorbanikhah, E. (2023). The Effectiveness of Mindful Hypnotherapy on Depression, Self-Compassion, and Psychological Inflexibility in Females with Major Depressive Disorder: A Single-Blind, Randomized Clinical Trial. Int J Clin Exp Hypn, 71(1), 63-78. doi: 10.1080/00207144.2022.2160257

Khoshbooii, R., Hassan, S. A., Deylami, N., Muhamad, R., Engku Kamarudin, E. M., & Alareqe, N. A. (2021). Effects of Group and Individual Culturally Adapted Cognitive Behavioral Therapy on Depression and Sexual Satisfaction among Perimenopausal Women. Int J Environ Res Public Health, 18(14). https://doi.org/10.3390/ijerph18147711

Kim YH, Choi KS, Han K, Kim HW. A psychological intervention programme for patients with breast cancer under chemotherapy and at a high risk of depression: a randomised clinical trial. Journal of clinical nursing 2018; 27(3-4): 572-81.

King M, Sibbald B, Ward E, Bower P, Lloyd M, Gabbay M, et al. Randomised controlled trial of non-directive counselling, cognitive-behaviour therapy and usual general practitioner care in the management of depression as well as mixed anxiety and depression in primary care. Health Technology Assessment. 2000;4(19):1-83.

Kivi M, Eriksson MCM, Hange D, Petersson E-L, Vernmark K, Johansson B, et al. Internet-based therapy for mild to moderate depression in Swedish primary care: Short term results from the PRIM-NET randomized controlled trial. Cognitive Behaviour Therapy. 2014;43(4):289-98.

Koelen, J., Klein, A., Wolters, N., Bol, E., De Koning, L., Roetink, S., ... & Wiers, R. (2024). Web-Based, Human-Guided, or Computer-Guided Transdiagnostic Cognitive Behavioral Therapy in University Students With Anxiety and Depression: Randomized Controlled Trial. JMIR Mental Health, 11, e50503.

Korrelboom K, Maarsingh M, Huijbrechts I. Competitive memory training (COMET) for treating low self-esteem in patients with depressive disorders: A randomized clinical trial. Depression and anxiety. 2012;29(2):102-10.

Korte J, Bohlmeijer ET, Cappeliez P, Smit F, Westerhof GJ. Life review therapy for older adults with moderate depressive symptomatology: A pragmatic randomized controlled trial. Psychological Medicine. 2012;42(6):1163-73.

Kramer, J., Conijn, B., Oijevaar, P., & Riper, H. (2014). Effectiveness of a web-based solution-focused brief chat treatment for depressed adolescents and young adults: randomized controlled trial. J Med Internet Res, 16(5), e141. doi:10.2196/jmir.3261

Krämer, L. V., Grünzig, S. D., Baumeister, H., Ebert, D. D., & Bengel, J. (2021). Effectiveness of a Guided Web-Based Intervention to Reduce Depressive Symptoms before Outpatient Psychotherapy: A Pragmatic Randomized Controlled Trial. Psychother Psychosom, 90(4), 233-242.

Krämer, R., Köhne-Volland, L., Schumacher, A., & Köhler, S. (2022). Efficacy of a Web-Based Intervention for Depressive Disorders: Three-Arm Randomized Controlled Trial Comparing Guided and Unguided Self-Help With Waitlist Control. JMIR Form Res, 6(4), e34330. doi:10.2196/34330

Laidlaw K, Davidson K, Toner H, Jackson G, Clark S, Law J, et al. A randomised controlled trial of cognitive behaviour therapy vs treatment as usual in the treatment of mild to moderate late life depression. International Journal of Geriatric Psychiatry. 2008;23(8):843-50.

Lamers F, Jonkers CC, Bosma H, Kempen GI, Meijer JA, Penninx BW, et al. A minimal psychological intervention in chronically ill elderly patients with depression: A randomized trial. Psychotherapy and Psychosomatics. 2010;79(4):217-26.

Lamers SMA, Bohlmeijer ET, Korte J, Westerhof GJ. The efficacy of life-review as online-guided self-help for adults: A randomized trial. The Journals of Gerontology: Series B: Psychological Sciences and Social Sciences 2015; 70B(1): 24-34.

Landreville P, Bissonnette L. Effects of cognitive bibliotherapy for depressed older adults with a disability. Clinical Gerontologist. 1997;17(4):35-55.

Lappalainen P, Langrial S, Oinas-Kukkonen H, Tolvanen A, Lappalainen R. Web-based acceptance and commitment therapy for depressive symptoms with minimal support: A randomized controlled trial. Behavior modification. 2015;39(6):805-34.

Larcombe NA, Wilson PH. An evaluation of cognitive-behaviour therapy for depression in patients with multiple sclerosis. The British Journal of Psychiatry. 1984;145:366-71.

Layton, H., Campbell, M., Huh, K., Mansoor, A., Serrano-Lomelin, J., Brown, J. S. L., . . . Van Lieshout, R. J. (2025). Public Health Nurse-Delivered 1-Day Cognitive Behavioral Therapy-Based Workshops for Treating Postpartum Depression: A Pilot Randomized Controlled Trial. J Clin Psychiatry, 86(2). doi:https://doi.org/10.4088/jcp.24m15712

Lee, E., Han, Y., Cha, Y. J., Oh, J. H., Hwang, N. R., Seo, H. J., & Choi, K. H. (2021). Community-Based Multi-Site Randomized Controlled Trial of Behavioral Activation for Patients with Depressive Disorders [Article in Press]. Community mental health journal.

Lee, S. W., Seo, H. S., Choi, M., & Lee, S. J. (2025). A Randomized Controlled Trial of Group-Based Acceptance and Commitment Therapy for Major Depressive Disorder. Psychiatry Investigation, 22(11), 1300-1308.

Lemma A, Fonagy P. Feasibility study of a psychodynamic online group intervention for depression. Psychoanalytic Psychology. 2013;30(3):367-80.

Lenze, S., & Potts, M. (2017). Brief Interpersonal Psychotherapy for depression during pregnancy in a low-income population: a randomized controlled trial. Journal of Affective Disorders, 210, 151-157.

Lerner D, Adler DA, Rogers WH, Chang H, Greenhill A, Cymerman E, et al. A randomized clinical trial of a telephone depression intervention to reduce employee presenteeism and absenteeism. Psychiatric Services. 2015;66(6):570-7.

Leung SS, Lee AM, Chiang VC, Lam SK, Kuen YW, Wong DF. Culturally sensitive, preventive antenatal group cognitive-behavioural therapy for Chinese women with depression. Internationan Journal of Nursing Practice. 2013;19(Supp 1):28-37.

Leung S, Lee A, Wong D, Wong C, Leung K, Chiang V, et al. A brief group intervention using a cognitive-behavioural approach to reduce postnatal depressive symptoms: A randomised controlled trial. Hong Kong Medical Journal. 2016;22(1 Supplement 2).

Lexis MA, Jansen NW, Huibers MJ, Amelsvoort LG, Berkouwer A, Tjin ATG, et al. Prevention of long-term sickness absence and major depression in high-risk employees: A randomised controlled trial. Occupational and Environmental Medicine. 2011;68(6):400-7.

Liang, L., Feng, L., Zheng, X., Wu, Y., Zhang, C., & Li, J. (2021). Effect of dialectical behavior group therapy on the anxiety and depression of medical students under the normalization of epidemic prevention and control for the COVID-19 epidemic: a randomized study. Ann Palliat Med, 10(10), 10591-10599. https://doi.org/10.21037/apm-21-2466

Linde JA, Simon GE, Ludman EJ, Ichikawa LE, Operskalski BH, Arterburn D, et al. A randomized controlled trial of behavioral weight loss treatment versus combined weight loss/depression treatment among women with comorbid obesity and depression. Annals of Behavioral Medicine. 2011;41(1):119-30.

Liu ET-H, Chen W-L, Li Y-H, Wang CH, Mok TJ, Huang HS. Exploring the efficacy of cognitive bibliotherapy and a potential mechanism of change in the treatment of depressive symptoms among the Chinese: A randomized controlled trial. Cognitive Therapy and Research. 2009;33(5):449-61.

Liu, H., & Yang, Y. (2021). Effects of a psychological nursing intervention on prevention of anxiety and depression in the postpartum period: a randomized controlled trial [Article]. Annals of General Psychiatry, 20(1).

Liu W, Yuan J, Wu Y, Xu L, Wang X, Meng J, et al. A randomized controlled trial of mindfulness-based cognitive therapy for major depressive disorder in undergraduate students: Dose- response effect, inflammatory markers and BDNF. Psychiatry Research. 2024;331.

Liu YE, Lv J, Sun FZ, Liang JJ, Zhang YY, Chen J, et al. Effectiveness of group acceptance and commitment therapy in treating depression for acute stroke patients. Brain Behav. 2023;13(12):e3260.

Lloyd-Williams M, Shiels C, Ellis J, et al. Pilot randomised controlled trial of focused narrative intervention for moderate to severe depression in palliative care patients: DISCERN trial. Palliative medicine 2018; 32(1): 206-15.

Lök, N., Bademli, K., & Selçuk-Tosun, A. (2019). The effect of reminiscence therapy on cognitive functions, depression, and quality of life in Alzheimer patients: randomized controlled trial. International Journal of Geriatric Psychiatry, 34(1), 47-53. doi: 10.1002/gps.4980

Losada A, Márquez-González M, Romero-Moreno R, Mausbach BT, López J, Fernández-Fernández V, et al. Cognitive–behavioral therapy (CBT) versus acceptance and commitment therapy (ACT) for dementia family caregivers with significant depressive symptoms: Results of a randomized clinical trial. Journal of Consulting and Clinical Psychology. 2015;83(4):760-72.

Lovell K, Bower P, Richards D, Barkham Mi, Sibbald B, Roberts C, et al. Developing guided self-help for depression using the medical research council complex interventions framework: A description of the modelling phase and results of an exploratory randomised controlled trial. BMC psychiatry. 2008;8.

Lund, C., Schneider, M., Garman, E. C., Davies, T., Munodawafa, M., Honikman, S., . . . Susser, E. (2019). Task-sharing of psychological treatment for antenatal depression in Khayelitsha, South Africa: Effects on antenatal and postnatal outcomes in an individual randomised controlled trial. Behaviour Research and. doi:10.1016/j.brat.2019.103466

Lundgren JG, Dahlstrom O, Andersson G, Jaarsma T, Karner Kohler A, Johansson P. The effect of guided web-based cognitive behavioral therapy on patients with depressive symptoms and heart failure: A pilot randomized controlled trial. Journal of Medical Internet Research. 2016;18(8):e194.

Lustman PJ, Griffith LS, Freedland KE, Kissel SS, Clouse RE. Cognitive behavior therapy for depression in type 2 diabetes mellitus. A randomized, controlled trial. Annals of Internal Medicine. 1998;129(8):613-21.

Lynch DJ, Tamburrino MB, Nagel R. Telephone counseling for patients with minor depression: Preliminary findings in a family practice setting. The Journal of family practice. 1997;44(3):293-8.

Lynch D, Tamburrino M, Nagel R, Smith MK. Telephone-based treatment for family practice patients with mild depression. Psychological reports. 2004;94(3 Pt 1):785-92.

Lynch, T. R., Hempel, R. J., Whalley, B., Byford, S., Chamba, R., Clarke, P., . . . Russell, I. T. (2019). Refractory depression - mechanisms and efficacy of radically open dialectical behaviour therapy (RefraMED): findings of a randomised trial on benefits and harms. The British journal of psychiatry : the journal of mental science, 1-9. doi:10.1192/bjp.2019.53

MacLean S, Corsi DJ, Litchfield S, et al. Coach-facilitated web-based therapy compared with information about web-based resources in patients referred to secondary mental health care for depression: Randomized controlled trial. Journal of Medical Internet Research. 2020;22(6).

MacPherson H, Richmond S, Bland M, Brealey S, Gabe R, Hopton A, et al. Acupuncture and counselling for depression in primary care: A randomised controlled trial. PLoS medicine. 2013;10(9):e1001518.

Mahmoodi, M., Bakhtiyari, M., Masjedi Arani, A., Mohammadi, A., & Saberi Isfeedvajani, M. (2021). The comparison between CBT focused on perfectionism and CBT focused on emotion regulation for individuals with depression and anxiety disorders and dysfunctional perfectionism: A randomized controlled trial. Behavioural and Cognitive Psychotherapy, 49(4), 454-471.

Maina G, Forner F, Bogetto F. Randomized controlled trial comparing brief dynamic and supportive therapy with waiting list condition in minor depressive disorders. Psychotherapy and Psychosomatics. 2005;74(1):43-50.

Majidzadeh S, Mirghafourvand M, Farvareshi M, Yavarikia P. The effect of cognitive behavioral therapy on depression and anxiety of women with polycystic ovary syndrome: a randomized controlled trial. BMC Psychiatry. 2023;23(1):332.

Malouff JM, Lanyon RI, Schutte NS. Effectiveness of a brief group RET treatment for divorce-related dysphoria. Journal of Rational-Emotive and Cognitive-Behavior Therapy. 1988;6(3):162-71.

Mansour, N., Labib, N., Khalil, M., & Esmat, S. (2022). Brief Cognitive Behavioral Therapy for Patients with Comorbid Depression and Type 2 Diabetes in an Urban Primary Care Facility: Randomized Controlled Trial. Open Access Macedonian Journal of Medical Sciences, 10, 60-67. doi:10.3889/oamjms.2022.7883

Martin PR, Aiello R, Gilson K, Meadows G, Milgrom J, Reece J. Cognitive behavior therapy for comorbid migraine and/or tension-type headache and major depressive disorder: An exploratory randomized controlled trial. Behaviour Research and Therapy. 2015;73:8-18.

Matsuzaka, C., Wainberg, M., Norcini, P. A., Hoffmann, E., Coimbra, B., Braga, R., . . . Mello, M. (2017). Task shifting interpersonal counseling for depression: a pragmatic randomized controlled trial in primary care. BMC Psychiatry, 17(1)

McClay CA, Collins K, Matthews L, Haig C, McConnachie A, Morrison J, et al. A community-based pilot randomised controlled study of life skills classes for individuals with low mood and depression. BMC psychiatry. 2015;15:17.

McCusker, J., Jones, J. M., Li, M., Faria, R., Yaffe, M. J., Lambert, S. D., Ciampi, A., Belzile, E., & de Raad, M. (2021). CanDirect: Effectiveness of a Telephone-Supported Depression Self-Care Intervention for Cancer Survivors. J Clin Oncol, 39(10), 1150-1161.

McIndoo CC, File AA, Preddy T, Clark CG, Hopko DR. Mindfulness-based therapy and behavioral activation: A randomized controlled trial with depressed college students. Behaviour Research and Therapy. 2016;77:118-28.

McKee MD, Zayas LH, Fletcher J, Boyd RC, Nam SH. Results of an intervention to reduce perinatal depression among low-income minority women in community primary care. Journal of Social Service Research. 2006;32(4):63-81.

Merza D, Amani B, Savoy C, Babiy Z, Bieling PJ, Streiner DL, et al. Online peer-delivered group cognitive-behavioral therapy for postpartum depression: A randomized controlled trial. Acta Psychiatrica Scandinavica. 2023.

Michalak J, Schultze M, Heidenreich T, Schramm E. A randomized controlled trial on the efficacy of mindfulness-based cognitive therapy and a group version of cognitive behavioral analysis system of psychotherapy for chronically depressed patients. Journal of Consulting and Clinical Psychology. 2015;83(5):951-63.

Milgrom J, Negri LM, Gemmill AW, McNeil M, Martin PR. A randomized controlled trial of psychological interventions for postnatal depression. British Journal of Clinical Psychology. 2005;44(4):529-42.

Milgrom J, Holt CJ, Gemmill AW, Ericksen J, Leigh B, Buist A, et al. Treating postnatal depressive symptoms in primary care: A randomised controlled trial of GP management, with and without adjunctive counselling. BMC psychiatry. 2011;11:95.

Milgrom J, Holt C, Holt CJ, Ross J, Ericksen J, Gemmill AW. Feasibility study and pilot randomised trial of an antenatal depression treatment with infant follow-up. Archives of Women's Mental Health. 2015;18(5):717-30.

Milgrom J, Danaher BG, Gemmill AW, Holt C, Holt CJ, Seeley JR, et al. Internet cognitive behavioral therapy for women with postnatal depression: A randomized controlled trial of MumMoodBooster. Journal of Medical Internet Research. 2016;18(3):e54.

Milgrom, J., Danaher, B. G., Seeley, J. R., Holt, C. J., Holt, C., Ericksen, J., Tyler, M. S., Gau, J. M., & Gemmill, A. W. (2021). Internet and Face-to-face Cognitive Behavioral Therapy for Postnatal Depression Compared With Treatment as Usual: Randomized Controlled Trial of MumMoodBooster. J Med Internet Res, 23(12), e17185. https://doi.org/10.2196/17185

Miranda J, Chung JY, Green BL, Krupnick J, Siddique J, Revicki DA, et al. Treating depression in predominantly low-income young minority women: A randomized controlled trial. Jama. 2003;290(1):57-65.

Mohr DC, Likosky W, Bertagnolli A, Goodkin DE, Van Der Wende J, Dwyer P, et al. Telephone-administered cognitive-behavioral therapy for the treatment of depressive symptoms in multiple sclerosis. Journal of Consulting and Clinical Psychology. 2000;68(2):356-61.

Mohr DC, Carmody T, Erickson L, Jin L, Leader J. Telephone-administered cognitive behavioral therapy for veterans served by community-based outpatient clinics. Journal of Consulting and Clinical Psychology. 2011;79(2):261-5.

Mohr DC, Duffecy J, Ho J, Kwasny M, Cai X, Burns MN, et al. A randomized controlled trial evaluating a manualized TeleCoaching protocol for improving adherence to a web-based intervention for the treatment of depression. PLoS One. 2013;8(8):e70086.

Moldovan R, Cobeanu O, David D. Cognitive bibliotherapy for mild depressive symptomatology: Randomized clinical trial of efficacy and mechanisms of change. Clinical Psychology and Psychotherapy. 2013;20(6):482-93.

Montero-Marín, J., Araya, R., Pérez-Yus, M. C., Mayoral, F., Gili, M., Botella, C., . . . López-Del-Hoyo, Y. (2016). An internet-based intervention for depression in primary Care in Spain: a randomized controlled trial. Journal of medical Internet research, 18(8), e231.

Moon, J. R., Huh, J., Song, J., Kang, I. S., Park, S. W., Chang, S. A., . . . Han, J. S. (2021). The effects of rational emotive behavior therapy for depressive symptoms in adults with congenital heart disease. Heart Lung, 50(6), 906-913.

Mossey JM, Knott KA, Higgins M, Talerico K. Effectiveness of a psychosocial intervention, interpersonal counseling, for subdysthymic depression in medically ill elderly. The journals of gerontology Series A, Biological sciences and medical sciences. 1996;51(4):M172-8.

Mueller-Weinitschke C, Bengel J, Baumeister H, Krämer LV. Effects of a Web-Based Behavioral Activation Intervention on Depressive Symptoms, Activation, Motivation, and Volition: Results of a Randomized Controlled Trial. Psychotherapy and Psychosomatics. 2023.

Mukhtar, F., Oei, T. P., & Yaacob, M. (2011). Effectiveness of group cognitive behaviour therapy augmentation in reducing negative cognitions in the treatment of depression in Malaysia. ASEAN Journal of Psychiatry, 12(1), 50-65.

Mulcahy R, Reay RE, Wilkinson RB, Owen C. A randomised control trial for the effectiveness of group interpersonal psychotherapy for postnatal depression. Archives of Women's Mental Health. 2010;13(2):125-39.

Musa, Z. A., Soh, K. L., Mukhtar, F., Soh, K. Y., Oladele, T. O., & Soh, K. G. (2021). Effectiveness of mindfulness-based cognitive therapy among depressed individuals with disabilities in Nigeria: A randomized controlled trial. Psychiatry Res, 296, 113680.

Mynors-Wallis L, Gath D, Lloyd-Thomas A, Tomlinson D. Randomised controlled trial comparing problem solving treatment with amitriptyline and placebo for major depression in primary care. BMJ. 1995;310(6977):441-5.

Naeem F, Sarhandi I, Gul M, Khalid M, Aslam M, Anbrin A. A multicentre randomised controlled trial of a carer supervised culturally adapted cbt (cacbt) based self-help for depression in pakistan. Journal of Affective Disorders. 2013;156:224-7.

Naeem F, Gul M, Irfan M, Munshi T, Asif A, Rashid S, et al. Brief Culturally adapted CBT (CaCBT) for depression: A randomized controlled trial from Pakistan. Journal of Affective Disorders. 2015;177:101-7.

Nakagawa, A., Mitsuda, D., Sado, M., Abe, T., Fujisawa, D., Kikuchi, T., . . . Ono, Y. (2017). Effectiveness of supplementary cognitive-behavioral therapy for pharmacotherapy-resistant depression: A randomized controlled trial. Journal of clinical psychiatry, 78(8), 1126-1135

Nakao, S., Nakagawa, A., Oguchi, Y., Mitsuda, D., Kato, N., Nakagawa, Y., Tamura, N., Kudo, Y., Abe, T., Hiyama, M., Iwashita, S., Ono, Y., & Mimura, M. Web-Based Cognitive Behavioral Therapy Blended With Face-to-Face Sessions for Major Depression: Randomized Controlled Trial. J Med Internet Res, 20(9), e10743.

Nakimuli-Mpungu E, Wamala K, Okello J, Alderman S, Odokonyero R, Mojtabai R, et al. Group support psychotherapy for depression treatment in people with HIV/AIDS in northern Uganda: A single-centre randomised controlled trial. The Lancet HIV. 2015;2(5):e190-e9.

Nasrin, F., Rimes, K., Reinecke, A., Rinck, M., & Barnhofer, T. (2017). Effects of Brief Behavioural Activation on Approach and Avoidance Tendencies in Acute Depression: preliminary Findings. Behavioural and Cognitive Psychotherapy, 45(1), 58-72.

Neugebauer R, Kline J, Markowitz JC, Bleiberg KL, Baxi L, Rosing MA, et al. Pilot randomized controlled trial of interpersonal counseling for subsyndromal depression following miscarriage. Journal of Clinical Psychiatry. 2006;67(8):1299-304.

Newby, J. M., et al. (2013). "Internet cognitive behavioural therapy for mixed anxiety and depression: a randomized controlled trial and evidence of effectiveness in primary care." Psychol Med 43(12): 2635-2648.

Newby JM, Lang T, Werner-Seidler A, Holmes E, Moulds ML. Alleviating distressing intrusive memories in depression: A comparison between computerised cognitive bias modification and cognitive behavioural education. Behaviour Research and Therapy. 2014;56:60-7.

Newby, J., Robins, L., Wilhelm, K., Smith, J., Fletcher, T., Gillis, I., . . . Andrews, G. (2017). Web-Based Cognitive Behavior Therapy for Depression in People With Diabetes Mellitus: a Randomized Controlled Trial. Journal of medical Internet research, 19(5), e157.

Nezu AM. Efficacy of a social problem-solving therapy approach for unipolar depression. Journal of Consulting and Clinical Psychology. 1986;54(2):196-202.

Nezu AM, Perri MG. Social problem-solving therapy for unipolar depression: An initial dismantling investigation. Journal of Consulting and Clinical Psychology. 1989;57(3):408-13.

Ngai FW, Wong P-C, Leung KY, Chau PH, Chung KF. The effect of telephone-based cognitive-behavioral therapy on postnatal depression: A randomized controlled trial. Psychotherapy and Psychosomatics. 2015;84(5):294-303.

Niedermoser DW, Kalak N, Kiyhankhadiv A, et al. Workplace-Related Interpersonal Group Psychotherapy to Improve Life at Work in Individuals With Major Depressive Disorders: a Randomized Interventional Pilot Study. Frontiers in psychiatry. 2020;11.

Nobis S, Lehr D, Ebert DD, Baumeister H, Snoek F, Riper H, et al. Efficacy of a web-based intervention with mobile phone support in treating depressive symptoms in adults with type 1 and type 2 diabetes: A randomized controlled trial. Diabetes care. 2015;38(5):776-83.

Nollett CL, Bray N, Bunce C, Casten RJ, Edwards RT, Hegel MT, et al. Depression in Visual Impairment Trial (DEPVIT): A randomized clinical trial of depression treatments in people with low vision. Investigative Ophthalmology and Visual Science. 2016;57(10): 4247-54.

Noone, D., Payne, J., Stott, J., Aguirre, E., Patel-Palfreman, M. M., Stoner, C., . . . Spector, A. (2022). The Feasibility of a Mindfulness Intervention for Depression in People with Mild Dementia: A Pilot Randomized Controlled Trial. Clinical gerontologist, 1-

Northwood AK, Vukovich MM, Beckman A, et al. Intensive psychotherapy and case management for Karen refugees with major depression in primary care: a pragmatic randomized control trial. BMC Fam Pract. 2020;21(1):17.

Nyström, M., Stenling, A., Sjöström, E., Neely, G., Lindner, P., HassmÈn, P., . . . Carlbring, P. (2017). Behavioral activation versus physical activity via the internet: a randomized controlled trial. Journal of Affective Disorders, 215, 85-93.

O'Hara MW, Stuart S, Gorman LL, Wenzel A. Efficacy of interpersonal psychotherapy for postpartum depression. Archives of General Psychiatry. 2000;57(11):1039-45.

O'Mahen H, Himle JA, Fedock G, Henshaw E, Flynn H. A pilot randomized controlled trial of cognitive behavioral therapy for perinatal depression adapted for women with low incomes. Depression and anxiety. 2013;30(7):679-87.

O'Neil A, Taylor B, Sanderson K, Cyril S, Chan B, Hawkes AL, et al. Efficacy and feasibility of a tele-health intervention for acute coronary syndrome patients with depression: Results of the "MoodCare" randomized controlled trial. Annals of Behavioral Medicine. 2014;48(2):163-74.

Oehler C, Görges F, Rogalla M, Rummel-Kluge C, Hegerl U. Efficacy of a Guided Web-Based Self-Management Intervention for Depression or Dysthymia: Randomized Controlled Trial With a 12-Month Follow-Up Using an Active Control Condition. J Med Internet Res. 2020;22(7):e15361.

Okeke NM, Onah BN, Ekwealor NE, Ekwueme SC, Ezugwu JO, Edeh EN, et al. Effect of a randomized group intervention for depression among Nigerian pre-service adult education teachers. Medicine (Baltimore). 2023;102(27):e34159.

Olukolade, O., & Osinowo, H. (2017). Efficacy of Cognitive Rehabilitation Therapy on Poststroke Depression among Survivors of First Stroke Attack in Ibadan, Nigeria. Behavioural Neurology, 2017

Omidi A, Mohammadkhani P, Mohammadi A, Zargar F. Comparing mindfulness based cognitive therapy and traditional cognitive behavior therapy with treatments as usual on reduction of major depressive disorder symptoms. Iranian Red Crescent Medical Journal. 2013;15(2):142-6.

Onuigbo, L. N., Eseadi, C., Ebifa, S., Ugwu, U. C., Onyishi, C. N., & Oyeoku, E. K. (2019). Effect of rational emotive behavior therapy program on depressive symptoms among university students with blindness in Nigeria. Journal of Rational-Emotive & Cognitive-Behavior Therapy, 37(1), 17-38. doi:10.1007/s10942-018-0297-3

Otu MS, Ebizie EN, Otu FM, Eseadi C. Efficacy of a counselling video blog intervention from YouTube on depression reduction among primary school teachers. Psychological Studies. 2023;68(3):335-41.

Pace TM, Dixon DN. Changes in depressive self-schemata and depressive symptoms following cognitive therapy. Journal of Counseling Psychology. 1993;40(3):288.

Pagoto S, Schneider KL, Whited MC, Oleski JL, Merriam P, Appelhans B, et al. Randomized controlled trial of behavioral treatment for comorbid obesity and depression in women: The Be Active Trial. International Journal of Obesity. 2013;37(11):1427-34.

Pan, W. L., Lin, L. C., Kuo, L. Y., Chiu, M. J., & Ling, P. Y. (2023). Effects of a prenatal mindfulness program on longitudinal changes in stress, anxiety, depression, and mother-infant bonding of women with a tendency to perinatal mood and anxiety disorder: a randomized controlled trial. BMC Pregnancy Childbirth, 23(1), 547.

Pan, J. Y., & Rafi, J. (2025). Culturally Adapted Guided Internet-Based Cognitive Behavioral Therapy for Hong Kong People With Depressive Symptoms: Randomized Controlled Trial. Journal of Medical Internet Research, 27.

Patel, V., Weobong, B., Weiss, H., Anand, A., Bhat, B., Katti, B., . . . Fairburn, C. (2017). The Healthy Activity Program (HAP), a lay counsellor-delivered brief psychological treatment for severe depression, in primary care in India: a randomised controlled trial. Lancet (london, england), 389(10065), 176-185.

Pecheur DR, Edwards KJ. A comparison of secular and religious versions of cognitive therapy with depressed Christian college students. Journal of Psychology and Theology. 1984.

Peden AR, Hall LA, Rayens MK, Beebe LL. Reducing negative thinking and depressive symptoms in college women. Journal of Nursing Scholarship. 2000;32(2):145-51.

Pellas, J., Renner, F., Ji, J. L., & Damberg, M. (2022). Telephone-based behavioral activation with mental imagery for depression: A pilot randomized clinical trial in isolated older adults during the Covid-19 pandemic. Int J Geriatr Psychiatry, 37(1).

Penckofer SM, Ferrans C, Mumby P, Byrn M, Emanuele MA, Harrison PR, et al. A psychoeducational intervention (SWEEP) for depressed women with diabetes. Annals of Behavioral Medicine. 2012;44(2):192-206.

Perini S, Titov N, Andrews G. Clinician-assisted Internet-based treatment is effective for depression: Randomized controlled trial. Australian and New Zealand Journal of Psychiatry. 2009;43(6):571-8.

Petersen I, Hanass Hancock J, Bhana A, Govender K. A group-based counselling intervention for depression comorbid with HIV/AIDS using a task shifting approach in South Africa: A randomized controlled pilot study. Journal of Affective Disorders. 2014;158:78-84.

Pibernik-Okanovic M, Begic D, Ajdukovic D, Andrijasevic N, Metelko Z. Psychoeducation versus treatment as usual in diabetic patients with subthreshold depression: Preliminary results of a randomized controlled trial. Trials. 2009;10:78.

Pibernik-Okanović M, Hermanns N, Ajduković D, Kos J, Prašek M, Šekerija M, Lovrenčić MV. Does treatment of subsyndromal depression improve depression-related and diabetes-related outcomes? A randomised controlled comparison of psychoeducation, physical exercise and enhanced treatment as usual. Trials. 2015 Jul 15;16:305.

Piers, R. J., Farchione, T. J., Wong, B., Rosellini, A. J., & Cronin-Golomb, A. (2023). Telehealth Transdiagnostic Cognitive Behavioral Therapy for Depression in Parkinson's Disease: A Pilot Randomized Controlled Trial. Movement Disorders Clinical Practice, 10(1), 79-85. doi:10.1002/mdc3.13587

Pihlaja S, Lahti J, Lipsanen JO, et al. Scheduled Telephone Support for Internet Cognitive Behavioral Therapy for Depression in Patients at Risk for Dropout: Pragmatic Randomized Controlled Trial. J Med Internet Res. 2020;22(7):e15732.

Pinniger, R., Brown, R. F., Thorsteinsson, E. B., & McKinley, P. (2012). Argentine tango dance compared to mindfulness meditation and a waiting-list control: A randomised trial for treating depression. Complement Ther Med, 20(6), 377-384.

Poleshuck EL, Gamble SA, Bellenger K, Lu N, Tu X, Sorensen S, et al. Randomized controlled trial of interpersonal psychotherapy versus enhanced treatment as usual for women with co-occurring depression and pelvic pain. Journal of psychosomatic research. 2014;77(4):264-72.

Pot AM, Bohlmeijer ET, Onrust S, Melenhorst A-S, Veerbeek M, De Vries W. The impact of life review on depression in older adults: A randomized controlled trial. International Psychogeriatrics. 2010;22(4):572-81.

Pots WTM, Meulenbeek PAM, Veehof MM, Klungers J, Bohlmeijer ET. The efficacy of mindfulness-based cognitive therapy as a public mental health intervention for adults with mild to moderate depressive symptomatology: A randomized controlled trial. PLoS One. 2014;9(10).

Pots WT, Fledderus M, Meulenbeek PA, ten Klooster PM, Schreurs KM, Bohlmeijer ET. Acceptance and commitment therapy as a web-based intervention for depressive symptoms: Randomised controlled trial. The British Journal of Psychiatry. 2016;208(1): 69-77.

Pott, S. L., Kellett, S., Green, S., Daughters, S., & Delgadillo, J. (2022). Behavioral activation for depression delivered by drug and alcohol treatment workers: A pilot randomized controlled trial. J Subst Abuse Treat, 139, 108769.

Power MJ, Freeman C. A randomized controlled trial of IPT versus CBT in primary care: With some cautionary notes about handling missing values in clinical trials. Clinical Psychology and Psychotherapy. 2012;19(2):159-69.

Powers, A., Lathan, E. C., Dixon, H. D., Mekawi, Y., Hinrichs, R., Carter, S., . . . Kaslow, N. J. (2023). Primary care-based mindfulness intervention for posttraumatic stress disorder and depression symptoms among Black adults: A pilot feasibility and acceptability randomized controlled trial. Psychol Trauma, 15(5), 858-867.

Prabhu S, George LS, Guruvare S, Noronha JA, Jose TT, Nayak BS, et al. Effectiveness of psychosocial education program on postnatal depression, stress, and perceived maternal parenting self-efficacy among pregnant women in South India. Patient Education and Counseling. 2025;130.

Prendergast J, Austin MP. Early childhood nurse-delivered cognitive behavioural counselling for post-natal depression. Australasian Psychiatry. 2001;9(3):255-9.

Preschl B, Maercker A, Wagner B, Forstmeier S, Banos RM, Alcaniz M, et al. Life-review therapy with computer supplements for depression in the elderly: A randomized controlled trial. Aging and Mental Health. 2012;16(8):964-74.

Propst LR, Ostrom R, Watkins P, Dean T, Mashburn D. Comparative efficacy of religious and nonreligious cognitive-behavioral therapy for the treatment of clinical depression in religious individuals. Journal of Consulting and Clinical Psychology. 1992;60(1):94-103.

Psaros, C., Stanton, A. M., Raggio, G. A., Mosery, N., Goodman, G. R., Briggs, E. S., . . . Safren, S. A. (2022). Optimizing PMTCT Adherence by Treating Depression in Perinatal Women with HIV in South Africa: A Pilot Randomized Controlled Trial. International journal of behavioral medicine. doi:10.1007/s12529-022-10071-z

Psarraki, E. E., Bacopoulou, F., Panagoulias, E., Michou, M., Pelekasis, P., Artemiadis, A., . . . Darviri, C. (2021). The effects of Pythagorean Self-Awareness Intervention on patients with major depressive disorder: A pilot randomized controlled trial. J Psychiatr Res, 138, 326-334. doi:10.1016/j.jpsychires.2021.03.067

Puckering C, McIntosh E, Hickey A, Longford J. Mellow Babies: A group intervention for infants and mothers experiencing postnatal depression. Counselling Psychology Review. 2010;25(1):28-38.

Pugh, N. E., Hadjistavropoulos, H. D., & Dirkse, D. (2016). A Randomised Controlled Trial of Therapist-Assisted, Internet-Delivered Cognitive Behavior Therapy for Women with Maternal Depression. PLoS ONE, 11(3), e0149186.

Qiu J, Chen W, Gao X, Xu Y, Tong H, Yang M, et al. A randomized controlled trial of group cognitive behavioral therapy for Chinese breast cancer patients with major depression. Journal of psychosomatic obstetrics and gynaecology. 2013;34(2):60-7.

Raevuori, A., Vahlberg, T., Korhonen, T., Hilgert, O., Aittakumpu-Hyden, R., & Forman-Hoffman, V. (2021). A therapist-guided smartphone app for major depression in young adults: A randomized clinical trial. J Affect Disord, 286, 228-238.

Raji Lahiji, M., Sajadian, A., Haghighat, S., Zarrati, M., Dareini, H., Raji Lahiji, M., & Razmpoosh, E. (2022). Effectiveness of logotherapy and nutrition counseling on psychological status, quality of life, and dietary intake among breast cancer survivors with depressive disorder: a randomized clinical trial. Supportive Care in Cancer. doi:10.1007/s00520-022-07237-6

Ramsauer, B. S., Michaelsen, J. L., & Zitzmann, S. (2026). The effects of mentalization and sensitivity of postpartum depression mothers after the circle of security-intensive intervention on the social-emotional development of three-year-old children. J Affect Disord, 396, 120838.

Ransom D, Heckman TG, Anderson T, Garske J, Holroyd K, Basta T. Telephone-delivered, interpersonal psychotherapy for HIV-infected rural persons with depression: A pilot trial. Psychiatric Services. 2008;59(8):871-7.

Raue, P. J., Sirey, J. A., Dawson, A., Berman, J., & Bruce, M. L. (2019). Lay-delivered behavioral activation for depressed senior center clients: Pilot RCT. International journal of geriatric psychiatry, 34(11), 1715-1723. doi:10.1002/gps.5186

Raya-Tena, A., Fernández-San-Martin, M. I., Martin-Royo, J., Casañas, R., Sauch-Valmaña, G., Cols-Sagarra, C., . . . Jiménez-Herrera, M. F. (2021). Effectiveness of a Psychoeducational Group Intervention Carried Out by Nurses for Patients with Depression and Physical Comorbidity in Primary Care: Randomized Clinical Trial. Int J Environ Res Public Health, 18(6).

Rehm LP, Kornblith SJ, O'Hara MW, Lamparski DM, Romano JM, Volkin JI. An evaluation of major components in a self-control therapy program for depression. Behavior modification. 1981;5(4): 459-89.

Reins, J. A., Boß, L., Lehr, D., Berking, M., & Ebert, D. D. (2019). The more I got, the less I need? Efficacy of Internet-based guided self-help compared to online psychoeducation for major depressive disorder. Journal of, 246, 695-705.

Richards D, Timulak L, O'Brien E, Hayes C, Vigano N, Sharry J, et al. A randomized controlled trial of an internet-delivered treatment: Its potential as a low-intensity community intervention for adults with symptoms of depression. Behaviour Research and Therapy. 2015;75:20-31.

Rief W, Bleichhardt G, Dannehl K, Euteneuer F, Wambach K. Comparing the Efficacy of CBASP with Two Versions of CBT for Depression in a Routine Care Center: a Randomized Clinical Trial. Psychotherapy and psychosomatics 2018;

Ritvo, P., et al. (2021). "Online Mindfulness-Based Cognitive Behavioral Therapy Intervention for Youth With Major Depressive Disorders: Randomized Controlled Trial." J Med Internet Res 23(3): e24380.

Rizvi SJ, Zaretsky A, Schaffer A, Levitt A. Is immediate adjunctive CBT more beneficial than delayed CBT in treating depression?: A Pilot Study. Journal of Psychiatric Practice. 2015;21(2):107-13.

Rohan KJ, Roecklein KA, Lindsey KT, Johnson LG, Lippy RD, Lacy TJ, et al. A randomized controlled trial of cognitive-behavioral therapy, light therapy, and their combination for seasonal affective disorder. Journal of Consulting and Clinical Psychology. 2007;75(3):489-500.

Rohde, P., Stice, E., Shaw, H., & Gau, J. M. (2014). Cognitive-behavioral group depression prevention compared to bibliotherapy and brochure control: Nonsignificant effects in pilot effectiveness trial with college students. Behaviour Research and Therapy, 55(1), 48-53.

Rohde P, Stice E, Shaw H, Gau JM. Pilot trial of a dissonance-based cognitive-behavioral group depression prevention with college students. Behaviour Research and Therapy. 2016;82:21-7.

Rohricht F, Papadopoulos N, Priebe S. An exploratory randomized controlled trial of body psychotherapy for patients with chronic depression. Journal of Affective Disorders. 2013;151(1):85-91.

Ross M, Scott M. An evaluation of the effectiveness of individual and group cognitive therapy in the treatment of depressed patients in an inner city health centre. The Journal of the Royal College of General Practitioners. 1985;35(274):239-42.

Rude SS. Relative benefits of assertion or cognitive self-control treatment for depression as a function of proficiency in each domain. Journal of Consulting and Clinical Psychology. 1986;54(3):390-4.

Russell, A., Gaunt, D. M., Cooper, K., Barton, S., Horwood, J., Kessler, D., . . . et al. (2019). The feasibility of low-intensity psychological therapy for depression co-occurring with autism in adults: the Autism Depression Trial (ADEPT) â€" a pilot randomised controlled trial. Autism. doi: 10.1177/1362361319889272

Ruwaard J, Schrieken B, Schrijver M, Broeksteeg J, Dekker J, Vermeulen H, et al. Standardized web-based cognitive behavioural therapy of mild to moderate depression: A randomized controlled trial with a long-term follow-up. Cognitive Behaviour Therapy. 2009;38(4):206-21.

Ruzickova, T., Carson, J., Murphy, S., & Harmer, C. (2021). P.230 Effects of online behavioural activation on depression during covid-19. European neuropsychopharmacology, 44, S36-S37. doi: 10.1016/j.euroneuro.2021.01.059

Sadler, P., McLaren, S., Klein, B., Harvey, J., & Jenkins, M. (2018). Cognitive behavior therapy for older adults with insomnia and depression: a randomized controlled trial in community mental health services. Sleep, 41(8) (no pagination).

Safren SA, O'Cleirigh C, Tan JY, Raminani SR, Reilly LC, Otto MW, et al. A randomized controlled trial of cognitive behavioral therapy for adherence and depression (CBT-AD) in HIV-infected individuals. Health Psychology. 2009;28(1):1-10.

Safren SA, Gonzalez JS, Wexler DJ, Psaros C, Delahanty LM, Blashill AJ, et al. A randomized controlled trial of cognitive behavioral therapy for adherence and depression (CBT-AD) in patients with uncontrolled type 2 diabetes. Diabetes care. 2014;37(3):625-33.

Safren SA, Bedoya CA, O'Cleirigh C, Biello KB, Pinkston MM, Stein MD, et al. Cognitive behavioural therapy for adherence and depression in patients with HIV: A three-arm randomised controlled trial. The Lancet HIV. 2016;3(11):e529-e38.

Safren, S. A., O'Cleirigh, C., Andersen, L. S., Magidson, J. F., Lee, J. S., Bainter, S. A., . . . Joska, J. A. (2021). Treating depression and improving adherence in HIV care with task-shared cognitive behavioural therapy in Khayelitsha, South Africa: a randomized controlled trial. J Int AIDS Soc, 24(10), e25823. doi:10.1002/jia2.25823

Şahin Tokatlıoğlu, T., & Oflaz, F. (2025). The effect of interpersonal psychotherapy on depression, emotion regulation, and perceived social support in breast cancer patients at risk of depression: A randomized controlled trial. Eur J Oncol Nurs, 79, 103042.

Salamanca-Sanabria A, Richards D, Timulak L, et al. A culturally adapted cognitive behavioral internet-delivered intervention for depressive symptoms: Randomized controlled trial. JMIR Mental Health. 2020;7(1).

Saloheimo HP, Markowitz J, Saloheimo TH, Laitinen JJ, Sundell J, Huttunen MO, et al. Psychotherapy effectiveness for major depression: A randomized trial in a Finnish community. BMC psychiatry. 2016;16:131.

Sanabria-Mazo JP, Colomer-Carbonell A, Borràs X, Castaño-Asins JR, McCracken LM, Montero-Marin J, et al. Efficacy of Videoconference Group Acceptance and Commitment Therapy (ACT) and Behavioral Activation Therapy for Depression (BATD) for Chronic Low Back Pain (CLBP) Plus Comorbid Depressive Symptoms: A Randomized Controlled Trial (IMPACT Study). J Pain. 2023;24(8):1522-40.

Sander LB, Paganini S, Terhorst Y, et al. Effectiveness of a Guided Web-Based Self-help Intervention to Prevent Depression in Patients with Persistent Back Pain: The PROD-BP Randomized Clinical Trial. JAMA Psychiatry. 2020;77(10):1001-1011.

Savard J, Simard S, Giguere I, Ivers H, Morin CM, Maunsell E, et al. Randomized clinical trial on cognitive therapy for depression in women with metastatic breast cancer: Psychological and immunological effects. Palliative and Supportive Care. 2006;4(3):219-37.

Savari, Y., Mohagheghi, H., & Petrocchi, N. (2021). A preliminary investigation on the effectiveness of compassionate mind training for students with major depressive disorder: A randomized controlled trial. Mindfulness, 12(5), 1159-1172.

Schlicker S, Baumeister H, Buntrock C, et al. A web- And mobile-based intervention for comorbid, recurrent depression in patients with chronic back pain on sick leave (get.back): Pilot randomized controlled trial on feasibility, user satisfaction, and effectiveness. JMIR Mental Health. 2020;7(4).

Schmidt MM, Miller WR. Amount of therapist contact and outcome in a multidimensional depression treatment program. Acta Psychiatrica Scandinavica. 1983;67(5):319-32.

Schramm E, Mack S, Thiel N, Jenkner C, Elsaesser M, Fangmeier T. Interpersonal psychotherapy vs treatment as usual for major depression related to work stress: A pilot randomized controlled study. Frontiers in Psychiatry. 2020;11.

Schulberg HC, Block MR, Madonia MJ, Scott CP, Rodriguez E, Imber SD, et al. Treating major depression in primary care practice. Eight-month clinical outcomes. Archives of General Psychiatry. 1996;53(10):913-9.

Schuster, R., Leitner, I., Carlbring, P., & Laireiter, A.-R. (2017). Exploring blended group interventions for depression: randomised controlled feasibility study of a blended computer- and multimedia-supported psychoeducational group intervention for adults with depressive symptoms. Internet Interventions, 8, 63-71.

Schuster, R., Fischer, E., Jansen, C., Napravnik, N., Rockinger, S., Steger, N., & Laireiter, A.-R. (2022). Blending Internet-based and tele group treatment: Acceptability, effects, and mechanisms of change of cognitive behavioral treatment for depression. Internet Interventions, 29, 100551. doi:https://doi.org/10.1016/j.invent.2022.100551

Scogin F, Hamblin D, Beutler L. Bibliotherapy for depressed older adults: A self-help alternative. The Gerontologist. 1987;27(3):383-7.

Scogin F, Jamison C, Gochneaur K. Comparative efficacy of cognitive and behavioral bibliotherapy for mildly and moderately depressed older adults. Journal of Consulting and Clinical Psychology. 1989;57(3):403-7.

Scogin F, Lichstein K, DiNapoli EA, et al. Effects of integrated telehealth-delivered cognitive-behavioral therapy for depression and insomnia in rural older adults. Journal of psychotherapy integration 2018; 28(3): 292-309.

Scott MJ, Stradling SG. Group cognitive therapy for depression produces clinically significant reliable change in community-based settings. Behavioural Psychotherapy. 1990;18(01):1-19.

Scott AI, Freeman CP. Edinburgh primary care depression study: Treatment outcome, patient satisfaction, and cost after 16 weeks. BMJ. 1992;304(6831):883-7.

Scott C, Tacchi MJ, Jones R, Scott J. Acute and one-year outcome of a randomised controlled trial of brief cognitive therapy for major depressive disorder in primary care. The British Journal of Psychiatry. 1997;171:131-4.

Segal, Z. V., Dimidjian, S., Beck, A., Boggs, J. M., Vanderkruik, R., Metcalf, C. A., . . . Levy, J. (2020). Outcomes of Online Mindfulness-Based Cognitive Therapy for Patients with Residual Depressive Symptoms: A Randomized Clinical Trial. JAMA Psychiatry, 77(6), 563-573. doi:10.1001/jamapsychiatry.2019.4693

Segre LS, Brock RL, O'Hara MW. Depression treatment for impoverished mothers by point-of-care providers: A randomized controlled trial. Journal of Consulting and Clinical Psychology. 2015;83(2):314-24.

Selmi PM, Klein MH, Greist JH, Sorrell SP, Erdman HP. Computer-administered cognitive-behavioral therapy for depression. American Journal of Psychiatry. 1990;147(1):51-6.

Serfaty MA, Haworth D, Blanchard M, Buszewicz M, Murad S, King M. Clinical effectiveness of individual cognitive behavioral therapy for depressed older people in primary care: A randomized controlled trial. Archives of General Psychiatry. 2009;66(12):1332-40.

Serfaty, M., King, M., Nazareth, I., Moorey, S., Aspden, T., Tookman, A., . . . Jones, L. (2019). Manualised cognitive and behavioural therapy in treating depression in advanced cancer: the CanTalk RCT. Health Technology Assessment, 23(19), 1-106.

Serrano Selva JP, Latorre Postigo JM, Ros Segura L, Navarro Bravo B, Aguilar Corcoles MJ, Nieto Lopez M, et al. Life review therapy using autobiographical retrieval practice for older adults with clinical depression. Psicothema. 2012;24(2):224-9.

Serrano JP, Latorre JM, Gatz M, Montanes J. Life review therapy using autobiographical retrieval practice for older adults with depressive symptomatology. Psychology and Aging. 2004;19(2):270-7.

Seshadri, A., Fuller-Tyszkiewicz, M., Harper, L., Clark, M. M., Singh, B., Chesak, S., . . . Frye, M. A. (2025). Randomized controlled trial of stress management and resiliency training for depression (SMART-D)-pilot study. PLOS ONE, 20(8 August).

Shan, Q., Xinxin, S., Zhijuan, X., Rongjing, D., & Minjie, Z. (2022). Effects of Cognitive Behavior Therapy on Depression, Illness Perception, and Quality of Life in Atrial Fibrillation Patients. Frontiers in Psychiatry, 13. doi:10.3389/fpsyt.2022.830363

Shaw BF. Comparison of cognitive therapy and behavior therapy in the treatment of depression. Journal of Consulting and Clinical Psychology. 1977;45(4):543.

Sheeber LB, Seeley JR, Feil EG, Davis B, Sorensen E, Kosty DB, et al. Development and pilot evaluation of an Internet-facilitated cognitive-behavioral intervention for maternal depression. Journal of Consulting and Clinical Psychology. 2012;80(5):739-49.

Sheeber, L., Feil, E., Seeley, J., Leve, C., Gau, J., Davis, B., . . . Allan, S. (2017). Mom-net: evaluation of an internet-facilitated cognitive behavioral intervention for low-income depressed mothers. Journal of Consulting and Clinical Psychology, 85(4), 355-366.

Sheline, Y. I., Thase, M. E., Hembree, E. A., Balderston, N. L., Nitchie, F. J., Batzdorf, A. S., . . . Lynch, K. G. (2025). Neuroimaging changes in major depression with brief computer-assisted cognitive behavioral therapy compared to waitlist. Molecular Psychiatry. doi:https://doi.org/10.1038/s41380-025-02945-x

Shih, V. W. Y., Chan, W. C., Tai, O. K., Wong, H. L., Cheng, C. P. W., & Wong, C. S. M. (2021). Mindfulness-Based Cognitive Therapy for Late-Life Depression: a Randomised Controlled Trial. East Asian Arch Psychiatry, 31(2), 27-35.

Simoni JM, Wiebe JS, Sauceda JA, Huh D, Sanchez G, Longoria V, et al. A preliminary RCT of CBT-AD for adherence and depression among HIV-positive Latinos on the U.S.-Mexico border: The Nuevo Dia study. AIDS and behavior. 2013;17(8):2816-29.

Simpson S, Corney R, Beecham J. A randomized controlled trial to evaluate the effectiveness and cost-effectiveness of psychodynamic counselling for general practice patients with chronic depression. Psychological Medicine. 2003;33(2):229-39.

Simson U, Nawarotzky U, Friese G, Porck W, Schottenfeld-Naor Y, Hahn S, et al. Psychotherapy intervention to reduce depressive symptoms in patients with diabetic foot syndrome. Diabetic Medicine. 2008;25(2):206-12.

Sinniah, A., Oei, T., Maniam, T., & Subramaniam, P. (2017). Positive effects of Individual Cognitive Behavior Therapy for patients with unipolar mood disorders with suicidal ideation in Malaysia: a randomised controlled trial. Psychiatry research, 254, 179-189.

Sirey, J. A., Solomonov, N., Guillod, A., Zanotti, P., Lee, J., Soliman, M., & Alexopoulos, G. S. (2021). PROTECT: a novel psychotherapy for late-life depression in elder abuse victims. Int Psychogeriatr, 33(5), 521-525.

Sloane R, Staples F, Schneider L. Interpersonal therapy versus nortriptyline for depression in the elderly. Clinical and Pharmacological Studies in Psychiatric Disorders. 1985:344-6.

Smit A, Kluiter H, Conradi HJ, Meer K, Tiemens BG, Jenner JA, et al. Short-term effects of enhanced treatment for depression in primary care: Results from a randomized controlled trial. Psychological Medicine. 2006;36(1):15-26.

Smith, J., Newby, J. M., Burston, N., Murphy, M. J., Michael, S., Mackenzie, A., . . . Andrews, G. (2017). Help from home for depression: A randomised controlled trial comparing internet-delivered cognitive behaviour therapy with bibliotherapy for depression. Internet Interventions, 9, 25-37.

Solis, E. C., Carlier, I. V. E., Kamminga, N. G. A., Giltay, E. J., & van Hemert, A. M. (2024). The clinical effectiveness of a self-management intervention for patients with persistent depressive disorder and their partners/caregivers: results from a multicenter, pragmatic randomized controlled trial. Trials, 25(1). doi:https://doi.org/10.1186/s13063-024-08033-9

Songprakun W, McCann TV. Evaluation of a cognitive behavioural self-help manual for reducing depression: A randomized controlled trial. Journal of psychiatric and mental health nursing. 2012;19(7):647-53.

Spek V, Nyklicek I, Smits N, Cuijpers P, Riper H, Keyzer J, et al. Internet-based cognitive behavioural therapy for subthreshold depression in people over 50 years old: A randomized controlled clinical trial. Psychological Medicine. 2007;37(12):1797-806.

Spinelli MG, Endicott J. Controlled clinical trial of interpersonal psychotherapy versus parenting education program for depressed pregnant women. American Journal of Psychiatry. 2003;160(3):555-62.

Spinelli MG, Endicott J, Leon AC, Goetz RR, Kalish RB, Brustman LE, et al. A controlled clinical treatment trial of interpersonal psychotherapy for depressed pregnant women at 3 New York City sites. Journal of Clinical Psychiatry. 2013;74(4):393-9.

Spruill, T. M., Friedman, D., Diaz, L., Butler, M. J., Goldfeld, K. S., O'Kula, S., . . . Devinsky, O. (2021). Telephone-based depression self-management in Hispanic adults with epilepsy: a pilot randomized controlled trial. Transl Behav Med, 11(7), 1451-1460.

Steel, C., Korrelboom, K., Baksh, M. F., Kingdon, D., Simon, J., Wykes, T., ... & van der Gaag, M. (2020). Positive memory training for the treatment of depression in schizophrenia: A randomised controlled trial. Behaviour Research and Therapy, 135, 103734.

Stiles-Shields, C., Montague, E., Kwasny, M. J., & Mohr, D. C. (2019). Behavioral and cognitive intervention strategies delivered via coached apps for depression: Pilot trial. Psychological services, 16(2), 233-238. doi:10.1037/ser0000261

Strauss C, Hayward M, Chadwick P. Group person-based cognitive therapy for chronic depression: A pilot randomized controlled trial. British Journal of Clinical Psychology. 2012;51(3):345-50.

Strong V, Waters R, Hibberd C, Murray G, Wall L, Walker J, et al. Management of depression for people with cancer (SMaRT oncology 1): A randomised trial. Lancet. 2008;372(9632):40-8.

Sugg HVR, Richards DA, Frost J. Morita Therapy for depression (Morita Trial): a pilot randomised controlled trial. BMJ open 2018; 8(8).

Sun, Q., Xu, H., Zhang, W., Zhou, Y., & Lv, Y. (2022). Behavioral Activation Therapy for Subthreshold Depression in Stroke Patients: An Exploratory Randomized Controlled Trial. Neuropsychiatric Disease and Treatment, 18, 2795-2805.

Swartz HA, Frank E, Zuckoff A, Cyranowski JM, Houck PR, Cheng Y, et al. Brief interpersonal psychotherapy for depressed mothers whose children are receiving psychiatric treatment. American Journal of Psychiatry. 2008;165(9):1155-62.

Szumska I, Gola M, Rusanowska M, et al. Mindfulness-based cognitive therapy reduces clinical symptoms, but do not change frontal alpha asymmetry in people with major depression disorder. International Journal of Neuroscience. 2020.

Taguchi, K., Miyoshi, M., Seki, Y., Baba, S., & Shimizu, E. (2025). Effectiveness of Cognitive Behavioral Therapy Provided Through a Web Application for Subthreshold Depression, Subthreshold Insomnia, and Subthreshold Panic: Open-Labeled 6-Arm Randomized Clinical Trial Pilot Study. JMIR Form Res, 9, e63139. doi:https://doi.org/10.2196/63139

Takagaki K, Okamoto Y, Jinnin R, Mori A, Nishiyama Y, Yamamura T, et al. Behavioral activation for late adolescents with subthreshold depression: A randomized controlled trial. European Child and Adolescent Psychiatry. 2016;25(11):1171-82.

Talbot NL, Chaudron LH, Ward EA, Duberstein PR, Conwell Y, O'Hara MW, et al. A randomized effectiveness trial of interpersonal psychotherapy for depressed women with sexual abuse histories. Psychiatric Services. 2011;62(4):374-80.

Tao, Z., Li, S., Nie, J., Ni, Z., Lei, J., & Ma, H. (2025). A Novel CBGT Model for Anxiety and Depression in Patients with Pulmonary Nodules. BMC Psychol, 13(1), 1095.

Taylor FG, Marshall WL. Experimental analysis of a cognitive-behavioral therapy for depression. Cognitive Therapy and Research. 1977;1(1):59-72.

Taylor CB, Conrad A, Wilhelm FH, Strachowski D, Khaylis A, Neri E, et al. Does improving mood in depressed patients alter factors that may affect cardiovascular disease risk? Journal of Psychiatric Research. 2009;43(16):1246-52.

Teasdale JD, Fennell MJ, Hibbert GA, Amies PL. Cognitive therapy for major depressive disorder in primary care. The British Journal of Psychiatry. 1984;144:400-6.

Teichman Y, Bar-el Z, Shor H, Sirota P, Elizur A. A comparison of two modalities of cognitive therapy (individual and marital) in treating depression. Psychiatry. 1995;58(2):136-48.

Teri L, Logsdon RG, Uomoto J, McCurry SM. Behavioral treatment of depression in dementia patients: A controlled clinical trial. The journals of gerontology Series B, Psychological sciences and social sciences. 1997;52(4):P159-66.

Thielecke, J., Buntrock, C., Titzler, I., Braun, L., Freund, J., Berking, M., . . . Ebert, D. D. (2024). Telephone coaching for the prevention of depression in farmers: Results from a pragmatic randomized controlled trial. J Telemed Telecare, 30(6), 918-930.

Thomas, S. A., Drummond, A. E., Lincoln, N. B., Palmer, R. L., das Nair, R., Latimer, N. R., . . . Topcu, G. (2019). Behavioural activation therapy for post-stroke depression: the BEADS feasibility RCT. Health Technol Assess, 23(47), 1-176.

Titov N, Andrews G, Davies M, McIntyre K, Robinson E, Solley K. Internet treatment for depression: A randomized controlled trial comparing clinician vs. technician assistance. PLoS One. 2010;5(6):e10939.

Titov N, Dear BF, Ali S, Zou JB, Lorian CN, Johnston L, et al. Clinical and cost-effectiveness of therapist-guided internet-delivered cognitive behavior therapy for older adults with symptoms of depression: A randomized controlled trial. Behavior Therapy. 2015;46(2):193-205.

Tobin, K., Davey-Rothwell, M. A., Nonyane, B. A. S., Knowlton, A., Wissow, L., & Latkin, C. A. (2017). RCT of an integrated CBT-HIV intervention on depressive symptoms and HIV risk.

Tong, P., Bu, P., Yang, Y., Dong, L., Sun, T., & Shi, Y. (2019). Group cognitive behavioural therapy can reduce stigma and improve treatment compliance in major depressive disorder patients. Early intervention in psychiatry. doi:10.1111/eip.12841

Tønning, M. L., et al. (2021). "The effect of smartphone-based monitoring and treatment on the rate and duration of psychiatric readmission in patients with unipolar depressive disorder: The RADMIS randomized controlled trial." J Affect Disord 282: 354-363.

Tovote KA, Fleer J, Snippe E, Peeters A, Emmelkamp PMG, Sanderman R, et al. Individual mindfulness-based cognitive therapy and cognitive behavior therapy for treating depressive symptoms in patients with diabetes: Results of a randomized controlled trial. Diabetes care. 2014;37(9):2427-34.

Town, J., Abbass, A., Stride, C., & Bernier, D. (2017). A randomised controlled trial of Intensive Short-Term Dynamic Psychotherapy for treatment resistant depression: the Halifax Depression Study. Journal of Affective Disorders, 214, 15-25.

Trevillion K, Ryan EG, Pickles A, et al. An exploratory parallel-group randomised controlled trial of antenatal Guided Self-Help (plus usual care) versus usual care alone for pregnant women with depression: DAWN trial. Journal of Affective Disorders. 2020;261:187-197.

Tulbure BT, Andersson G, Salagean N, Pearce M, Koenig HG. Religious versus Conventional Internet-based Cognitive Behavioral Therapy for Depression. J Relig Health 2018; 57(5): 1634-48.

Turner A, Hambridge J, Baker A, Bowman J, McElduff P. Randomised controlled trial of group cognitive behaviour therapy versus brief intervention for depression in cardiac patients. Australian and New Zealand Journal of Psychiatry. 2013;47(3): 235-43.

Tyson GM, Range LM. Gestalt dialogues as a treatment for mild depression: Time works just as well. Journal of clinical psychology. 1987;43(2):227-31.

Unlu Ince B, Cuijpers P, t Hof E, Ballegooijen W, Christensen H, Riper H. Internet-based, culturally sensitive, problem-solving therapy for Turkish migrants with depression: Randomized controlled trial. Journal of Medical Internet Research. 2013;15(10):e227.

van Bastelaar KM, Pouwer F, Cuijpers P, Riper H, Snoek FJ. Web-based depression treatment for type 1 and type 2 diabetic patients: A randomized, controlled trial. Diabetes care. 2011;34(2):320-5.

van der Zanden, R., Kramer, J., Gerrits, R., & Cuijpers, P. (2012). Effectiveness of an online group course for depression in adolescents and young adults: a randomized trial. Journal of medical Internet research, 14(3), e86.

Van Horne, B. S., Nong, Y. H., Cain, C. M., Sampson, M., Greeley, C. S., & Puryear, L. (2022). A promising new model of care for postpartum depression: A randomised controlled trial of a brief home visitation program conducted in Houston, Texas, USA. Health Soc Care Community, 30(5), e2203-e2213. doi:10.1111/hsc.13658

Van Lieshout, R. J., Layton, H., Savoy, C. D., Brown, J. S. L., Ferro, M. A., Streiner, D. L., . . . Hanna, S. (2021). Effect of Online 1-Day Cognitive Behavioral Therapy-Based Workshops Plus Usual Care vs Usual Care Alone for Postpartum Depression: A Randomized Clinical Trial. JAMA Psychiatry, 78(11), 1200-1207. doi:10.1001/jamapsychiatry.2021.2488

Van Lieshout, R. J., Layton, H., Savoy, C. D., Haber, E., Feller, A., Biscaro, A., . . . Ferro, M. A. (2022). Public Health Nurse-delivered Group Cognitive Behavioural Therapy for Postpartum Depression: A Randomized Controlled Trial. Can J Psychiatry, 67(6), 432-440. doi:10.1177/07067437221074426

Van Lieshout RJ, Layton H, Savoy CD, Xie F, Brown JSL, Huh K, et al. In-person 1-day cognitive behavioral therapy-based workshops for postpartum depression: A randomized controlled trial. Psychological Medicine. 2023;53(14):6888-98.

van Luenen, S., Garnefski, N., Spinhoven, P., & Kraaij, V. (2018). Guided internet-based intervention for people with HIV and depressive symptoms: a randomised controlled trial in the Netherlands. The lancet HIV, 5(9), e488-e497.

van Schaik A, van Marwijk H, Adèr H, van Dyck R, de Haan M, Penninx B, et al. Interpersonal psychotherapy for elderly patients in primary care. The American Journal of Geriatric Psychiatry. 2006;14(9):777-86.

Varela-Moreno, E., Anarte-Ortiz, M. T., Jodar-Sanchez, F., Garcia-Palacios, A., Monreal-Bartolomé, A., Gili, M., . . . Mayoral-Cleries, F. (2024). Economic Evaluation of a Web Application Implemented in Primary Care for the Treatment of Depression in Patients With Type 2 Diabetes Mellitus: Multicenter Randomized Controlled Trial. JMIR mHealth and uHealth, 12, e55483. doi:https://doi.org/10.2196/55483

Vavani, B., Garnefski, N., van Luenen, S., Dusseldorp, E., Amone-P'Olak, K., Spinhoven, P., & Kraaij, V. (2025). A booklet self-help intervention for people living with hiv and depressive symptoms in botswana: A randomized controlled trial. AIDS and Behavior. doi:https://doi.org/10.1007/s10461-025-04742-7

Vázquez González FL, Otero Otero P, Torres Iglesias A, Hermida Garcìa E, Blanco Seoane V, Dìaz Fernández O. A brief problem-solving indicated-prevention intervention for prevention of depression in nonprofessional caregivers. Psicothema. 2013;25(1):87-92.

Vázquez, F. L., Torres, Á., Otero, P., Blanco, V., Díaz, O., & Estévez, L. E. (2017). Analysis of the components of a cognitive-behavioral intervention administered via conference call for preventing depression among non-professional caregivers: A pilot study. Aging Ment Health, 21(9), 938-946.

Vázquez FL, López L, Torres Á J, et al. Analysis of the Components of a Cognitive-Behavioral Intervention for the prevention of Depression Administered via Conference Call to Nonprofessional Caregivers: A Randomized Controlled Trial. Int J Environ Res Public Health. 2020;17(6).

Vázquez, F. L., Blanco, V., Hita, I., Torres, Á. J., Otero, P., Páramo, M., & Salmerón, M. (2023). Efficacy of a Cognitive Behavioral Intervention for the Prevention of Depression in Nonprofessional Caregivers Administered through a Smartphone App: A Randomized Controlled Trial. Journal of Clinical Medicine, 12(18).

Verduyn C, Barrowclough C, Roberts J, Tarrier N, Harrington R. Maternal depression and child behaviour problems: Randomised placebo-controlled trial of a cognitive-behavioural group intervention. British Journal of Psychiatry. 2003;183(OCT.):342-8.

Vernmark K, Lenndin J, Bjärehed J, Carlsson M, Karlsson J, Oberg J, et al. Internet administered guided self-help versus individualized e-mail therapy: A randomized trial of two versions of CBT for major depression. Behaviour Research and Therapy. 2010;48(5):368-76.

Victor-Aigbodion, V., Eseadi, C., Ardi, Z., Sewagegn, A. A., Ololo, K., Abonor, L. B., . . . Effanga, O. A. (2023). Effectiveness of rational emotive behavior therapy in reducing depression among undergraduate medical students. Medicine (Baltimore), 102(4).

Vigod, S. N., Slyfield Cook, G., Macdonald, K., Hussain-Shamsy, N., Brown, H. K., de Oliveira, C., . . . Dennis, C.-L. (2021). Mother matters: Pilot randomized wait-list controlled trial of an online therapist-facilitated discussion board and support group for postpartum depression symptoms. Depress Anxiety, 38(8), 816-825. doi:10.1002/da.23163

Vitriol VG, Ballesteros ST, Florenzano RU, Weil KP, Benadof DF. Evaluation of an outpatient intervention for women with severe depression and a history of childhood trauma. Psychiatric Services. 2009;60(7):936-42.

Warmerdam L, Straten A, Twisk J, Riper H, Cuijpers P. Internet-based treatment for adults with depressive symptoms: Randomized controlled trial. Journal of Medical Internet Research. 2008;10(4):e44.

Watt LM, Cappeliez P. Integrative and instrumental reminiscence therapies for depression in older adults: Intervention strategies and treatment effectiveness. Aging and Mental Health. 2000;4(2):166-77.

Weissman MM, Prusoff BA, Dimascio A, Neu C, Goklaney M, Klerman GL. The efficacy of drugs and psychotherapy in the treatment of acute depressive episodes. American Journal of Psychiatry. 1979;136(4b):555-8.

Westerhof, G. J., Lamers, S. M. A., Postel, M. G., & Bohlmeijer, E. T. (2019). Online Therapy for Depressive Symptoms: An Evaluation of Counselor-Led and Peer-Supported Life Review Therapy. The Gerontologist, 59(1), 135-146.

Wickberg B, Hwang CP. Counselling of postnatal depression: A controlled study on a population based Swedish sample. Journal of Affective Disorders. 1996;39(3):209-16.

Wiebe, S. A., Johnson, S. M., Allan, R., Campbell, T. L., Greenman, P. S., Fairweather, D. R., . . . Tasca, G. A. (2025). A randomized controlled trial of emotionally focused individual therapy (EFIT) for depression and anxiety. Psychotherapy (Chic), 62(3), 414-423.

Wiechers M, Strupf M, Bajbouj M, Böge K, Karnouk C, Goerigk S, et al. Empowerment group therapy for refugees with affective disorders: results of a multicenter randomized controlled trial. Eur Psychiatry. 2023;66(1):e64.

Wiersma JE, Schaik DJF, Hoogendorn AW, Dekker JJ, Van HL, Schoevers RA, et al. The effectiveness of the cognitive behavioral analysis system of psychotherapy for chronic depression: A randomized controlled trial. Psychotherapy and Psychosomatics. 2014;83(5):263-9.

Wiklund I, Mohlkert P, Edman G. Evaluation of a brief cognitive intervention in patients with signs of postnatal depression: A randomized controlled trial. Acta obstetricia et gynecologica Scandinavica. 2010;89(8):1100-4.

Williams JW, Barrett J, Oxman T, Frank E, Katon W, Sullivan M, et al. Treatment of dysthymia and minor depression in primary care: A randomized controlled trial in older adults. JAMA. 2000;284(12):1519-26.

Williams C, Wilson P, Morrison J, McMahon A, Andrew W, Allan L, et al. Guided self-help cognitive behavioural therapy for depression in primary care: A randomised controlled trial. PLoS One. 2013;8(1):e52735.

Williams C, McClay CA, Matthews L, et al. Community-based group guided self-help intervention for low mood and stress: Randomised controlled trial. British Journal of Psychiatry 2018; 212(2): 88-95.

Wilson PH, Goldin JC, Charbonneau-Powis M. Comparative efficacy of behavioral and cognitive treatments of depression. Cognitive Therapy and Research. 1983;7(2):111-24.

Winnebeck, E., Fissler, M., Gärtner, M., Chadwick, P., & Barnhofer, T. (2017). Brief training in mindfulness meditation reduces symptoms in patients with a chronic or recurrent lifetime history of depression: A randomized controlled study. Behaviour Research and Therapy, 99, 124-130. doi:10.1016/j.brat.2017.10.005

Wollersheim JP, Wilson GL. Group treatment of unipolar depression: A comparison of coping, supportive, bibliotherapy, and delayed treatment groups. Professional Psychology: Research and Practice. 1991;22(6):496.

Wong DF. Cognitive and health-related outcomes of group cognitive behavioural treatment for people with depressive symptoms in Hong Kong: Randomized wait-list control study. Australian and New Zealand Journal of Psychiatry. 2008;42(8):702-11.

Wong DF. Cognitive behavioral treatment groups for people with chronic depression in Hong Kong: A randomized wait-list control design. Depression and anxiety. 2008;25(2):142-8.

Wong SYS, Sun YY, Chan ATY, et al. Treating Subthreshold Depression in Primary Care: a Randomized Controlled Trial of Behavioral Activation With Mindfulness. Annals of family medicine 2018; 16(2): 111-9.

Wright JH, Wright AS, Albano AM, Basco MR, Goldsmith LJ, Raffield T, et al. Computer-assisted cognitive therapy for depression: Maintaining efficacy while reducing therapist time. American Journal of Psychiatry. 2005;162(6):1158-64.

Wright, J. H., Owen, J., Eells, T. D., Antle, B., Bishop, L. B., Girdler, R., . . . Ali, S. (2022). Effect of Computer-Assisted Cognitive Behavior Therapy vs Usual Care on Depression Among Adults in Primary Care: A Randomized Clinical Trial. JAMA Netw Open, 5(2), e2146716. doi:10.1001/jamanetworkopen.2021.46716

Wuthrich VM, Rapee RM. Randomised controlled trial of group cognitive behavioural therapy for comorbid anxiety and depression in older adults. Behaviour Research and Therapy. 2013;51(12):779-86.

Wuthrich VM, Rapee RM, Kangas M, Perini S. Randomized controlled trial of group cognitive behavioral therapy compared to a discussion group for co-morbid anxiety and depression in older adults. Psychological Medicine. 2016;46(4):785-95.

Xiang, X., Kayser, J., Turner, S., Ash, S., & Himle, J. A. (2024). Layperson-Supported, Web-Delivered Cognitive Behavioral Therapy for Depression in Older Adults: Randomized Controlled Trial. Journal of Medical Internet Research, 26(1).

Xie, J., He, G., Ding, S., Pan, C., Zhang, X., Zhou, J., & Iennaco, J. D. (2019). A randomized study on the effect of modified behavioral activation treatment for depressive symptoms in rural left-behind elderly. Psychotherapy research : journal of the Society for Psychotherapy Research, 29(3), 372-382. doi:10.1080/10503307.2017.1364444

Yan S, Zhong S, Lyu S, Lai S, Zhang Y, Luo Y, et al. A randomized trial of virtual reality eye movement desensitization and reprocessing therapy for major depressive disorder with childhood trauma: A 3-month follow-up study. Psychological Trauma: Theory, Research, Practice, and Policy. 2025.

Yang X, Zhao J, Chen Y, Zu S, Zhao J. Comprehensive self-control training benefits depressed college students: a six-month randomized controlled intervention trial. Journal of affective disorders 2018; 226: 251-60.

Yang, L., Nan, Y., Zhou, J., Sun, J., Huo, L., Liu, F., . . . Li, X. (2025). The effectiveness of a nurse-assisted digital thinking healthy program for postpartum depression: A randomized controlled trial. Psychiatry Research, 351.

Yeung, A., Wang, F., Feng, F., Zhang, J., Cooper, A., Hong, L., . . . Fava, M. (2017). Outcomes of an online computerized cognitive behavioral treatment program for treating chinese patients with depression: A pilot study. Asian journal of psychiatry.

Ying, Y., Ji, Y., Kong, F., Wang, M., Chen, Q., Wang, L., . . . Ruan, L. (2022). Efficacy of an internet-based cognitive behavioral therapy for subthreshold depression among Chinese adults: a randomized controlled trial. Psychological medicine, 1-11.

Yuan J, Yin Y, Tang X, et al. Culturally adapted and lay-delivered cognitive behaviour therapy for older adults with depressive symptoms in rural China: a pilot trial. Behavioural and cognitive psychotherapy. 2020:1-5.

Zemestani M, Davoodi I, Honarmand MM, Zargar Y, Ottaviani C. Comparative effects of group metacognitive therapy versus behavioural activation in moderately depressed students. Journal of Mental Health. 2016;25(6):479-85.

Zemestani, M., & Fazeli Nikoo, Z. (2020). Effectiveness of mindfulness-based cognitive therapy for comorbid depression and anxiety in pregnancy: a randomized controlled trial. Archives of Women's Mental Health. doi:10.1007/s00737-019-00962-8

Zhao, Y., Munro-Kramer, M. L., Shi, S., Wang, J., & Zhao, Q. (2019). Effects of antenatal depression screening and intervention among Chinese high-risk pregnant women with medically defined complications: A randomized controlled trial. Early intervention in psychiatry, 13(5), 1090-1098. doi:10.1111/eip.12731

Zhao, Y., Lin, Q., & Wang, J. (2021). An evaluation of a prenatal individualised mixed management intervention addressing breastfeeding outcomes and postpartum depression: A ramdomised controlled trial. J Clin Nurs, 30(9-10), 1347-1359.

Zu S, Xiang Y-T, Liu J, Zhang L, Wang G, Ma X, et al. A comparison of cognitive-behavioral therapy, antidepressants, their combination and standard treatment for Chinese patients with moderate–severe major depressive disorders. Journal of Affective Disorders. 2014;152-154:262-7.

### Supplement H. Study characteristics

##### Table H1

*Aggregated study characteristics*

| Characteristic | N(%), range, Mean (SD) |
|---|---|
| Publication year, mean (SD) | 2013.7 (SD = 9.7) |
| Publication year, range | 1977 to 2026 |
| Total sample size, mean (SD) | 128.7 (SD = 149.6) |
| Total sample size, range | 14 to 1899 |
| Mean age, mean (SD) | 43.3 (SD = 14.3) |
| Percentage women, mean (SD) | 72.7 (SD = 21.6) |
| **Psychotherapy type** | |
| Cognitive behavioural therapy | 261 (50.8%) |
| Behavioural activation | 39 (7.6%) |
| Interpersonal psychotherapy | 30 (5.8%) |
| Problem-solving therapy | 28 (5.4%) |
| Psychodynamic therapy | 12 (2.3%) |
| Life review therapy | 16 (3.1%) |
| Supportive therapy | 11 (2.1%) |
| Third-wave therapies | 51 (9.9%) |
| Other | 66 (12.8%) |
| **Delivery format** | |
| Individual | 165 (32.1%) |
| Group | 146 (28.4%) |
| Guided self-help | 114 (22.2%) |
| Telephone | 24 (4.7%) |
| Other | 65 (12.6%) |
| **Control condition** | |
| Waitlist | 192 (37.4%) |
| Care as usual | 239 (46.5%) |
| Other control | 83 (16.1%) |
| **Inclusion criterion for depression** | |
| Clinical diagnosis | 230 (44.7%) |
| Cut-off/subclinical score | 284 (55.3%) |
| **Recruitment setting** | |
| Community | 208 (40.5%) |
| Clinical | 121 (23.5%) |
| Other/not reported | 185 (36%) |
| **Country income level** | |
| High income | 396 (77%) |
| Low/middle income | 116 (22.6%) |
| Not classifiable | 2 (0.4%) |

### Supplement I. Sensitivity analysis restricted to studies published after CONSORT guidelines

#### Table I1

*Prevalence of RoB*

| Domain | Low risk | Some concerns | High risk |
|---|---|---|---|
| **Full dataset (N = 514)** | | | |
| Domain 1 - Randomization process | 263 (51.2%) | 234 (45.5%) | 17 (3.3%) |
| Domain 2 - Deviations from intended interventions | 365 (71%) | 16 (3.1%) | 133 (25.9%) |
| Domain 3 - Missing outcome data | 258 (50.2%) | 71 (13.8%) | 185 (36%) |
| Domain 4 - Measurement of the outcome | 479 (93.2%) | 0 (0%) | 35 (6.8%) |
| Domain 5 - Selective outcome reporting | 131 (25.5%) | 360 (70%) | 23 (4.5%) |
| Overall RoB score | 74 (14.4%) | 172 (33.5%) | 268 (52.1%) |
| **Restricted to studies published from 2001 onwards (n = 461)** | | | |
| Domain 1 - Randomization process | 257 (55.7%) | 187 (40.6%) | 17 (3.7%) |
| Domain 2 - Deviations from intended interventions | 352 (76.4%) | 12 (2.6%) | 97 (21%) |
| Domain 3 - Missing outcome data | 249 (54%) | 68 (14.8%) | 144 (31.2%) |
| Domain 4 - Measurement of the outcome | 436 (94.6%) | 0 (0%) | 25 (5.4%) |
| Domain 5 - Selective outcome reporting | 131 (28.4%) | 307 (66.6%) | 23 (5%) |
| Overall RoB score | 74 (16.1%) | 165 (35.8%) | 222 (48.2%) |

#### Table I2

*Trial characteristics associated with Risk of Bias. Restricted to studies published from 2001 onwards (n = 459)*

| Predictor | OR | 95% CI | p |
|---|---|---|---|
| **Overall RoB score** | | | |
| Control: waitlist vs. other | 1.15 | [0.765, 1.716] | 0.510 |
| Psychotherapy: CBT vs. other | 1.35 | [0.931, 1.952] | 0.114 |
| Format: group vs. individual | 1.43 | [0.888, 2.318] | 0.141 |
| Format: guided self-help vs. individual* | 0.40 | [0.233, 0.673] | < .001 |
| Format: other vs. individual | 0.99 | [0.57, 1.72] | 0.972 |
| Diagnosis: cut-off vs. clinical | 1.31 | [0.902, 1.905] | 0.155 |
| Income: low/middle vs. high | 1.13 | [0.74, 1.729] | 0.569 |
| Sample size (log)* | 0.45 | [0.344, 0.583] | < .001 |
| **Domain 1. Randomization process** | | | |
| Control: waitlist vs. other | 0.98 | [0.642, 1.480] | 0.904 |
| Psychotherapy: CBT vs. other | 1.37 | [0.930, 2.032] | 0.111 |
| Format: group vs. individual | 0.91 | [0.568, 1.473] | 0.713 |
| Format: guided self-help vs. individual* | 0.30 | [0.166, 0.552] | < .001 |
| Format: other vs. individual | 0.68 | [0.383, 1.202] | 0.184 |
| Diagnosis: cut-off vs. clinical | 1.36 | [0.921, 2.020] | 0.121 |
| Income: low/middle vs. high | 1.41 | [0.906, 2.184] | 0.129 |
| Sample size (log)* | 0.65 | [0.496, 0.850] | 0.002 |
| **Domain 2. Deviations from intended interventions** | | | |
| Control: waitlist vs. other | 0.89 | [0.533, 1.49] | 0.661 |
| Psychotherapy: CBT vs. other | 1.14 | [0.708, 1.823] | 0.597 |
| Format: group vs. individual | 1.21 | [0.686, 2.144] | 0.506 |
| Format: guided self-help vs. individual | 0.51 | [0.229, 1.134] | 0.099 |
| Format: other vs. individual | 1.47 | [0.759, 2.842] | 0.254 |
| Diagnosis: cut-off vs. clinical | 1.57 | [0.975, 2.53] | 0.064 |

| | | | |
|---|---|---|---|
| Income: low/middle vs. high* | 3.52 | [2.158, 5.756] | < .001 |
| Sample size (log)* | 0.49 | [0.348, 0.69] | < .001 |
| **Domain 3. Missing outcome data** | | | |
| Control: waitlist vs. other | 1.28 | [0.854, 1.909] | 0.233 |
| Psychotherapy: CBT vs. other | 1.14 | [0.786, 1.651] | 0.490 |
| Format: group vs. individual | 1.04 | [0.653, 1.659] | 0.866 |
| Format: guided self-help vs. individual* | 0.56 | [0.323, 0.981] | 0.043 |
| Format: other vs. individual | 0.84 | [0.485, 1.461] | 0.539 |
| Diagnosis: cut-off vs. clinical* | 1.61 | [1.104, 2.358] | 0.013 |
| Income: low/middle vs. high | 1.12 | [0.736, 1.718] | 0.588 |
| Sample size (log)* | 0.54 | [0.417, 0.701] | < .001 |
| **Domain 4. Measurement of the outcome** | | | |
| Control: waitlist vs. other | 1.16 | [0.459, 2.930] | 0.753 |
| Psychotherapy: CBT vs. other | 0.46 | [0.183, 1.145] | 0.095 |
| Format: group vs. individual | 0.64 | [0.236, 1.736] | 0.381 |
| Format: guided self-help vs. individual | 0.14 | [0.017, 1.132] | 0.065 |
| Format: other vs. individual | 0.63 | [0.194, 2.060] | 0.447 |
| Diagnosis: cut-off vs. clinical | 0.50 | [0.213, 1.189] | 0.117 |
| Income: low/middle vs. high | 0.54 | [0.175, 1.668] | 0.285 |
| Sample size (log) | 0.62 | [0.329, 1.149] | 0.127 |
| **Domain 5. Selective outcome reporting** | | | |
| Control: waitlist vs. other | 1.05 | [0.67, 1.655] | 0.823 |
| Psychotherapy: CBT vs. other | 1.44 | [0.951, 2.192] | 0.085 |
| Format: group vs. individual | 1.40 | [0.808, 2.426] | 0.231 |
| Format: guided self-help vs. individual* | 0.29 | [0.16, 0.519] | < .001 |
| Format: other vs. individual | 1.09 | [0.582, 2.046] | 0.785 |
| Diagnosis: cut-off vs. clinical | 1.09 | [0.716, 1.65] | 0.697 |
| Income: low/middle vs. high | 1.22 | [0.751, 1.979] | 0.422 |
| Sample size (log)* | 0.57 | [0.429, 0.764] | < .001 |

##### Table I3

*Association Between Risk of Bias and Effect Size Estimates. Sensitivity analysis: pairwise contrasts restricted to trials published from 2001 onwards (n = 461, 756 effect sizes)*

| Model | Contrast | Δg | SE | 95% CI |
|---|---|---|---|---|
| **Overall RoB score** | | | | |
| Unadjusted | Low risk vs. some concerns | −0.201 | 0.055 | [−0.312, −0.091] |
| | Low risk vs. high risk | −0.335 | 0.059 | [−0.452, −0.217] |
| | Some concerns vs. high risk | −0.134 | 0.060 | [−0.254, −0.013] |
| Adjusted | Low risk vs. some concerns | −0.206 | 0.055 | [−0.315, −0.097] |
| | Low risk vs. high risk | −0.328 | 0.059 | [−0.444, −0.212] |
| | Some concerns vs. high risk | −0.122 | 0.060 | [−0.240, −0.004] |
| **Domain 1: Randomization process** | | | | |
| Unadjusted | Low risk vs. some concerns | −0.161 | 0.058 | [−0.287, −0.036] |
| | Low risk vs. high risk | −0.170 | 0.132 | [−0.458, 0.118] |
| | Some concerns vs. high risk | −0.008 | 0.130 | [−0.291, 0.274] |
| Adjusted | Low risk vs. some concerns | −0.162 | 0.056 | [−0.284, −0.041] |
| | Low risk vs. high risk | −0.196 | 0.137 | [−0.492, 0.099] |
| | Some concerns vs. high risk | −0.034 | 0.133 | [−0.323, 0.255] |
| **Domain 2: Deviations from intended interventions** | | | | |
| Unadjusted | Low risk vs. some concerns | −0.230 | 0.165 | [−0.593, 0.134] |

| | | | | |
|---|---|---|---|---|
| | Low risk vs. high risk | −0.387 | 0.095 | [−0.595, −0.178] |
| | Some concerns vs. high risk | −0.157 | 0.185 | [−0.565, 0.251] |
| Adjusted | Low risk vs. some concerns | −0.291 | 0.173 | [−0.673, 0.091] |
| | Low risk vs. high risk | −0.406 | 0.093 | [−0.610, −0.202] |
| | Some concerns vs. high risk | −0.115 | 0.193 | [−0.540, 0.310] |
| **Domain 3: Missing outcome data** | | | | |
| Unadjusted | Low risk vs. some concerns | 0.020 | 0.075 | [−0.129, 0.169] |
| | Low risk vs. high risk | −0.118 | 0.073 | [−0.263, 0.027] |
| | Some concerns vs. high risk | −0.138 | 0.083 | [−0.303, 0.027] |
| Adjusted | Low risk vs. some concerns | 0.013 | 0.075 | [−0.137, 0.163] |
| | Low risk vs. high risk | −0.093 | 0.072 | [−0.237, 0.050] |
| | Some concerns vs. high risk | −0.106 | 0.084 | [−0.274, 0.061] |
| **Domain 4: Measurement of the outcome** | | | | |
| Unadjusted | Low risk vs. high risk | −0.281 | 0.100 | [−0.488, −0.074] |
| Adjusted | Low risk vs. high risk | −0.285 | 0.099 | [−0.490, −0.080] |
| **Domain 5: Selective outcome reporting** | | | | |
| Unadjusted | Low risk vs. some concerns | −0.188 | 0.058 | [−0.308, −0.069] |
| | Low risk vs. high risk | −0.216 | 0.133 | [−0.492, 0.060] |
| | Some concerns vs. high risk | −0.028 | 0.134 | [−0.306, 0.250] |
| Adjusted | Low risk vs. some concerns | −0.196 | 0.057 | [−0.313, −0.079] |
| | Low risk vs. high risk | −0.196 | 0.130 | [−0.464, 0.072] |
| | Some concerns vs. high risk | 0.000 | 0.130 | [−0.269, 0.270] |

*Note.* Δg = difference in pooled Hedges' g between RoB categories. Adjusted models control for control group type (waitlist vs. other). CR2 = cluster-robust variance estimation. Results are largely consistent with the full sample analyses (N = 514 trials, 941 effect sizes).

### Supplement J. Trends in Risk of Bias Over Time

#### *Domain-Level Results and Model Coefficients*

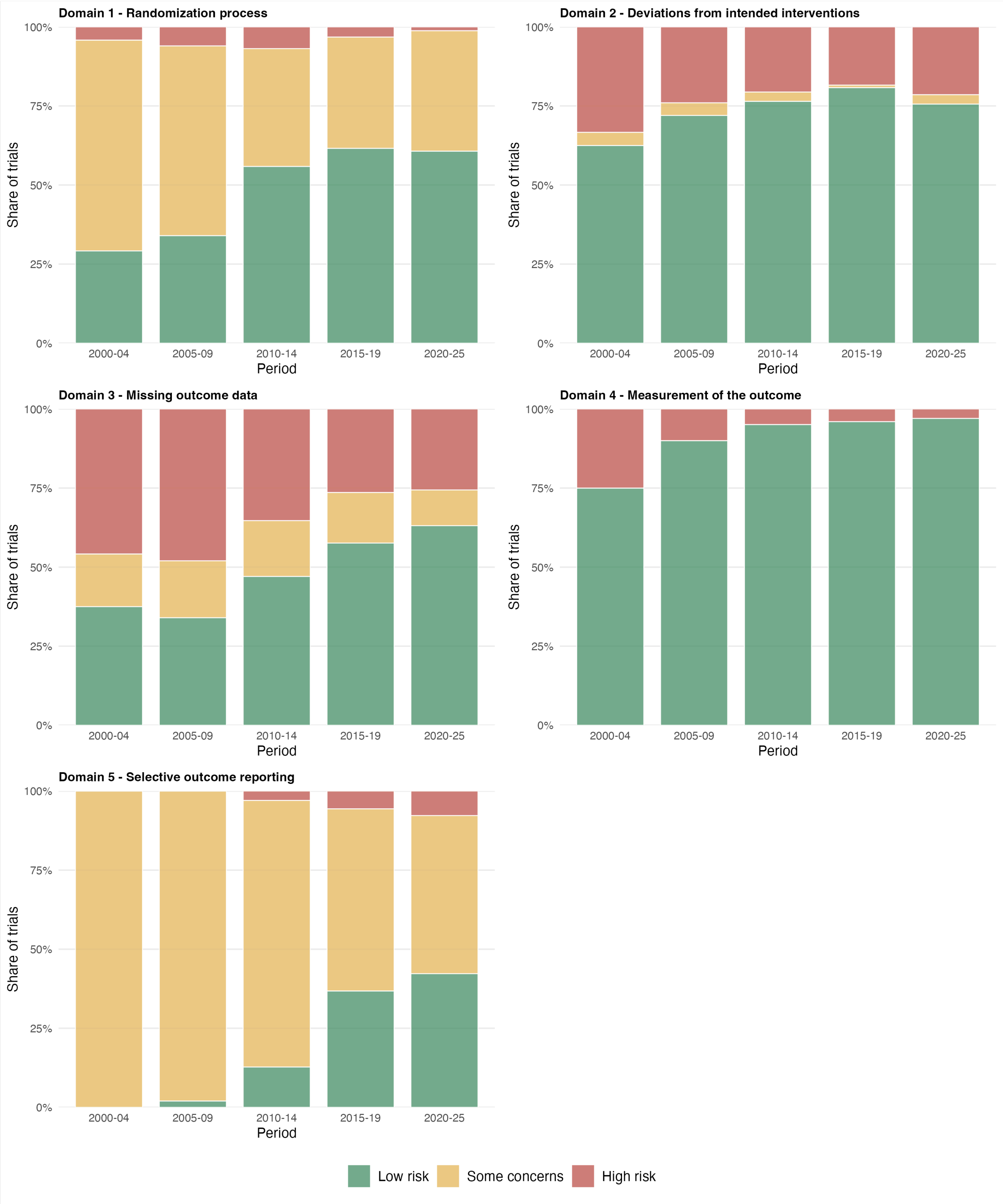

**Figure J1.** Share of trials rated as low risk, some concerns, or high risk of bias per 5-year period for each of the five domains (N = 514). Proportions are based on the study-level risk of bias score for all domains. For Domain 5 (selective outcome reporting), ratings other than some concerns are only possible when a prospective trial registration or protocol is available, explaining the absence of low risk and high risk ratings in earlier periods.

*Poisson regression model coefficients for trends in risk of bias over time*

Separate Poisson regression models were fitted for the overall risk of bias score and each of the five domains, with annual trial counts as the outcome. Each model included publication year (scaled to mean zero and unit standard deviation) and RoB category as predictors, along with their interaction. The interaction term tests whether the growth rate of a given category differs significantly from that of the high risk category (reference) over time. A positive interaction coefficient indicates that the category is growing faster than high risk, that is, its proportion is increasing over time. A negative coefficient indicates it is growing slower. Coefficients represent change per standard deviation of publication year (approximately 10 years in this dataset).

**Table J1**

*Poisson regression model coefficients for trends in risk of bias over time (N = 514)*

| Term | β | SE | z | p |
|---|---|---|---|---|
| **Overall RoB score** | | | | |
| (Intercept) | 1.525 | 0.075 | 20.414 | < .001 |
| Year | 0.678 | 0.071 | 9.573 | < .001 |
| Low risk vs. high risk | −2.467 | 0.309 | −7.999 | < .001 |
| Some concerns vs. high risk | −0.807 | 0.144 | −5.590 | < .001 |
| Year × low risk | 1.250 | 0.245 | 5.107 | < .001 |
| Year × some concerns | 0.476 | 0.129 | 3.688 | < .001 |
| **Domain 1. Randomization process** | | | | |
| (Intercept) | −1.377 | 0.334 | −4.120 | < .001 |
| Year | 0.890 | 0.306 | 2.912 | .004 |
| Low risk vs. high risk | 2.290 | 0.354 | 6.472 | < .001 |
| Some concerns vs. high risk | 2.816 | 0.343 | 8.212 | < .001 |
| Year × low risk | 0.504 | 0.321 | 1.572 | .116 |
| Year × some concerns | −0.297 | 0.314 | −0.946 | .344 |
| **Domain 2. Deviations from intended interventions** | | | | |
| (Intercept) | 0.917 | 0.098 | 9.393 | < .001 |
| Year | 0.507 | 0.095 | 5.335 | < .001 |
| Low risk vs. high risk | 0.527 | 0.130 | 4.039 | < .001 |
| Some concerns vs. high risk | −2.128 | 0.300 | −7.085 | < .001 |
| Year × low risk | 0.677 | 0.121 | 5.587 | < .001 |
| Year × some concerns | 0.020 | 0.292 | 0.069 | .945 |
| **Domain 3. Missing outcome data** | | | | |
| (Intercept) | 1.229 | 0.084 | 14.594 | < .001 |
| Year | 0.545 | 0.082 | 6.683 | < .001 |
| Low risk vs. high risk | −0.254 | 0.140 | −1.821 | .069 |
| Some concerns vs. high risk | −1.307 | 0.199 | −6.559 | < .001 |
| Year × low risk | 0.768 | 0.125 | 6.145 | < .001 |
| Year × some concerns | 0.508 | 0.180 | 2.824 | .005 |
| **Domain 4. Measurement of the outcome** | | | | |
| (Intercept) | −0.322 | 0.174 | −1.854 | .064 |
| Year | 0.236 | 0.174 | 1.356 | .175 |
| Low risk vs. high risk | 2.183 | 0.187 | 11.703 | < .001 |
| Year × low risk | 0.782 | 0.184 | 4.245 | < .001 |
| **Domain 5. Selective outcome reporting** | | | | |
| (Intercept) | −2.287 | 0.581 | −3.938 | < .001 |
| Year | 2.075 | 0.447 | 4.639 | < .001 |
| Low risk vs. high risk | 1.912 | 0.623 | 3.070 | .002 |

| | | | | |
|---|---|---|---|---|
| Some concerns vs. high risk | 4.098 | 0.584 | 7.013 | < .001 |
| Year × low risk | −0.144 | 0.481 | −0.300 | .764 |
| Year × some concerns | −1.382 | 0.452 | −3.060 | .002 |

*Note*. Publication year was standardized (centered at the mean and divided by the standard deviation, SD = 14.2 years); coefficients therefore represent the change in log count associated with a 14.2-year increase in publication year. High risk is the reference category for all category comparisons. Interaction terms (Year × category) test whether a category grew at a significantly different rate than high risk over time. Some concerns estimates for Domain 4 are not reported because this category was not observed in the data.

### Supplement K. Association Between Risk of Bias and Effect Size Estimates

**Table K1**

*Pooled effect sizes per risk of bias category. Unadjusted and adjusted models (N = 514 trials, 941 effect sizes)*

| Domain / RoB category | Model | *g* | 95% CI | 95% PI |
|---|---|---|---|---|
| **Overall RoB score** | | | | |
| Low risk | Unadjusted | 0.479 | 0.342 to 0.615 | −0.72 to 1.67 |
| | Adjusted | 0.366 | 0.229 to 0.503 | −0.79 to 1.52 |
| Some concerns | Unadjusted | 0.682 | 0.594 to 0.771 | −0.51 to 1.87 |
| | Adjusted | 0.573 | 0.479 to 0.666 | −0.58 to 1.73 |
| High risk | Unadjusted | 0.817 | 0.740 to 0.893 | −0.37 to 2.01 |
| | Adjusted | 0.688 | 0.603 to 0.773 | −0.46 to 1.84 |
| **Domain 1: Randomization process** | | | | |
| Low risk | Unadjusted | 0.635 | 0.561 to 0.710 | −0.57 to 1.84 |
| | Adjusted | 0.520 | 0.439 to 0.601 | −0.64 to 1.68 |
| Some concerns | Unadjusted | 0.812 | 0.731 to 0.893 | −0.39 to 2.01 |
| | Adjusted | 0.683 | 0.594 to 0.771 | −0.48 to 1.85 |
| High risk | Unadjusted | 0.809 | 0.545 to 1.072 | −0.42 to 2.04 |
| | Adjusted | 0.719 | 0.459 to 0.979 | −0.47 to 1.91 |
| **Domain 2: Deviations from intended interventions** | | | | |
| Low risk | Unadjusted | 0.628 | 0.563 to 0.692 | −0.55 to 1.80 |
| | Adjusted | 0.503 | 0.429 to 0.576 | −0.63 to 1.64 |
| Some concerns | Unadjusted | 0.875 | 0.561 to 1.189 | −0.34 to 2.09 |
| | Adjusted | 0.757 | 0.451 to 1.063 | −0.42 to 1.93 |
| High risk | Unadjusted | 0.963 | 0.851 to 1.076 | −0.22 to 2.14 |
| | Adjusted | 0.843 | 0.728 to 0.958 | −0.29 to 1.98 |
| **Domain 3: Missing outcome data** | | | | |
| Low risk | Unadjusted | 0.678 | 0.599 to 0.756 | −0.53 to 1.89 |
| | Adjusted | 0.569 | 0.485 to 0.654 | −0.60 to 1.74 |
| Some concerns | Unadjusted | 0.657 | 0.514 to 0.800 | −0.56 to 1.87 |
| | Adjusted | 0.553 | 0.410 to 0.696 | −0.63 to 1.73 |
| High risk | Unadjusted | 0.805 | 0.710 to 0.901 | −0.40 to 2.02 |
| | Adjusted | 0.662 | 0.558 to 0.766 | −0.51 to 1.84 |
| **Domain 4: Measurement of the outcome** | | | | |
| Low risk | Unadjusted | 0.706 | 0.649 to 0.763 | −0.50 to 1.91 |
| | Adjusted | 0.583 | 0.516 to 0.650 | −0.58 to 1.75 |
| High risk | Unadjusted | 0.985 | 0.830 to 1.140 | −0.23 to 2.20 |
| | Adjusted | 0.863 | 0.705 to 1.021 | −0.31 to 2.04 |
| **Domain 5: Selective outcome reporting** | | | | |
| Low risk | Unadjusted | 0.573 | 0.465 to 0.682 | −0.63 to 1.78 |
| | Adjusted | 0.451 | 0.338 to 0.563 | −0.72 to 1.62 |
| Some concerns | Unadjusted | 0.769 | 0.702 to 0.837 | −0.43 to 1.97 |
| | Adjusted | 0.648 | 0.572 to 0.724 | −0.52 to 1.81 |
| High risk | Unadjusted | 0.789 | 0.525 to 1.053 | −0.44 to 2.02 |
| | Adjusted | 0.642 | 0.382 to 0.902 | −0.55 to 1.83 |

*Note. g* = pooled Hedges' *g*. CI = 95% confidence interval. PI = 95% prediction interval. Adjusted models control for control group type (waitlist vs. other); adjusted pooled estimates refer to trials with non-waitlist control groups.

**Table K2**

*Pairwise contrasts in pooled effect sizes across risk of bias categories, unadjusted and adjusted models (N = 514 trials, 941 effect sizes)*

| Model | Contrast | Δg | SE | 95% CI |
|---|---|---|---|---|
| **Overall RoB score** | | | | |
| Unadjusted | Low risk vs. some concerns | −0.20 | 0.05 | [−0.31, −0.09] |
| | Low risk vs. high risk | −0.34 | 0.06 | [−0.45, −0.23] |
| | Some concerns vs. high risk | −0.13 | 0.06 | [−0.25, −0.02] |
| Adjusted | Low risk vs. some concerns | −0.21 | 0.05 | [−0.32, −0.10] |
| | Low risk vs. high risk | −0.32 | 0.06 | [−0.43, −0.21] |
| | Some concerns vs. high risk | −0.12 | 0.06 | [−0.23, −0.01] |
| **Domain 1: Randomization process** | | | | |
| Unadjusted | Low risk vs. some concerns | −0.18 | 0.05 | [−0.29, −0.06] |
| | Low risk vs. high risk | −0.17 | 0.13 | [−0.46, 0.12] |
| | Some concerns vs. high risk | 0.00 | 0.13 | [−0.28, 0.29] |
| Adjusted | Low risk vs. some concerns | −0.16 | 0.05 | [−0.27, −0.05] |
| | Low risk vs. high risk | −0.20 | 0.14 | [−0.50, 0.10] |
| | Some concerns vs. high risk | −0.04 | 0.13 | [−0.33, 0.25] |
| **Domain 2: Deviations from intended interventions** | | | | |
| Unadjusted | Low risk vs. some concerns | −0.25 | 0.13 | [−0.53, 0.04] |
| | Low risk vs. high risk | −0.34 | 0.08 | [−0.50, −0.17] |
| | Some concerns vs. high risk | −0.09 | 0.15 | [−0.40, 0.23] |
| Adjusted | Low risk vs. some concerns | −0.25 | 0.14 | [−0.55, 0.05] |
| | Low risk vs. high risk | −0.34 | 0.08 | [−0.50, −0.18] |
| | Some concerns vs. high risk | −0.09 | 0.16 | [−0.42, 0.24] |
| **Domain 3: Missing outcome data** | | | | |
| Unadjusted | Low risk vs. some concerns | 0.02 | 0.07 | [−0.12, 0.16] |
| | Low risk vs. high risk | −0.13 | 0.06 | [−0.26, 0.00] |
| | Some concerns vs. high risk | −0.15 | 0.08 | [−0.30, 0.00] |
| Adjusted | Low risk vs. some concerns | 0.02 | 0.07 | [−0.13, 0.16] |
| | Low risk vs. high risk | −0.09 | 0.06 | [−0.22, 0.04] |
| | Some concerns vs. high risk | −0.11 | 0.08 | [−0.26, 0.05] |
| **Domain 4: Measurement of the outcome** | | | | |
| Unadjusted | Low risk vs. high risk | −0.28 | 0.08 | [−0.45, −0.11] |
| Adjusted | Low risk vs. high risk | −0.28 | 0.08 | [−0.45, −0.12] |
| **Domain 5: Selective outcome reporting** | | | | |
| Unadjusted | Low risk vs. some concerns | −0.20 | 0.05 | [−0.31, −0.08] |
| | Low risk vs. high risk | −0.22 | 0.13 | [−0.49, 0.06] |
| | Some concerns vs. high risk | −0.02 | 0.13 | [−0.29, 0.25] |
| Adjusted | Low risk vs. some concerns | −0.20 | 0.05 | [−0.31, −0.09] |
| | Low risk vs. high risk | −0.19 | 0.13 | [−0.46, 0.08] |
| | Some concerns vs. high risk | 0.01 | 0.13 | [−0.26, 0.27] |

*Note.* Δg = difference in pooled Hedges' g between RoB categories. Positive Δg indicates larger effect sizes in the first category. Adjusted models control for control group type (waitlist vs. other). CR2 = cluster-robust variance estimation (clubSandwich).

**Table K3**

*Heterogeneity statistics. Unadjusted and adjusted models (N = 514 trials, 941 effect sizes)*

| Model | $\tau^2$_between | $\tau^2$_within | $I^2$ (%) |
|---|---|---|---|
| **Overall RoB score** | | | |
| Unadjusted | 0.2822 | 0.0832 | 77.1 |
| Adjusted | 0.2615 | 0.0816 | 76.0 |

| | | | |
|---|---|---|---|
| **Domain 1: Randomization process** | | | |
| Unadjusted | 0.2913 | 0.0821 | 77.5 |
| Adjusted | 0.2700 | 0.0804 | 76.4 |
| **Domain 2: Deviations from intended interventions** | | | |
| Unadjusted | 0.2749 | 0.0833 | 76.8 |
| Adjusted | 0.2507 | 0.0818 | 75.4 |
| **Domain 3: Missing outcome data** | | | |
| Unadjusted | 0.2946 | 0.0830 | 77.7 |
| Adjusted | 0.2738 | 0.0816 | 76.6 |
| **Domain 4: Measurement of the outcome** | | | |
| Unadjusted | 0.2967 | 0.0800 | 77.7 |
| Adjusted | 0.2744 | 0.0784 | 76.5 |
| **Domain 5: Selective outcome reporting** | | | |
| Unadjusted | 0.2914 | 0.0832 | 77.6 |
| Adjusted | 0.2687 | 0.0816 | 76.4 |

*Note.* Three-level CHE models. $\tau^2$_between = between-study variance; $\tau^2$_within = within-study variance (across effect sizes from the same study). $I^2$ = total heterogeneity as a percentage of total variance. Adjusted models control for control group type (waitlist vs. other).

##### Table K4

*Exploratory moderation analysis: interaction between RoB group and study characteristics (N = 514 trials, 941 effect sizes)*

| Moderator | Interaction term | Estimate | SE | 95% CI |
|---|---|---|---|---|
| Control group type | Higher risk × waitlist vs. other | 0.213 | 0.149 | [−0.08, 0.51] |
| Psychotherapy type | Higher risk × CBT vs. other | 0.017 | 0.137 | [−0.25, 0.28] |
| Delivery format | Higher risk × group vs. individual | 0.102 | 0.293 | [−0.47, 0.68] |
| | Higher risk × guided self-help vs. individual | −0.051 | 0.237 | [−0.52, 0.41] |
| | Higher risk × other vs. individual | 0.208 | 0.262 | [−0.31, 0.72] |
| Diagnosis criterion | Higher risk × cut-off vs. clinical | 0.049 | 0.154 | [−0.25, 0.35] |
| Income level* | Higher risk × low/middle vs. high | 0.542 | 0.197 | [0.16, 0.93] |
| Sample size (log) | Higher risk × sample size (log) | −0.078 | 0.090 | [−0.25, 0.10] |

*Note.* Interaction estimates reflect the difference in Δg (higher risk minus low risk) as a function of each moderator. A positive estimate indicates that the association between higher RoB and larger effect sizes is stronger in the comparison group (e.g., low/middle income vs. high income). All moderators entered in separate models. Higher risk = some concerns + high risk combined. Significant moderators are marked with an asterisk.
